# Brain-Derived Neurotrophic Factor (*BDNF*) Epigenomic Modifications and Brain-Related Phenotypes in Humans: A Systematic Review

**DOI:** 10.1101/2022.09.13.22279723

**Authors:** Amery Treble-Barna, Lacey W. Heinsberg, Zachary Stec, Stephen Breazeale, Tara S. Davis, Aboli A. Kesbhat, Ansuman Chattopadhyay, Helena M. VonVille, Andrea M. Ketchum, Keith Owen Yeates, Patrick M. Kochanek, Daniel E. Weeks, Yvette P. Conley

**Author notes:** Corresponding Author: Department of Physical Medicine & Rehabilitation, School of Medicine, University of Pittsburgh, PA 15261, USA.

## Abstract

Epigenomic modifications of the brain-derived neurotrophic factor (*BDNF*) gene have been postulated to underlie the pathogenesis of neurodevelopmental, psychiatric, and neurological conditions. This systematic review summarizes current evidence investigating the association of *BDNF* epigenomic modifications (DNA methylation, non-coding RNA, histone modifications) with brain-related phenotypes in humans. A novel contribution is our creation of an open access web-based application, the *BDNF* DNA Methylation Map, to interactively visualize specific positions of CpG sites investigated across all studies for which relevant data were available. Our literature search of four databases through September 27, 2021 returned 1,701 articles, of which 153 met inclusion criteria. Our review revealed exceptional heterogeneity in methodological approaches, hindering the identification of clear patterns of robust and/or replicated results. We summarize key findings and provide recommendations for future epigenomic research. The existing literature appears to remain in its infancy and requires additional rigorous research to fulfill its potential to explain *BDNF*-linked risk for brain-related conditions and improve our understanding of the molecular mechanisms underlying their pathogenesis.

**Highlights:**

- 153 articles examined *BDNF* epigenomic modifications and brain-related phenotypes
- Novel *BDNF* DNA Methylation Map allows users to interactively visualize CpGs
- *BDNF* epigenomics lack robust/replicated results due to methodological heterogeneity

## 1. Introduction

Brain-derived neurotrophic factor (BDNF) is a well-studied member of the neurotrophin family of growth factors. Released pre- and post-synaptically from neurons, BDNF mediates apoptosis, neuronal differentiation, outgrowth of neurites, cell survival, and synaptic strengthening (Barker, 2009; Lu et al., 2005). BDNF is therefore essential to processes of brain development, neuroplasticity, and neuronal survival (Chen et al., 2004; Huang and Reichardt, 2001), as well as complex cognitive functions (Cortés-Mendoza et al., 2013; Koven and Collins, 2014; Leckie et al., 2014; Manju et al., 2017; van der Kolk et al., 2015; Wiłkość et al., 2016). Variation in BDNF concentration (Camuso et al., 2022; Cattaneo et al., 2016; Fernandes et al., 2014; Molendijk et al., 2014), differences in *BDNF* gene polymorphisms (Brown et al., 2020; Hong et al., 2011; Notaras et al., 2015), and—most recently—epigenomic modifications of the *BDNF* gene (Chen and Chen, 2017; Kader et al., 2018; Lin and Huang, 2020; Poon et al., 2021; Teroganova et al., 2016) have been implicated in the pathogenesis of a multitude of neurodevelopmental, psychiatric, and neurological (henceforth referred to as “brain-related”) conditions.

The human *BDNF* gene is mapped to chromosome 11p13 and is composed of 11 exons and nine tissue- and brain region-specific functional promoters (Pruunsild et al., 2007). The most commonly examined *BDNF* single nucleotide polymorphism (SNP) produces a valine-to-methionine substitution at codon 66 (Val66Met; rs6265), with the Met allele associated with reduced activity-dependent secretion of BDNF (Egan et al., 2003). While hundreds of studies have examined the association between rs6265 (and other BDNF SNPs) and brain-related conditions, results have been mixed, as is observed in many complex phenotypes (Ahmed et al., 2015; Ferreira Fratelli et al., 2021; Pereira et al., 2018). Also, when examined in consortium studies with very large sample sizes, some of the *BDNF* associations previously seen in small candidate gene studies have not replicated (Border et al., 2019; Johnson et al., 2017). Thus, substantial unexplained heterogeneity in risk for brain-related conditions remains that is not accounted for by genetic variation alone or in combination with environmental risk factors (Cattaneo et al., 2016; Dall’Aglio et al., 2018; Di Carlo et al., 2019).

The field of epigenomics has risen to the forefront as a potential means to account for some of the unexplained heterogeneity in the risk for brain-related conditions, and to foster our understanding of the molecular mechanisms underlying their pathogenesis (Diwadkar et al., 2014; Millan, 2013). Broadly defined, epigenomics involves potentially heritable biochemical processes that regulate gene expression without altering the corresponding primary DNA sequence (Nikolova and Hariri, 2015). Through epigenomic processes, the biological and social environments of an individual impact when and to what extent genes are expressed within each cell type. The most investigated epigenomic modification is DNA methylation (DNAm), which involves the addition of a methyl group to cytosine-guanine dinucleotides (CpG). Higher DNAm in CpG rich promoters or gene regulatory regions is usually (but not always) associated with lower gene expression (Jang et al., 2017). Two additional epigenomic modifications of increasing interest are (1) non-coding RNAs, such as microRNA (miRNAs), which bind to mRNAs to suppress gene expression post-transcriptionally (Chuang and Jones, 2007; Yao et al., 2019) and (2) histone modifications, involving the addition or removal of chemical groups to nucelosomes around which DNA is wound, thereby affecting chromatin structure and resulting in variations in DNA expression (Bannister and Kouzarides, 2011).

Because of its pleiotropic effects in the brain, much can potentially be learned by surveying the evidence for epigenomic effects of the *BDNF* gene across many brain-related conditions. Such a transdiagnostic approach (Notaras and van den Buuse, 2020; Pinto et al., 2017; Smoller et al., 2013) is aligned with recent theoretical frameworks such as the National Institute of Mental Health’s Research Domain Criteria (RDoC) framework for psychiatric disorders (Cuthbert and Insel, 2013), the Developmental Origins of Health and Disease model (Kofink et al., 2013; Lester et al., 2012; Van Den Bergh, 2011), and the study of endophenotypes in the move towards precision medicine (Hellhammer et al., 2018; Notaras and van den Buuse, 2020). Moreover, summarizing the methodological approaches used and the specific positions of CpG sites examined in DNAm studies (which compose the vast majority of epigenomic studies of BDNF) will likely be informative in guiding the design of future epigenomic studies of the association of BDNF with brain-related conditions.

Thus, our primary aim was to systematically review studies investigating *BDNF* epigenomic modifications in association with brain-related phenotypes in humans. We adopted a broad definition of brain-related phenotypes by aiming to include both brain-related “conditions” and brain-related “correlates”. Brain-related conditions included psychiatric disorders, neurological disorders, neurodevelopmental disorders, and other brain-related pathologies. Brain-related “correlates” included dependent variables closely related to brain function, such as cognitive processes and brain structure or function examined via neuroimaging, or independent variables with demonstrated effects on brain function, mainly trauma exposure and psychotropic medication treatment. We summarize the evidence relating to: (1) type of epigenomic modifications (e.g. DNAm, non-coding RNA, or histone modification); (2) brain-related phenotypes; (3) specific positions of CpG sites examined in DNAm studies; (4) genotype by DNAm results for DNAm studies; (5) methodological approaches (e.g., candidate vs. epigenome-wide association study [EWAS], tissue type, platform, adjustment for covariates, and correction for multiple comparisons); and (6) significance and direction of associations. Finally, we provide recommendations for future epigenomic studies based on the results of the present systematic review.

A particularly novel contribution of our review is the creation of an open access web-based application, the *BDNF* DNA Methylation Map available at https://lwheinsberg.shinyapps.io/BDNF_DNAmMap/, which allows users to interactively visualize the specific positions of *BDNF* DNAm CpG sites investigated across all studies for which relevant data were available, and customize a figure based on phenotype of interest, tissue type, statistical significance, and CpG position number. We anticipate that this application will be a valuable tool for the scientific community and encourage more innovative and dynamic approaches to interacting with epigenomic data in reporting scientific results.

## 2. Methods

This systematic review followed the Preferred Reporting Items for Systematic Reviews and Meta-Analyses (PRISMA) (Moher et al., 2009) and Meta-Analysis of Observational Studies in Epidemiology (MOOSE) (Stroup et al., 2000) publishing guidelines.

### 2.1. Literature search

PubMed (NLM), Embase (Elsevier), APA PsycInfo (Ovid), and Cochrane Protocols (Wiley) were searched; health sciences librarians with systematic review experience (AMK, HMV) assisted with the development of all searches. The completion date of the initial searches was 20 April 2020. A citation analysis of included studies from the original search indicated that only studies from PubMed (NLM) and Embase (Elsevier) were included, which were last updated 17 September 2021 and 27 September 2021 respectively. Concepts that made up the search were: (1) BDNF; (2) epigenomic modification (e.g. DNAm, non-coding RNA, or histone modification); and (3) brain-related conditions or correlates (e.g. central nervous system diseases, psychiatric disorders, trauma, cognition, etc.). A combination of MeSH terms and title, abstract, and keywords were used to develop the initial PubMed search which was checked against a known set of studies. The search was then adapted to search the other databases. EndNote (Clarivate) was used to sort and store citations; duplicates were removed using the processes described by Bramer et al and Otten et al (Bramer et al., 2016; Otten et al., 2019). We limited the search to studies on humans and published in English. The complete search strategy can be found in Appendix A.

### 2.2. Study selection and inclusion criteria

Studies were included if they described an epigenomic modification (DNAm, non-coding RNA, or histone modification) in association with the *BDNF* gene and a brain-related phenotype in humans. Additional criteria for inclusion were: (1) the presentation of original study data in a peer-reviewed journal article (i.e., dissertations, conference proceedings, commentaries, editorials, letters, books, book chapters, and reviews were excluded), and (2) the full-text article was available in English.

Results of our literature search were imported into Distiller SR (Evidence Partners) literature review software (https://www.evidencepartners.com/products/distillersr-systematic-review-software) for web-based independent review. Each title and abstract were screened by two of four reviewers (ATB, LWH, TSD, SB) who independently assigned the record a binary ‘‘Yes” (i.e., retain abstract and retrieve full-text article) or ‘‘No” (exclude from consideration). Distiller SR was also used to evaluate records selected for full-text article review after screening. Each full-text record was assessed by two reviewers (ATB, LWH, TSD, SB) who progressed through inclusion criteria until either all inclusion criteria were met, or the record was excluded for a particular reason (see Fig. 1). At both screening and full-text review, reviewers could only view records assigned to them and were not privy to decisions submitted by fellow reviewers. If reviewers were not in agreement, a final decision was reached in consensus and with a third reviewer (YPC) if needed.

**Figure 1.**
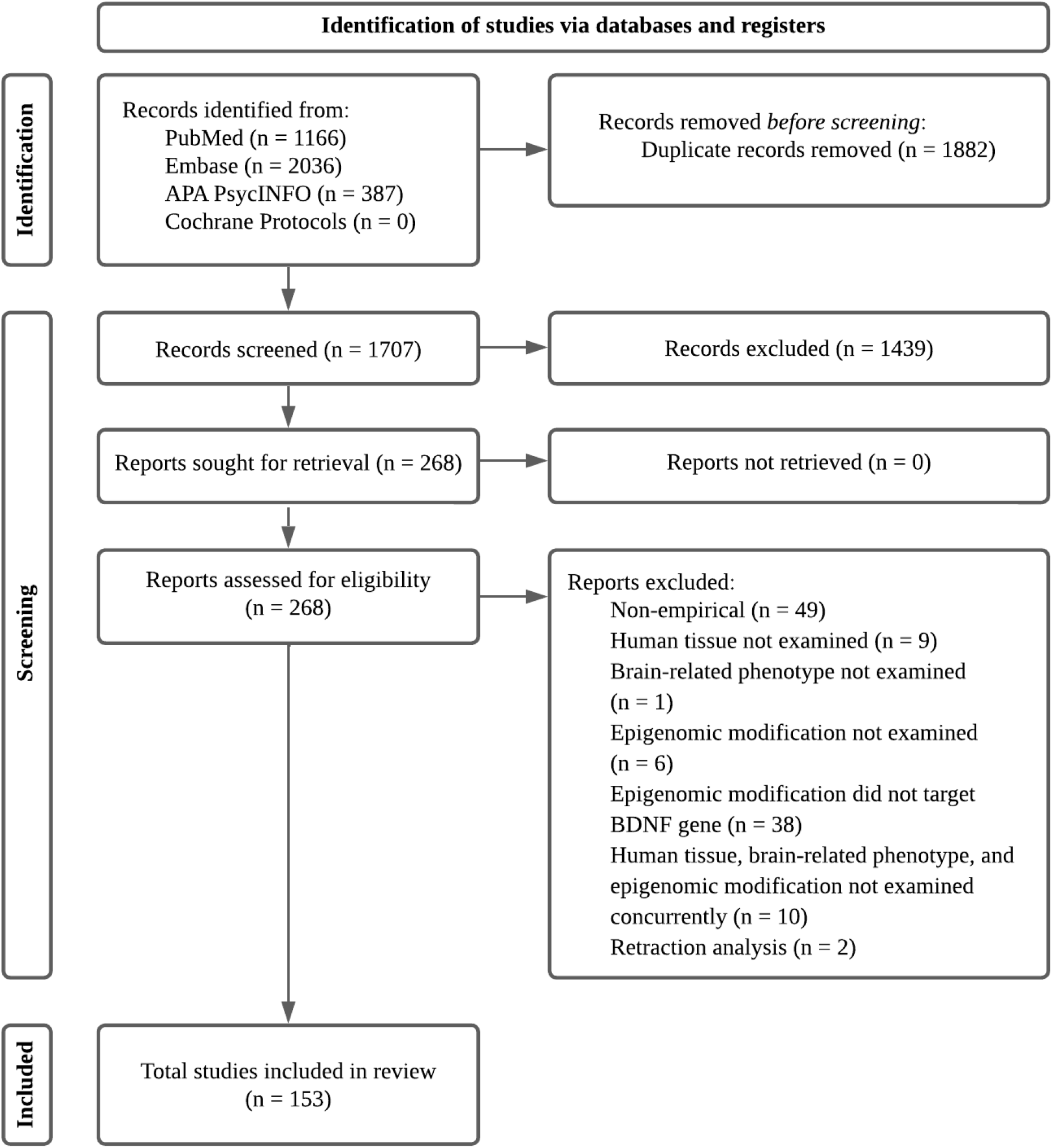
PRISMA flow chart displaying the screening process for studies included in this systematic review

Prior to data extraction, we searched for each study that was to be included in Retraction Watch (http://www.retractionwatch.com) and PubMed to identify retraction notices or notices of correction. Articles were then excluded if retracted or if significant concern about scientific misconduct was present based on other retractions from the author group.

### 2.3. Data extraction

Data from included studies were collected using predesigned data extraction forms in Distiller SR, according to the type of epigenomic modification (DNA methylation, histone modification, or non-coding RNA). Data extracted included type of epigenomic modification, participant characteristics (phenotype/groups, sample size, % male, age, country/race/ethnicity), timing of sample collection, tissue and cell type (or brain region for brain tissues), candidate gene vs. EWAS approach, platform, CpG sites(s), non-coding RNA(s), or histone(s) examined, covariates, whether genotype x DNAm was examined, main results, and threshold for statistical significance. Data extraction was independently performed by four authors (ATB, LWH, TSD, SB) after training using five articles and reaching at least 80% agreement per DistillerSR’s weighted overall kappa statistic. Any questions regarding data extraction were discussed and resolved as a group by consensus in consultation with the senior author (YPC). We did not pool results for meta-analysis given the high heterogeneity among studies regarding phenotype, CpG positions examined, and tissue type.

For each DNAm study, we extracted exact genome positions of examined CpG sites, or if not reported, information that could be used to identify them, including DNA sequences of CpG sites and their surrounding regions, genome position ranges of CpG sites, and primer sequences. Bioinformatic tools, including UCSC Genome Browser, NCBI nucleotide blast, and CLC Genomics Workbench were used to align reported sequences to the Genome Reference Consortium Human Build 38 (hg38). If insufficient information was provided in the article to identify exact positions of CpG sites examined, we emailed the corresponding author requesting this information. We subsequently coded each CpG site examined as significant or not significant in association with the phenotype of interest according to the authors’ specified threshold for significance (consistent with results reported in summary tables).

### 2.4. BDNF DNA Methylation Map

To interactively visualize the specific positions of *BDNF* DNAm CpG sites investigated across all studies for which these data were available, we developed the *BDNF* DNA Methylation Map, an open access web-based application available at https://lwheinsberg.shinyapps.io/BDNF_DNAmMap/. The *BDNF* DNA Methylation Map was developed in R version 4.1.2 (Team, 2021) using a suite of packages including shiny (Chang et al., 2015), ggplot2 (Wickham, 2016), and plotly (Sievert, 2020). The application runs on data synthesized through this systematic review, curated in long form. In brief, we created a to-scale depiction of the *BDNF* gene (i.e., base plot), including exons, promoter regions, and transcription start sites according to Pruunsild and colleagues (Pruunsild et al., 2007). Next, CpG sites and study x CpG x phenotype x tissue specific results were added to the figure, which was then made customizable via several user input filters (i.e., user-defined plot). Finally, we provided access to an interactive searchable database that contains all information synthesized in the plot (i.e., table). All figure data were mapped to human genome assembly 38 (hg38). For more complete details on the application configuration and development, please refer to the full source code and database publicly available via GitHub [https://github.com/lwheinsberg/BDNF_DNAmMap]. Additional details on the use and navigation of the *BDNF* DNA Methylation Map can be found in section 3.3.1.2.

### 2.5. Quality assessment

Although PRISMA and MOOSE guidelines require quality assessment of included studies, we did not perform a formal quality assessment due to the lack of specific guidelines for epigenomic studies (e.g., required sample size according to approach, platform coverage, performance of validation). However, we did extract data related to sample size, covariate adjustment, correction for multiple comparisons, and replication, and provide additional commentary on methodological considerations associated with study quality.

## 3. Results

### 3.1. Study inclusion

As shown in Figure 1, the search strategy identified a total of 3,589 records, of which 1882 were duplicates, resulting in 1707 unique records. Screening of titles and abstracts excluded 1439 records, resulting in 268 full-text reports sought for retrieval. An additional 113 reports were excluded after full-text review, resulting in a total of 153 studies included in the present review.

### 3.2. Study characteristics

The 153 studies included 112 studies of DNAm (Tables 2-14), 37 studies of non-coding RNA (Table 15), and 4 studies of histone modifications (Table 16) in the *BDNF* gene in association with brain-related phenotypes in humans. We categorized each study into broad brain-related phenotypes, shown in Table 1, to present results by phenotype in section 3.4 and in the *BDNF* DNA Methylation Map. Several studies included participants that could be assigned to multiple broad phenotypes (e.g. participants who both had major depressive disorder and were suicidal; participants with depression and stroke). In these instances, we assigned studies to only the one broad phenotype that appeared to best align with the primary objectives of the study. However, readers interested in multiple phenotypes can select to view results for multiple phenotypes concurrently or by specific reference identifier (RefID) in the online *BDNF* DNA Methylation Map.

**Table 1.** Epigenomic modifications and Broad Brain-Related Phenotypes of Included Studies.

|  |  |  |  |
| --- | --- | --- | --- |
| Broad Phenotype | DNA<sup>m</sup> (n=112) | Non-coding RNA (n=37) | Histone modification (n=4) |
| Psychopathology: Depressive Disorders | 23 | 7 | 0 |
| Psychopathology: Suicide & Suicidal Ideation | 7 | 0 | 0 |
| Psychopathology: Bipolar Disorder | 7 | 1 | 0 |
| Psychopathology: Schizophrenia | 10 | 3 | 0 |
| Psychopathology: Substance Abuse | 7 | 1 | 0 |
| Psychopathology: Trauma/Stress Exposure and Trauma-Related Disorders | 17 | 0 | 1 |
| Psychopathology: Other | 8 | 0 | 0 |
| Neurological Injury or Disorder: Dementia and Mild Cognitive Impairment | 12 | 12 | 2 |
| Neurological Injury or Disorder: Stroke | 5 | 4 | 0 |
| Neurological Injury or Disorder: Other | 7 | 7 | 1 |
| Chronic Pain/Fatigue | 3 | 0 | 0 |
| Pediatric Cognition and Behavior | 3 | 1 | 0 |
| Aging | 3 | 0 | 0 |
| Other | 0 | 1 | 0 |

### 3.3. Study results

For ease of readability and referencing, studies are henceforth referred to in the text by their RefID in square brackets consistent with Tables 2-16 (e.g. [1]). Additionally, hovering over shapes in the *BDNF* DNA Methylation Map will reveal pop-up boxes containing information that corresponds to Tables 2-14.

#### 3.3.1. Transdiagnostic overview of DNAm studies

##### 3.3.1.1. Summary tables

Details of each DNAm study are summarized by broad phenotype in Tables 2-14. The results column summarizes DNAm results using study-specific CpG site labels (e.g. CpG1). The directions of statistically significant associations (e.g., hypo- or hypermethylation; positive or negative association) are specified. Non-significant associations are reported as “NS”. In the summary tables, results are summarized after covariate adjustment when applicable (see “Covariates” column for details). Likewise, statistical significance reported in the summary tables is consistent with the study-specific approach and authors’ definition of “significance”— regardless of adjustment for multiple comparisons. Specifically, results of studies that conducted multiple statistical tests and either (1) interpreted any nominal finding of *p* < .05 as significant, or (2) interpreted only findings of *p* < a Bonferonni-adjusted significance threshold as significant, are both reported as statistically significant in the summary tables and in the *BDNF* DNA Methylation Map. Therefore, the results are not always directly comparable between studies and should be interpreted considering differences in study-specific statistical approach. We provide more details regarding heterogeneity in statistical approaches below.

##### 3.3.1.2. BDNF DNA Methylation Map

We were able to identify exact positions of examined CpG sites for 94/112 DNAm studies (84%); therefore, only these studies are represented in the *BDNF* DNA Methylation Map, as indicated in the final column of each summary table. Studies were not included in the *BDNF* DNA Methylation Map if they did not provide sufficient information to identify exact positions of CpG sites examined and this information could not be obtained from corresponding authors. A static version of the *BDNF* DNA Methylation Map is provided in Figure 2.

**Figure 2.**
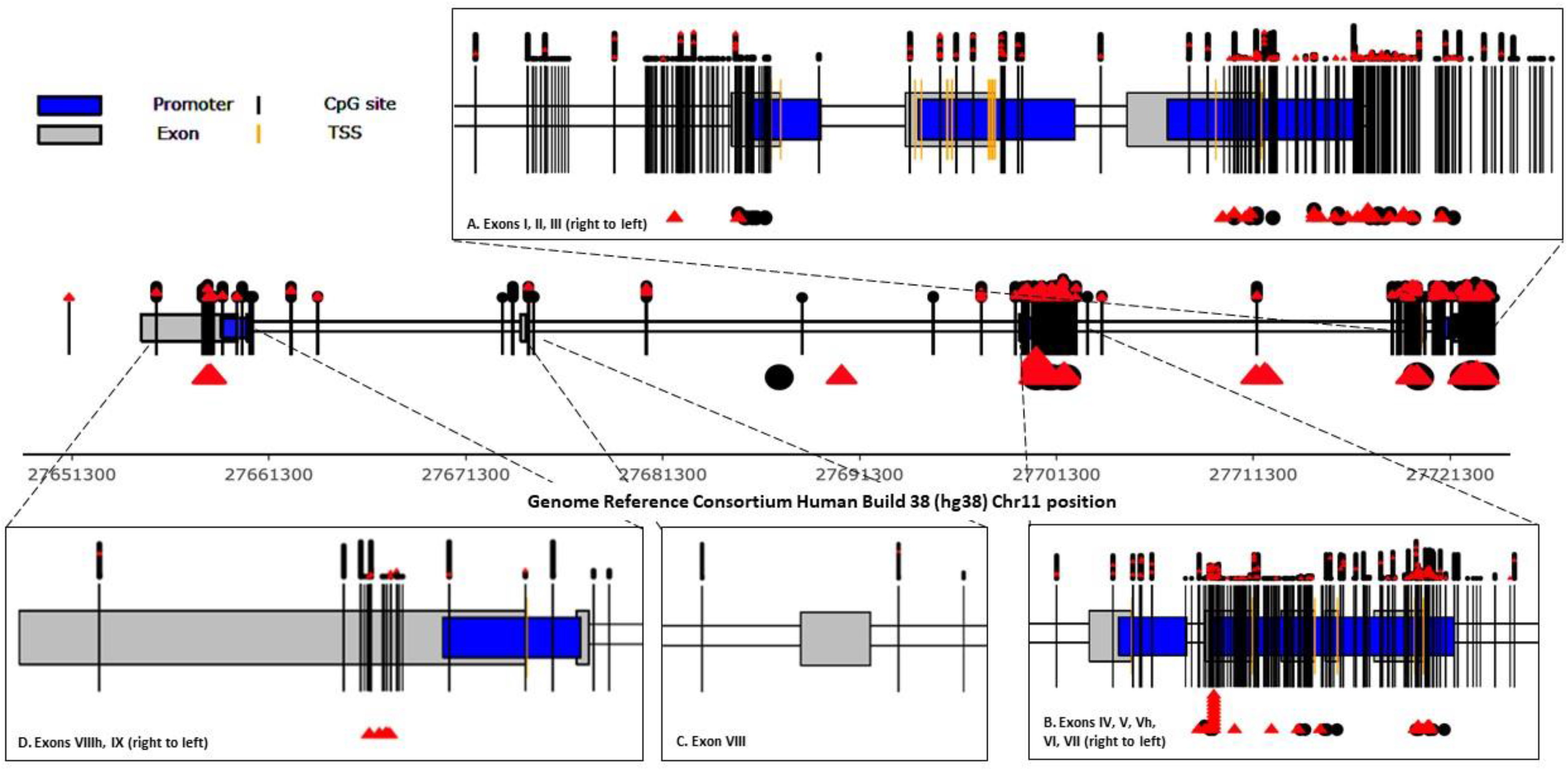
Static view of the *BDNF* DNA Methylation Map, an interactive web application containing a to-scale visualization of the *BDNF* gene and the positions of all CpG sites examined in association with brain-related phenotypes in humans, available at https://lwheinsberg.shinyapps.io/BDNF_DNAmMap/. The present figure shows the positions of each CpG site (black vertical lines) for which this data was available. CpG sites are mapped to the Genome Reference Consortium Human Build 38 (hg38) and shown in relation to *BDNF* exons (gray boxes), promoter regions (blue boxes), and transcription start sites (gold vertical lines) according to Pruunsild and colleagues. In this static view, each study that examined a particular CpG site is represented by a black circle (non-significant in association with brain-related phenotype) or red triangle (significant in association with brain-related phenotype) above each CpG site (vertical black line). The taller the tower of shapes above each CpG site, the higher the number of studies examined that particular CpG site. Larger black circles (non-significant) and red triangles (significant) below the CpG sites represent each study that examined the mean DNAm across several CpG sites, with the circle or triangle positioned in the center horizontally of the CpG sites for which the mean DNAm was calculated. The taller the tower of shapes below each CpG site, the higher the number of studies examined the mean DNAm centered at that particular CpG site. This figure shows data extracted across all studies, including all broad brain-related phenotypes and all tissue types. Panels A through D provide “zoomed in” views of the exon regions. The online version of the *BDNF* DNA Methylation Map can be used to zoom in to view additional detail of any region of the figure; customize the figure by broad phenotype, tissue, and statistical significance; filter by RefID or CpG position number; color the study-specific shapes by either statistical significance, phenotype, or tissue; hover over CpG sites or shapes for exact position and/or additional study details; and click on each shape to navigate to the website containing the abstract and/or full text of the study.

As described in section 2.4, the *BDNF* DNA Methylation Map, available at https://lwheinsberg.shinyapps.io/BDNF_DNAmMap/, is an open access, interactive web application designed to synthesize the DNAm-specific results of this study. In the to-scale interactive depiction of the *BDNF* gene, CpG sites are shown in relation to *BDNF* exons (gray boxes), promoter regions (blue boxes), and transcription start sites (gold vertical lines). Users can zoom in to view specific regions of the figure in more detail or hover over shapes to view pop-up boxes containing information that corresponds to Tables 2-14. Users can also customize the figure based on their interests and preferences with filters available for RefID, CpG position, broad phenotype, tissue type, statistical significance, and “color by” option (statistical significance, phenotype, or tissue). For additional details on use and navigation of the *BDNF* DNA Methylation Map, refer to more detailed instructions in the Figure 2 caption and the online application itself.

##### 3.3.1.3. Methodological approaches

The total sample size across all DNAm studies ranged from 11 to 3,300 participants with a mean of 283 and median of 131. Thirty-six percent of studies (n = 40) had a total sample size < 100 participants. Sixteen percent of studies (n = 18) had a total sample size > 500 participants.

The reporting of the timing of biosample collection and tissue type was extremely variable across studies. Because DNAm is dynamic, the timing of biosample collection relative to condition onset or duration, exposure (e.g., trauma), or intervention (e.g., psychotropic medication initiation) is a key consideration in interpreting associated DNAm results. Despite its importance, data on timing of biosample collection were not reported in approximately 40% of pertinent studies. Likewise, because DNAm patterns are tissue-specific and reflective of the local environment of each cell type, and examination of brain tissue is not feasible in many brain-related phenotypes, determining which types of tissue biosamples serve as effective proxies for the brain environment is an important methodological consideration. DNA was extracted from blood in the vast majority of studies (n = 89; 79%), followed by brain (n = 10; 9%), saliva (n = 6; 5%), buccal cells (n = 6; 5%), umbilical cord blood (n = 4; 4%), placenta (n = 2; 2%), muscle (n = 1; 0.9%), spermatozoa (n = 1; 0.9%), and cerebrospinal fluid (n = 1; 0.9%). More than half of the studies in which DNA was extracted from blood further specified whole blood, with some specifying cell types (e.g., leukocytes, mononuclear cells). Most studies that extracted DNA from brain tissue specified the brain region from which tissue was collected. Among studies in which DNA was extracted from mixed cell types from any tissue, only approximately 15% adjusted for cell type heterogeneity.

Regarding candidate gene vs. EWAS approaches, 100 studies (89%) focused on *BDNF* with a candidate gene approach, six (5%) used an EWAS approach only, and six (5%) reported results of both candidate and EWAS approaches. EWASs that, by definition, examined the *BDNF* gene but did not call out *BDNF* in titles, abstracts, keywords, or MeSH terms would not have been identified by our literature search strategy and, therefore, were not included in the present review.

Quantification of DNAm in the 12 EWASs was performed using the Illumina Infinium HumanMethylation 450 BeadChip in all but two studies, one of which used the Illumina Infinium MethylationEPIC BeadChip and the other used an earlier version microarray approach with less genome coverage. Quantification of DNAm in the candidate gene studies was performed using pyrosequencing most often (n = 62; 58.5%), followed by Illumina Infinium HumanMethylation 450 BeadChip (n = 15; 14.2%), MassARRAY (n=10; 9.4%), quantitative polymerase chain reaction (n = 9; 8.5%), targeted bisulfite sequencing (n = 8; 7.5%), Illumina Infinium MethylationEPIC BeadChip (n = 4; 3.8%), matrix-assisted laser desorption/ionization-time-of-flight (n = 2, 1.9%), methylation sensitive high resolution melting (n = 1, 0.9%), and Illumina Infinium 27K human DNA methylation BeadChip (n = 1. 0.9%).

Statistical approaches also reflected substantial heterogeneity, notably, specification of covariate adjustment and multiple comparison correction. In addition to adjustment for cell type heterogeneity mentioned above, the number of covariates for which adjustments were made ranged from 0 (n = 42; 38%) up to 17 in a single study. The most examined covariates were age, sex, education, body mass index, smoking status, antidepressant use, race/ethnicity, history of depression, income/socioeconomic status, and disability status. Fewer than half of the studies (n = 47; 42%) applied any method to correct for multiple comparisons.

Remarkably, only two studies tested for replication of their *BDNF* DNAm findings in independent samples. Study [23], however, tested for replication at the gene level rather than at the CpG site level, with no comparison of the results for the three overlapping CpG sites between the discovery and replication samples. Investigators in study [98] tested for replication of their epigenome-wide significant *BDNF* CpG site, Illumina probe cg13974632, in association with seizures in adolescence. This site was not significant in the replication sample but a meta-analysis of both samples revealed evidence of DNAm differences after Bonferroni correction.

##### 3.3.1.4. Patterns of DNAm results

Eighty-five percent (n = 95) of included studies reported at least one statistically significant (per author definition) association between *BDNF* DNAm and a brain-related phenotype. The studies reflected a mix of those reporting higher vs. lower DNAm in patients relative to healthy controls or in association with disease/disordered phenotypes, with a slightly higher number of results reporting higher DNAm associated with disease/disorder phenotypes.

As can be observed from the *BDNF* DNA Methylation Map and Figure 2, studies of *BDNF* DNAm in association with brain related-phenotypes across all tissue types have examined CpG sites (black vertical lines) in and around all 11 *BDNF* exons (grey boxes) and their associated promoter regions (blue boxes). All additional CpG sites that have been examined outside of these exon or promoter regions are Illumina probes included on the 450K or EPIC beadchips (tallest towers of shapes above CpG sites representing the number of studies that examined each Illumina probe). The *BDNF* DNA Methylation Map and Figure 2 also make clear that, aside from the Illumina probes, the most frequently examined CpG sites (as represented by taller towers of shapes above each CpG site) were positioned around exon/promoter I (see Figure 2, panel A) and exons/promoters IV and VI (see Figure 2, panel B). A smaller cluster of studies examined CpG sites in exon IX (Figure 2, panel D).

In the static view presented in Figure 2 (and when the “color by” option for the *BDNF* DNA Methylation Map is set to ‘statistical significance’), the color of the shapes above and below the CpG sites represents the statistical significance (per study author definition) of each CpG site (above), or calculation of mean DNAm across several CpG sites in a region (e.g., region-based mean DNAm; below), in association with brain-related phenotypes within each study. The *BDNF* DNA Methylation Map can be examined for clusters of red towers to locate the positions of “hot spots” wherein investigated CpG sites were most often significantly associated with brain-related phenotypes, customizable to the user’s interests and specifications. As can be appreciated in the online *BDNF* DNA Methylation Map and Figure 2, the most prominent “hot spots” across all brain-related phenotypes and tissue types are in the following hg38 approximate regions: (1) chr11:27,721,905-27,723,016 in exon/promoter I; (2) chr11:27,701,549-27,701,744 in exon/promoter IV; and (3) chr11:27,700,516-27,700,187 in exon/promoter VI.

Interestingly, when the *BDNF* DNA Methylation Map is filtered by tissue type to view the tissues much less frequently examined than blood, a clear “hot spot” remains around hg38 chr11:27,701,578-27,701,743 in exon IV, which is repeated across brain, saliva, placenta, and buccal tissues. Coloring the *BDNF* DNA Methylation Map by tissue type or broad phenotype (via the “color by” user input option) does not reveal any clearly appreciable regions of particular significance for a certain tissue type or broad phenotype. Notable patterns of results within each broad phenotype are discussed in section 3.3.2.

Of the 12 EWASs, four reported significant probes in *BDNF* after epigenome-wide multiple comparison correction. Phenotypes were lifetime exposure to childhood domestic violence [65], neighborhood socioeconomic disadvantage or neighborhood social environment [69], seizures in adolescence [98], and fibromyalgia [105]. Study [65] reports on DNA extracted from saliva while the rest reported on DNA extracted from blood. None of the 11 significant probes across the four studies overlapped. While they tended to cluster in exon and promoter regions, no enrichment was clearly apparent near any specific exon.

##### 3.3.1.5. DNAm by genotype results

Approximately one third (n = 35, 31%) of studies examined DNAm in association with *BDNF* genotype. Of these, 24 studies (69%) reported significant DNAm by genotype results. The most examined SNP by far was rs6265, which was included in over 90% of such studies. Results of rs6265 by DNAm analyses were mixed regarding both significance and the direction of associations, with about half of studies reporting a significant association, and of these, about half reporting higher, and half reporting lower, DNAm in carriers of the risk (Met) allele (associated with reduced activity-dependent secretion of *BDNF*). Most of these studies were in blood, with no discernable pattern of DNAm x rs6265 results by tissue type. Other SNPs examined in association with DNAm included: rs908867, rs7103411, rs1491850, rs11030101, rs28722151, rs11030094, rs11602246, rs925946, rs4923463, rs2030324, rs12273363, rs2049048, rs1491851, rs962369, rs1519480, rs7127507rs10767664, rs12273539, rs11030096, rs11030102, rs11030108, rs10501087, rs11030104, and rs988748. All these SNPs were reported as non-significant in association with DNAm except for three of four studies of rs908867 [4, 14, 36] and one of two studies of rs2030324 [87]. See summary tables 2-14 for these results.

#### 3.3.2. DNAm studies by broad brain-related phenotype

##### 3.3.2.1. Psychopathology: depressive disorders

The most examined phenotype among *BDNF* DNAm studies was depressive disorders, with a total of 23 studies (Table 2). Most studies examined major depressive disorder (MDD) in adults, often compared to a healthy control group. Additional phenotypes examined in individuals with depression included cognitive function [4, 5] and brain structure via neuroimaging [6, 7]. Several studies examined the effect of medication [8, 10, 11, 12] or electroconvulsive therapy [9]. Several studies examined depressive symptoms within cohorts of individuals with other common characteristics or conditions, including older adults [13, 14], individuals with breast cancer [15], individuals with acute coronary syndrome, Japanese workers [18], and monozygotic twin pairs with or without a history of trauma [23]. Two studies examined the effect of prenatal depression in mothers on DNAm in infants [20, 21]. And finally, one study examined healthy adults with high sociotropy, a personality vulnerability factor for depression [22]. Two studies [16 and 17] appear to report results from the same cohort of individuals.

Most studies reported on DNA extracted from blood, two from saliva, two from buccal swabs, and one from neonatal umbilical cord blood. Notably, a candidate approach was used in all the studies. Thirteen studies included adjustments for covariates and nine carried out corrections for multiple comparisons. The depressive disorders phenotype contained one of the two studies in the present review that included testing for replication [23].

Given that depressive disorders were the most studied broad phenotype, patterns of examined and significant CpG sites for which positions could be identified are generally consistent with the transdiagnostic description in section 3.3.1.4, with notable “hot spots” of significant CpG sites in exon/promoters I and VI. The exon/promoter VI “hot spot” comprises studies published almost exclusively by the same group of authors [13, 15-17], who studied the identical collection of CpG sites in association with depressive symptoms in several cohorts of individuals with acute coronary syndrome, older individuals, and individuals with breast cancer, respectively. Significant individual CpG sites were also located slightly downstream of exon III, in exon/promoter IV, in exon Vh, and exon IX. Studies in which region-based mean DNAm was examined reported significant associations across many CpGs in multiple exons/promoters [9-10].

**Table 2.**
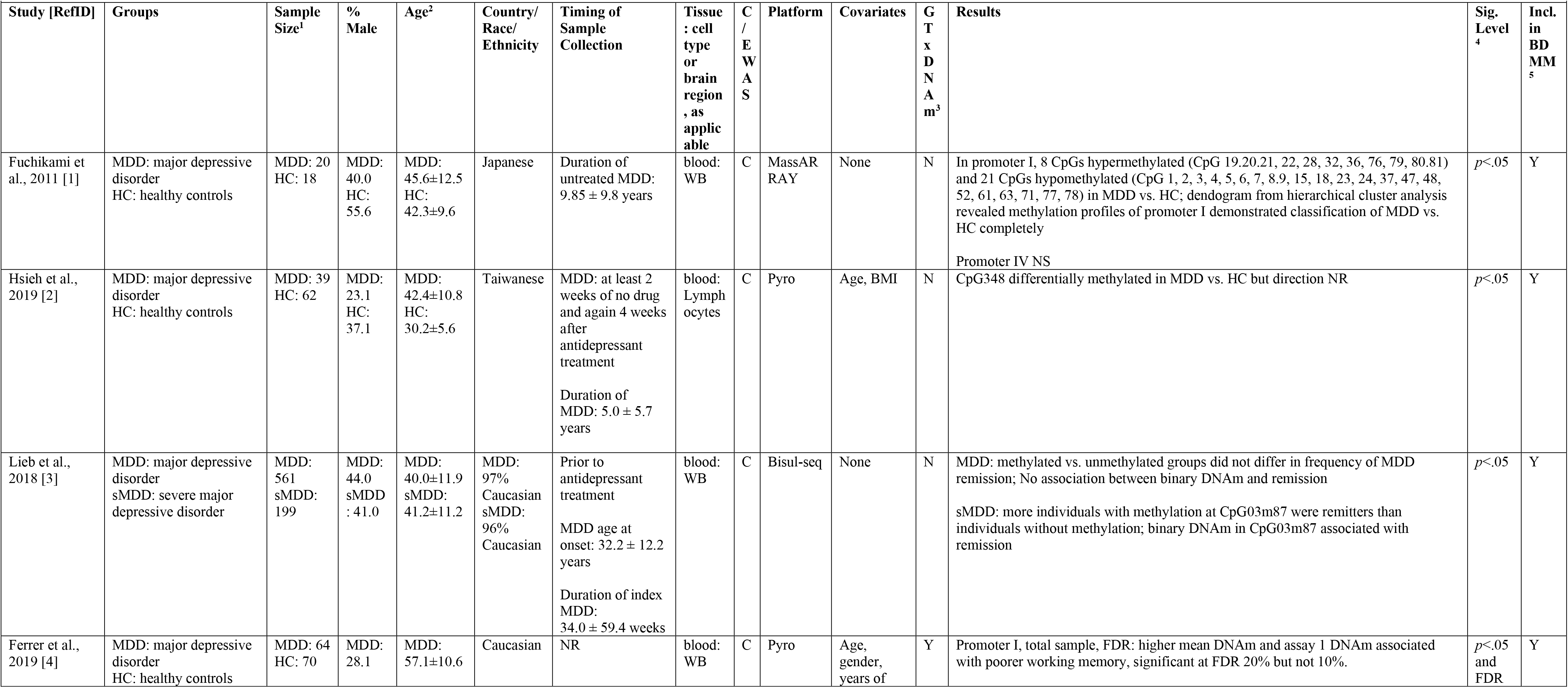
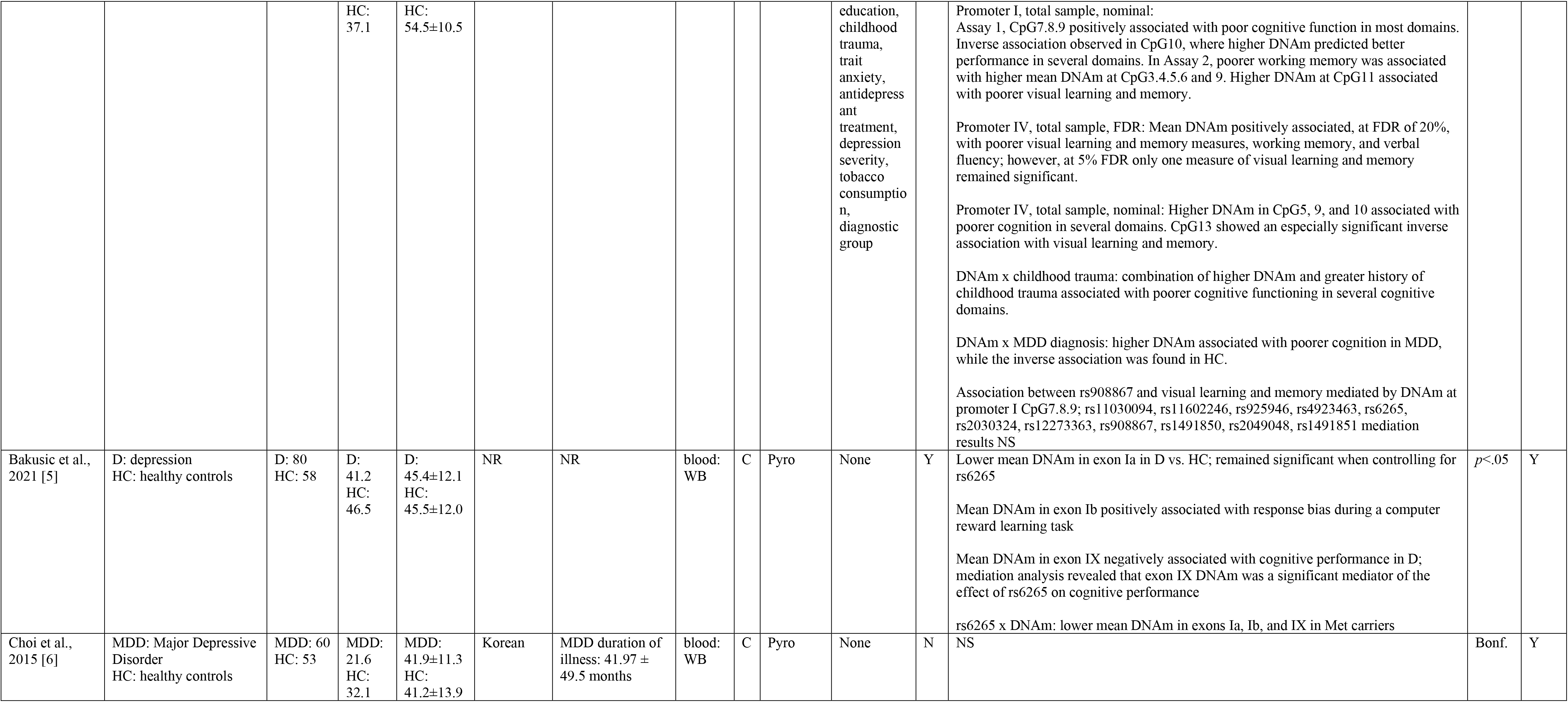
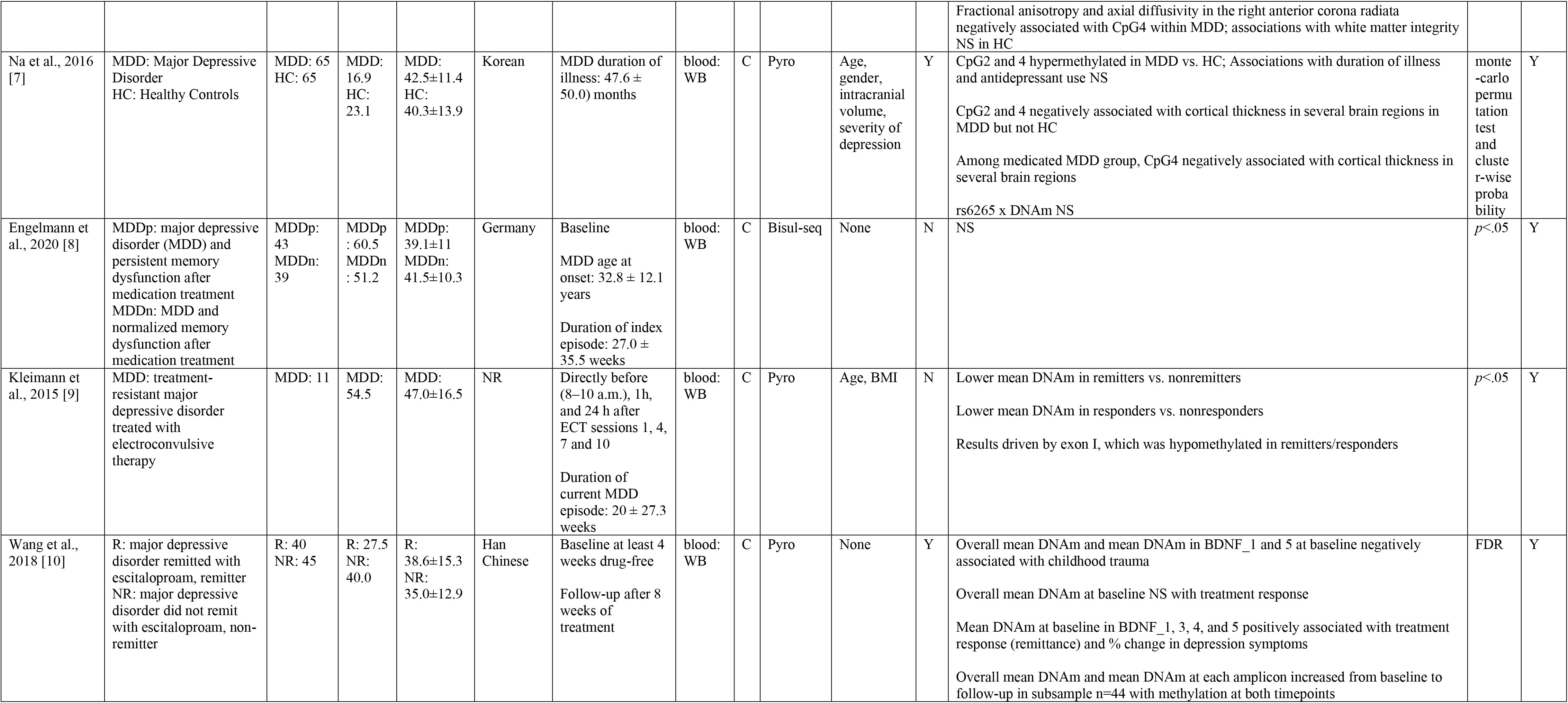
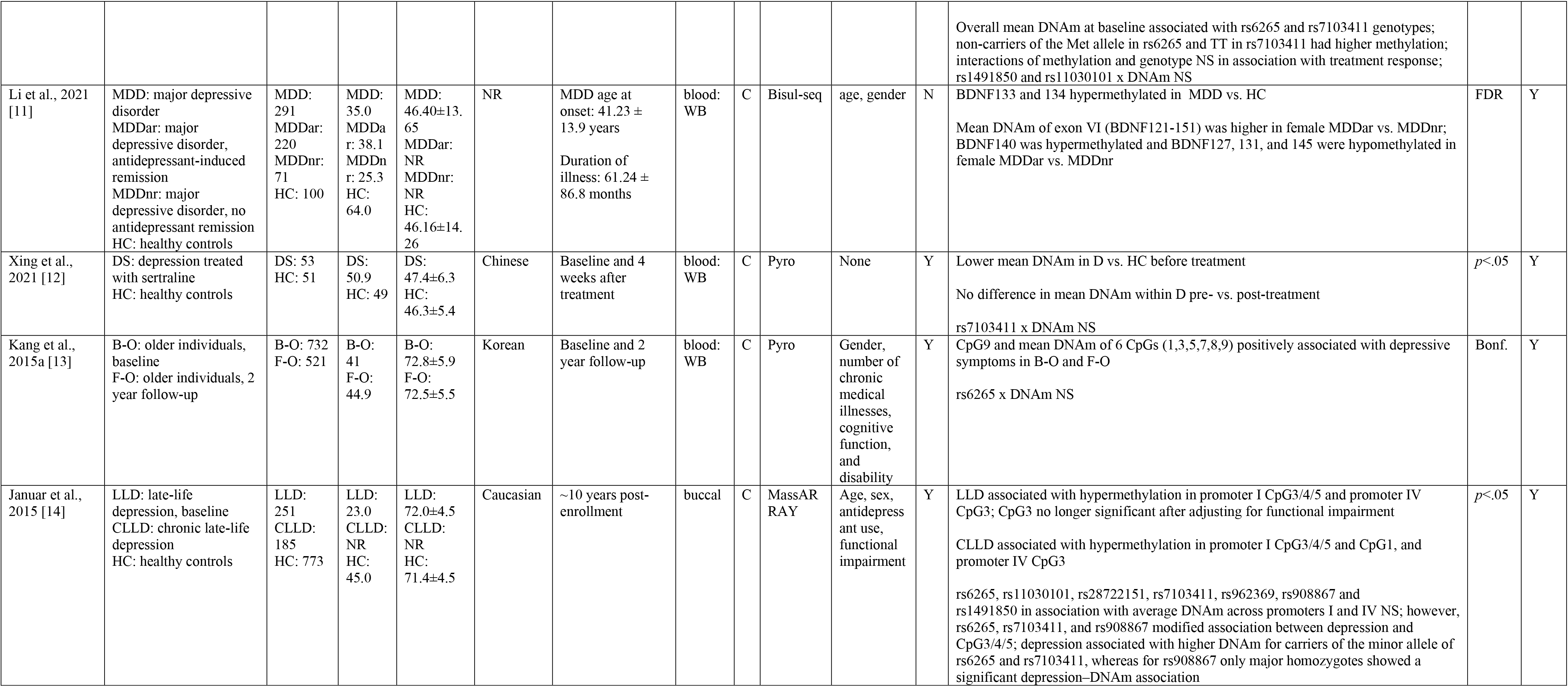
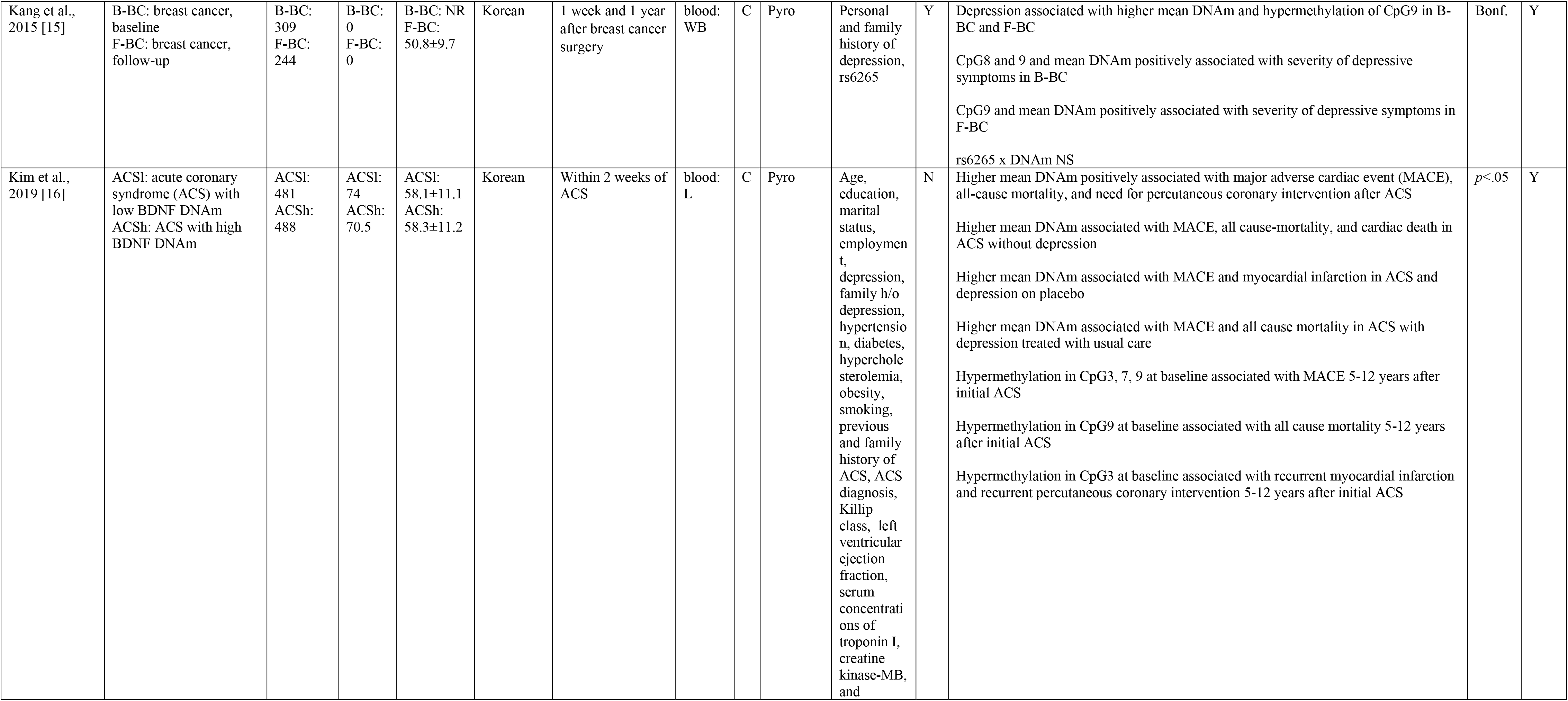
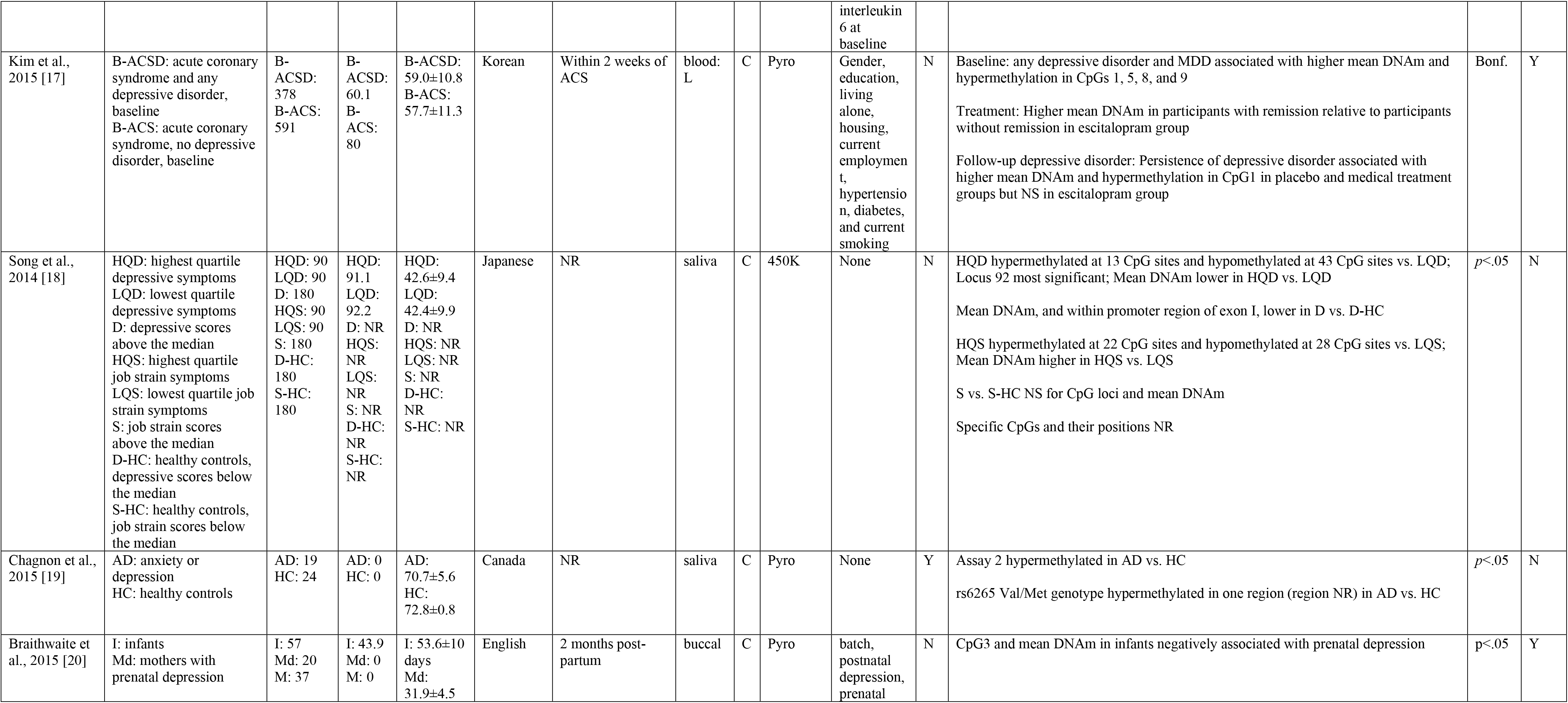
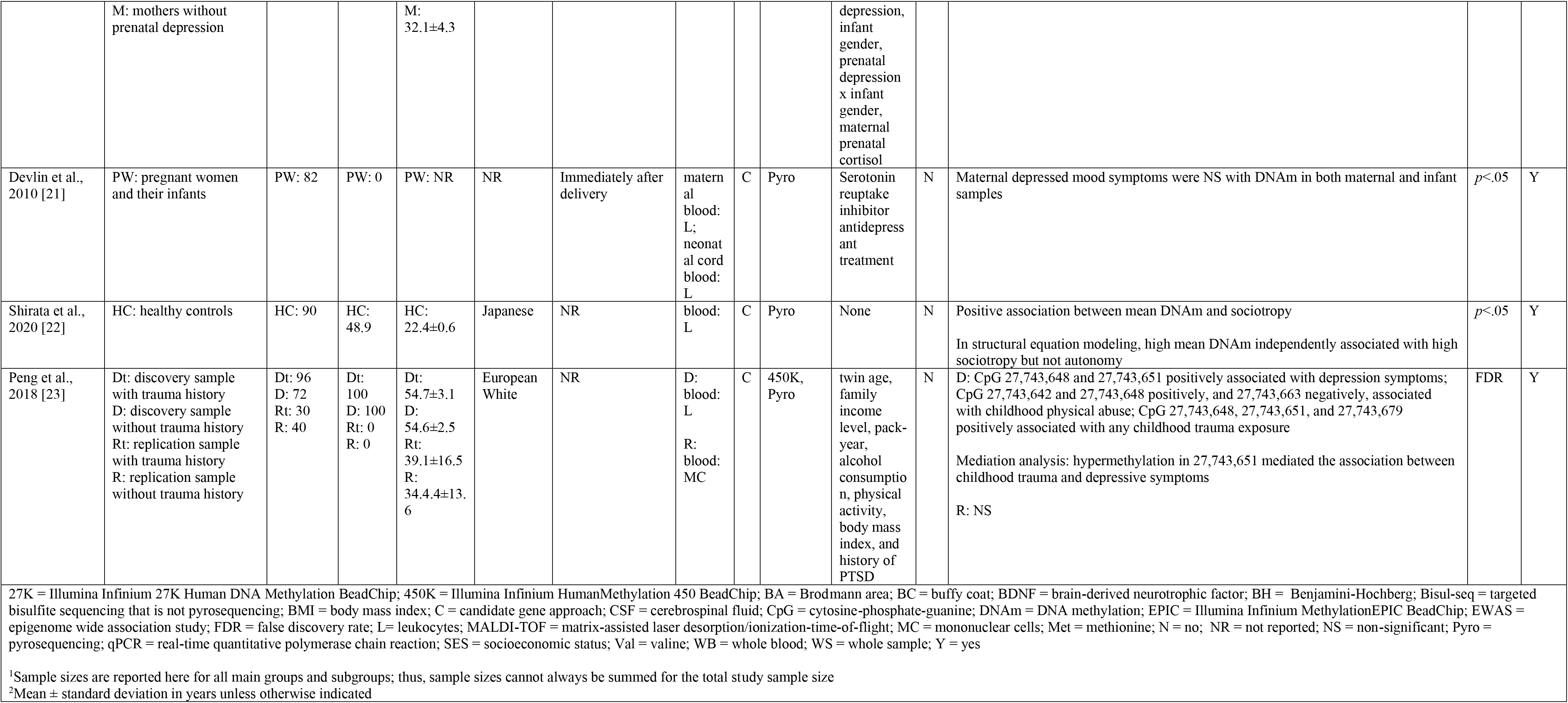
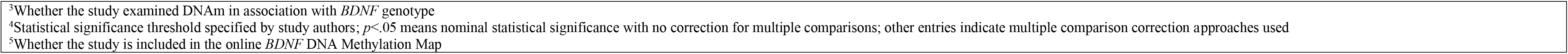
DNAm Studies. Psychopathology: Depressive Disorders.

##### 3.3.2.2. Psychopathology: suicide and suicidal ideation

Seven studies examined *BDNF* DNAm in association with suicide and suicidal ideation (Table 3) in individuals with major depressive disorder [24, 25], serious suicidal ideation [27, 29, 30], acute coronary syndrome [26], and breast cancer [28]. In five of these, DNA was extracted from blood and in two from brain. Six studies used a candidate approach and one used an EWAS approach [29]. Five studies adjusted for covariates and four studies corrected for multiple comparisons. Of studies for which positions could be identified, examined CpG sites were located across all exon/promoter regions with relative enrichment in exons/promoters I, IV, and VI. Significant individual CpG sites and region-based mean DNAm were located in exons/promoters I, IV, and VI.

**Table 3.**
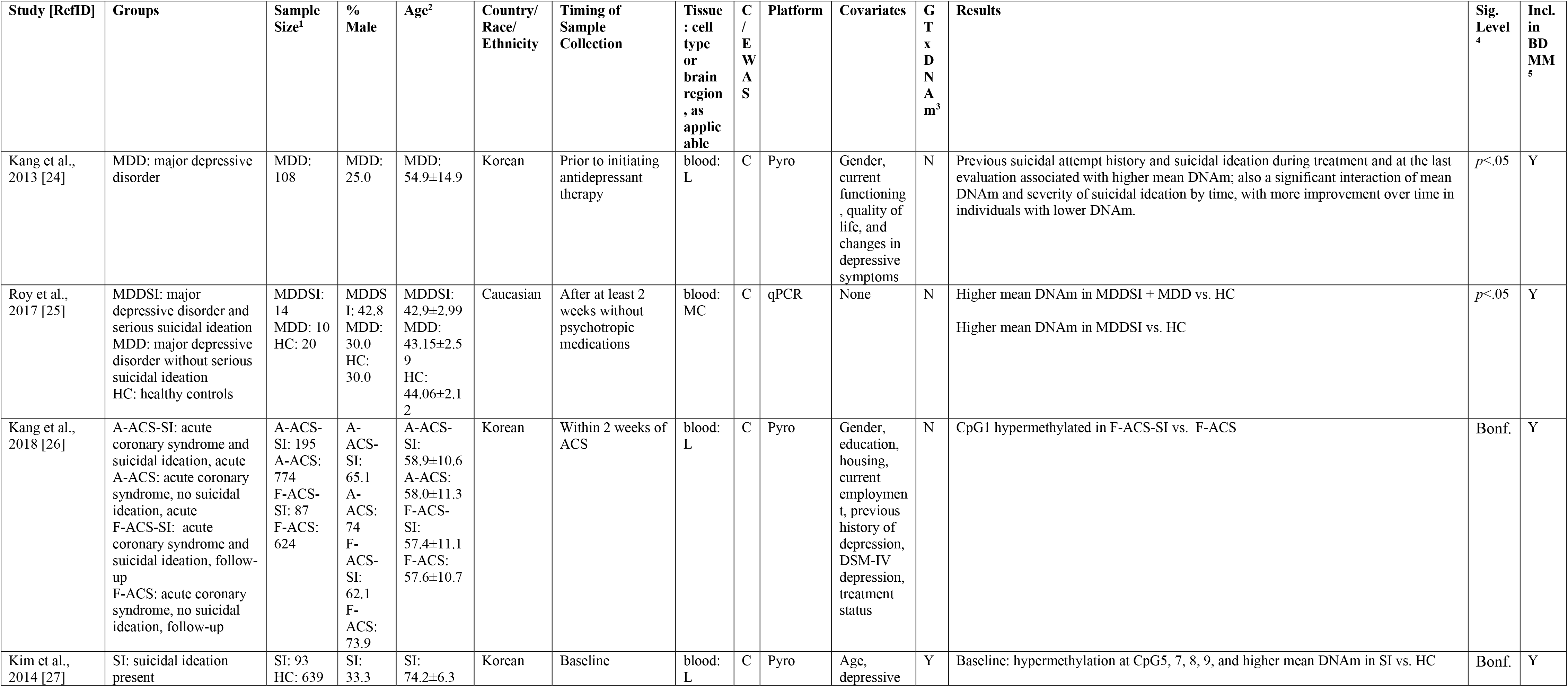
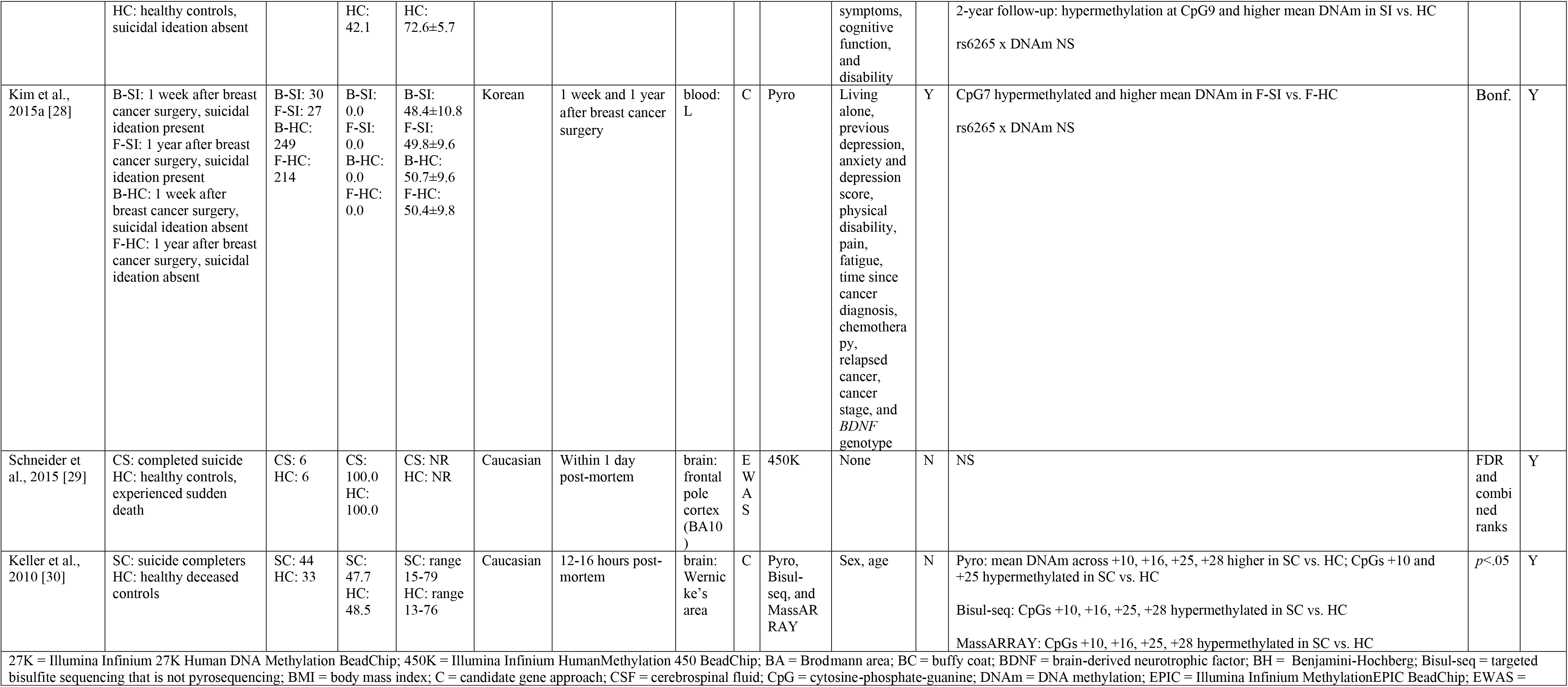
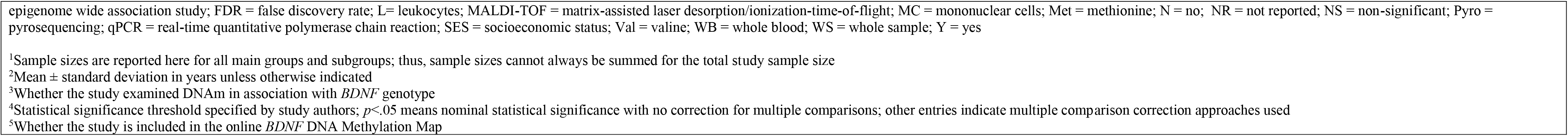
DNAm Studies. Psychopathology: Suicide & Suicidal Ideation.

##### 3.3.2.3. Psychopathology: bipolar disorder

Seven studies examined *BDNF* DNAm in association with bipolar disorder (Table 4) and in response to drug therapies [36, 34]. Several studies included multiple mood disorders, including major depressive disorder (or unipolar depression), bipolar type I, and bipolar type II [36, 35, 34]. In six, DNA was extracted from blood and in one from saliva. All studies used a candidate gene approach. Three studies adjusted for covariates and four studies corrected for multiple comparisons. Of studies for which positions could be identified, examined CpG sites were located only in the promoter of exon I, exon IV, exon/promoter Vh, and exon IX. Significant individual CpG sites and region-based mean DNAm were located in exon/promoter I, exon/promoter Vh, and exon IX.

**Table 4.**
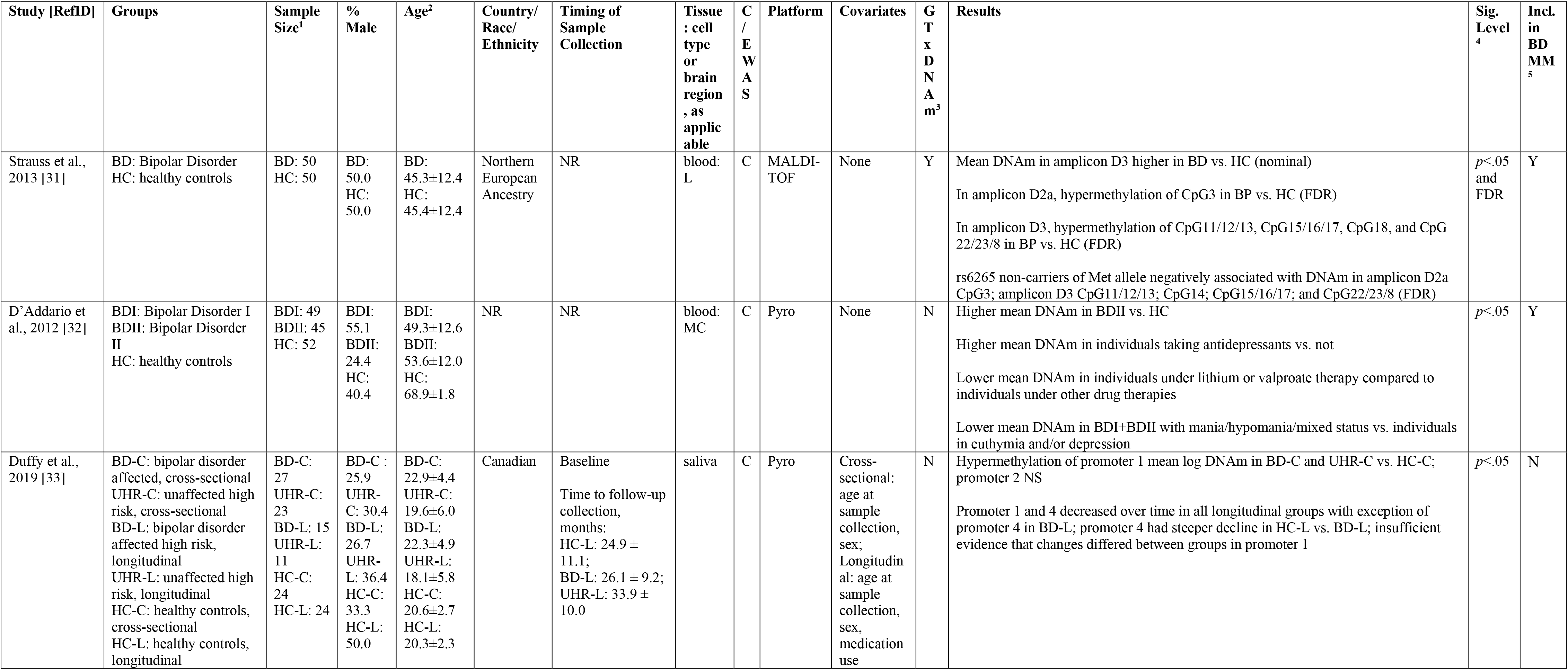
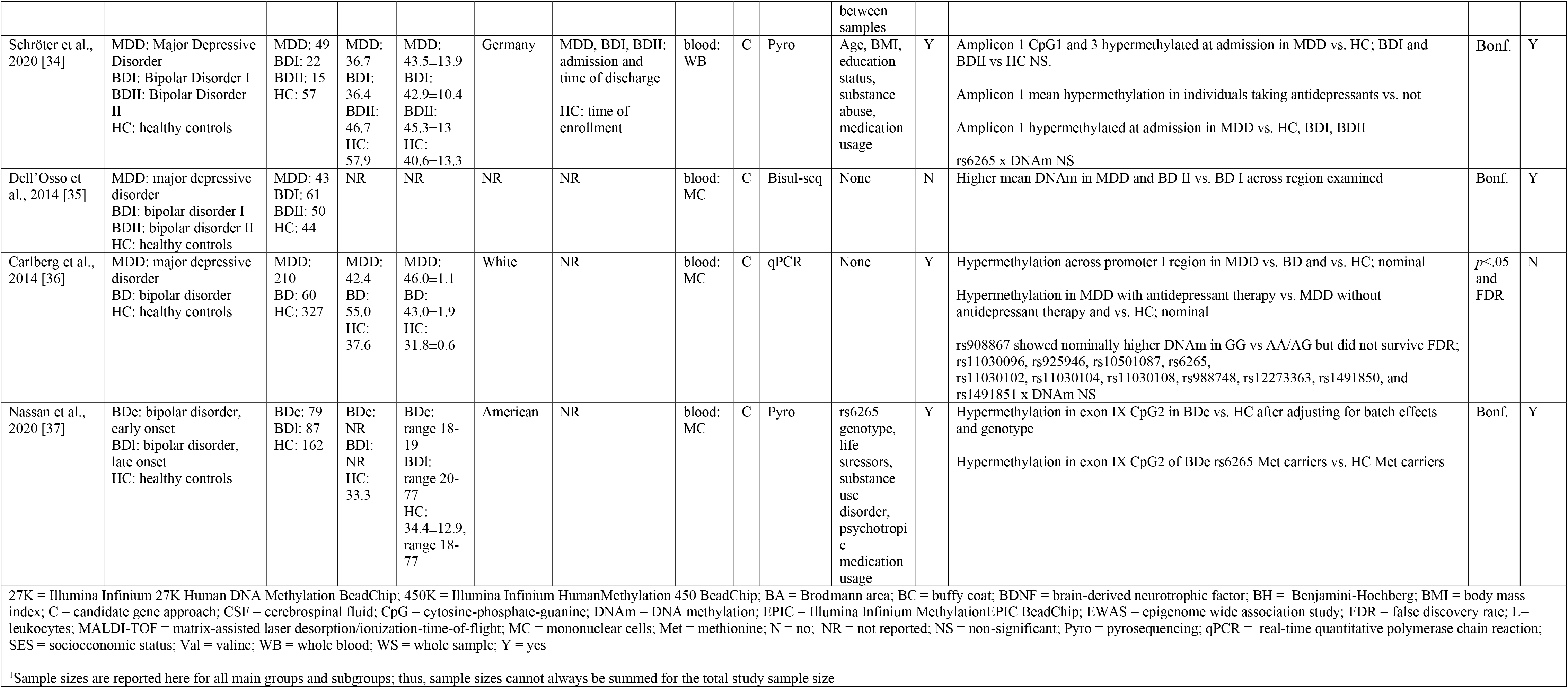
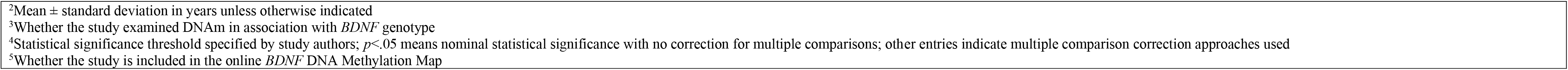
DNAm Studies. Psychopathology: Bipolar Disorder.

##### 3.3.2.4. Psychopathology: schizophrenia

Ten studies examined *BDNF* DNAm in association with schizophrenia (Table 5). In six studies DNA was extracted from blood, in one from buccal tissue, in three from brain, and in one from spermatozoa. Timing of tissue collection relative to disease onset was reported in five studies. All studies used a candidate approach and one also used an EWAS approach [39]. Four studies adjusted for covariates and three studies corrected for multiple comparisons. Of studies for which positions could be identified, examined CpG sites were located in exon/promoter I, exon III, exon/promoter IV, exon/promoter V, exon/promoter Vh, and exon IX. Significant individual CpG sites and region-based mean DNAm were located in exon/promoter I, exon III, and exon/promoter IV.

**Table 5.**
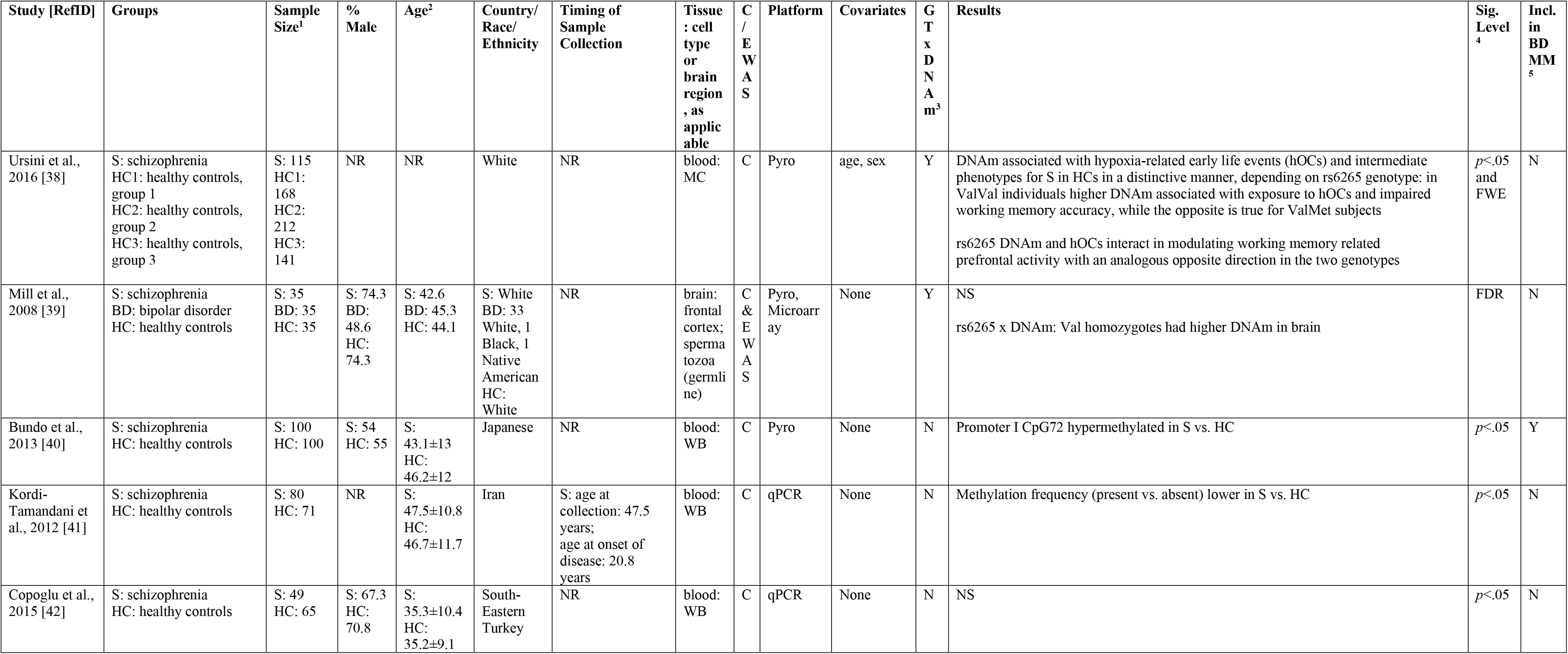
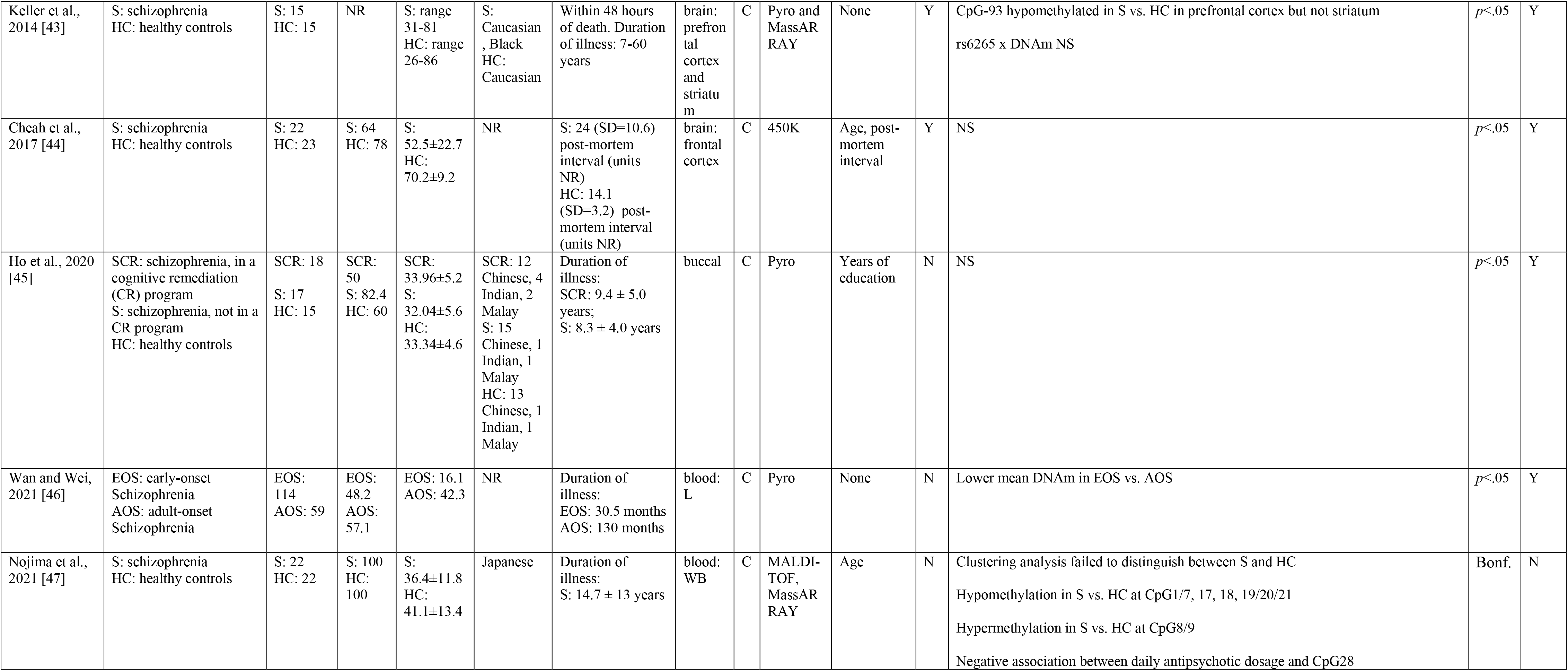
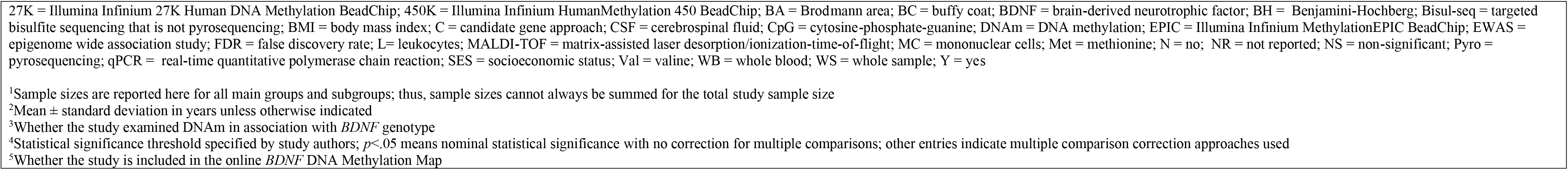
DNAm Studies. Psychopathology: Schizophrenia.

##### 3.3.2.5. Psychopathology: substance abuse

Seven studies examined *BDNF* DNAm in association with substance abuse, including substance abuse related outcomes and response to drug treatment in individuals with dependences to alcohol or drugs (Table 6). In all studies, DNA was extracted from blood and a candidate gene approach was used. Four studies adjusted for covariates and two corrected for multiple comparisons. Of studies for which positions could be identified, examined CpG sites were located only in exon/promoter I, exon/promoter III and a region downstream of exon III, exon/promoter IV, and exon/promoter Vh. There was a notably high proportion of significant CpG sites in exon/promoter I reported in two studies [50,54] and additional significant CpG sites in the promoter of exon IV.

**Table 6.**
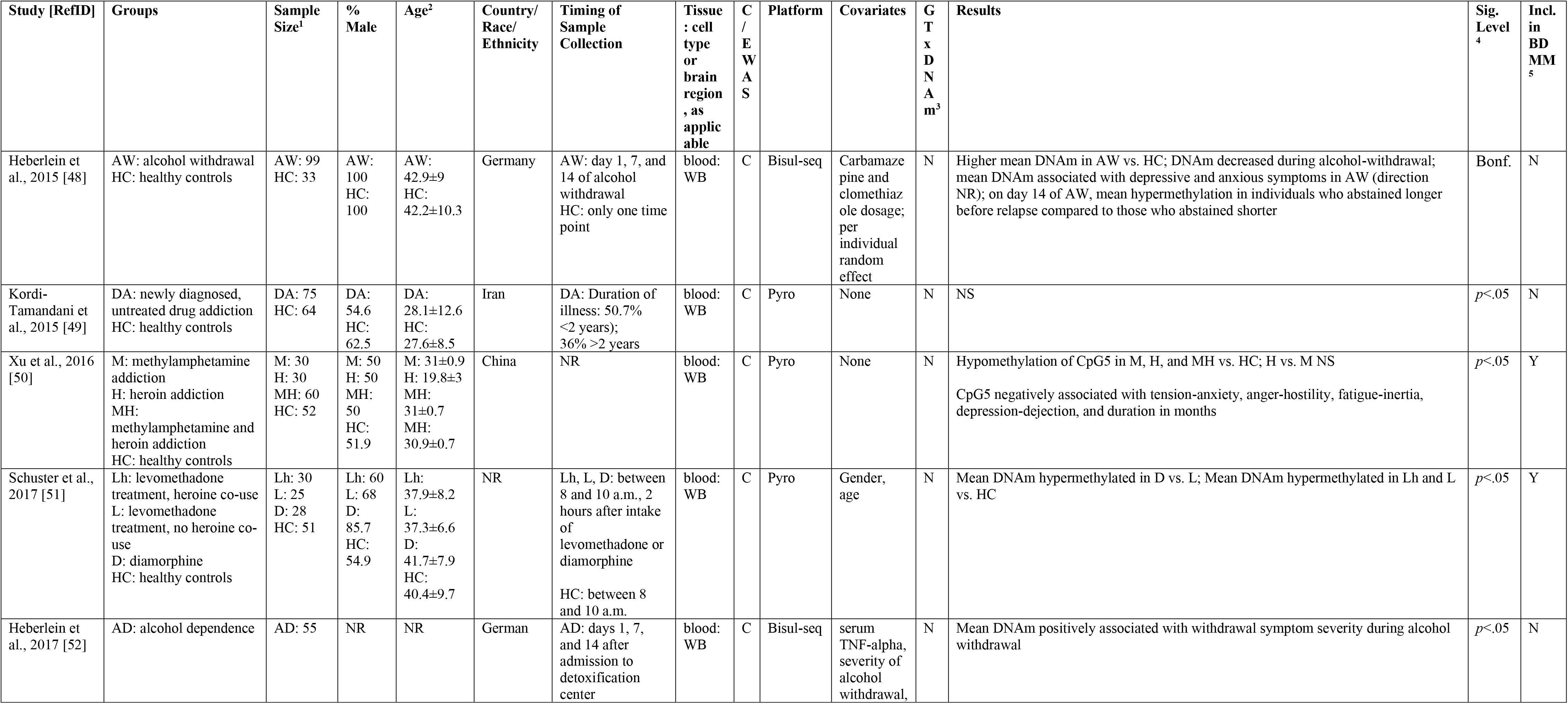
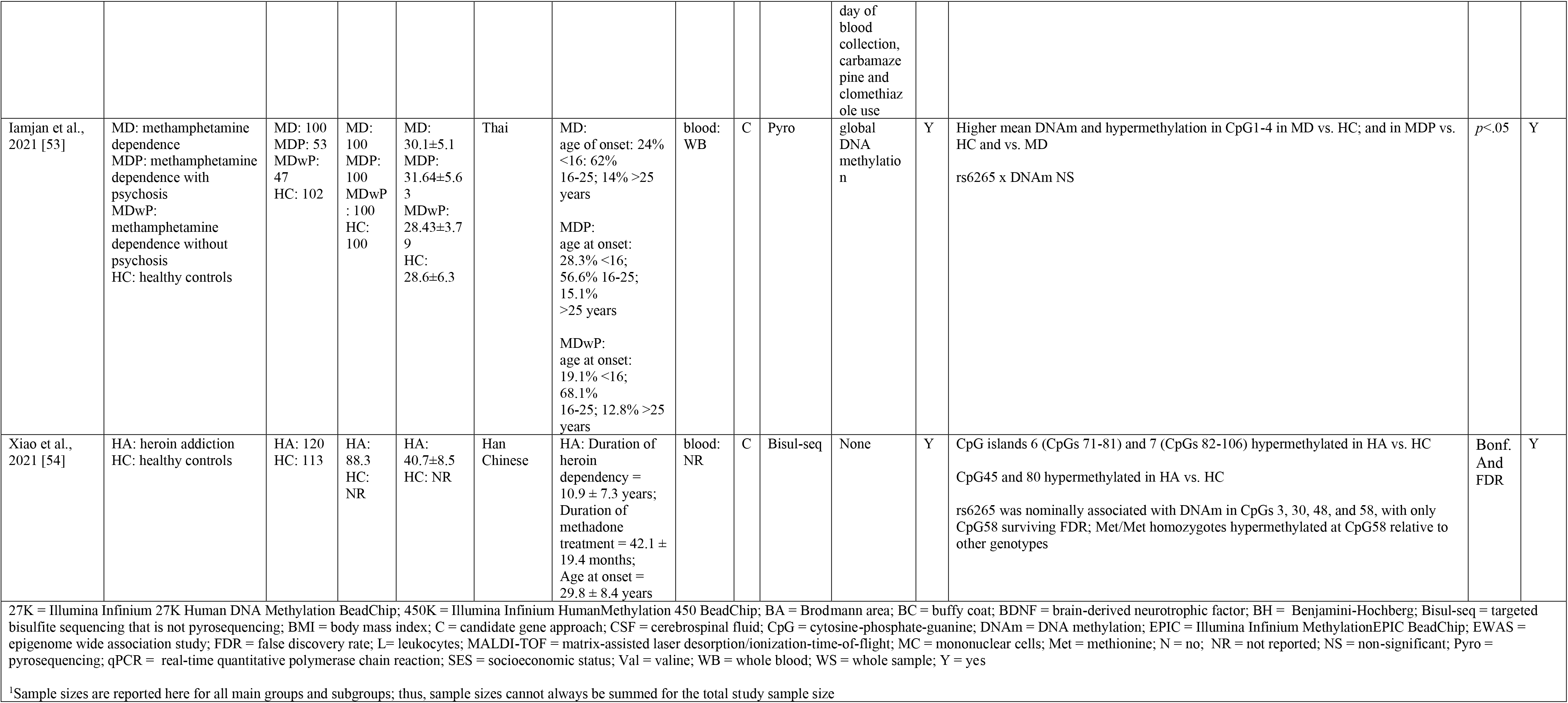
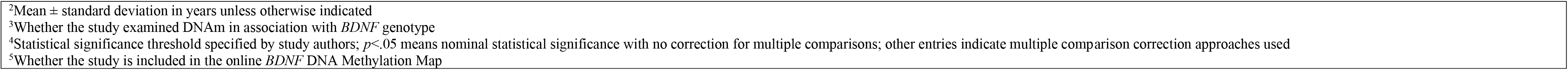
DNAm Studies. Psychopathology: Substance abuse.

##### 3.3.2.6. Psychopathology: trauma/stress exposure and trauma-related disorders

Seventeen studies examined *BDNF* DNAm in association with trauma/stress exposure and/or trauma-related disorders (Table 7). Of these, seven studies [55, 56 57, 58, 59, 60, 64] examined adults with post-traumatic stress disorder (PTSD), one of which was in the context of a randomized controlled trial [58], and one of which focused on mothers [64]. Several additional studies focused on mothers and/or their children, including a study of perceived discrimination in women during pregnancy and post-partum [61], a study of newborns of mothers with histories of child abuse or interpersonal or non-interpersonal trauma [63], and a study of Congolese mothers and infants exposed to chronic stress or war trauma [62]. Four studies focused on trauma or stress during childhood, including studies of depression in maltreated children [67], polyvictimization during childhood and/or adolescence [66], exposure to childhood domestic violence examined across three generations [65], and adults reporting high vs. low childhood maternal care [68]. The final three studies examined neighborhood-level socioeconomic disadvantage and social environment in a large atherosclerosis cohort [69], adults with bulimia nervosa with or without histories of physical abuse, sexual abuse, or bipolar disorder [70], and healthy adults before and after an acute psychosocial stressor [71].

In most of these studies, DNA was extracted from blood while in two it was extracted from saliva. Two of the five studies that extracted DNA from umbilical cord blood [62, 63], and one of the two studies that extracted from placenta [62], were in this phenotype category. Five of the 12 EWASs included in the present review were in this phenotype [65, 66, 58, 69, 67]; two of which [65 and 69] reported differentially methylated regions in *BDNF* in association with lifetime exposure to childhood domestic violence [65] and neighborhood socioeconomic disadvantage or neighborhood social environment [69]. The studies in this phenotype were among the most rigorous and well-controlled, with thirteen studies including covariates, six of which adjusted for cell type heterogeneity [57, 66, 58, 63, 69, 71], and ten studies correcting for multiple comparisons.

Given that this was the second most studied broad phenotype identified, patterns of examined and significant CpG sites for which positions could be identified are generally consistent with the transdiagnostic description in section 3.3.1.4, with notable “hot spots” of significant CpG sites in exon/promoters I, II, and IV. Significant CpG sites were also found in several other regions throughout the gene (see online *BDNF* DNA Methylation Map for details).

**Table 7.** DNAm Studies. Psychopathology: Trauma/Stress Exposure and Trauma-related Disorders.

|  |  |  |  |  |  |  |  |  |  |  |  |  |  |  |
| --- | --- | --- | --- | --- | --- | --- | --- | --- | --- | --- | --- | --- | --- | --- |
| Study [RefID] | Groups | Sample Size<sup>1</sup> | % Male | Age<sup>2</sup> | Country/<br>Race/<br>Ethnicity | Timing of<br>Sample<br>Collection | Tissue<br>: cell<br>type<br>or<br>brain<br>region<br>, as<br>applic<br>able | C<br>/<br>E<br>W<br>A<br>S | Platform | Covariates | G<br>T<br>x<br>D<br>N<br>A<br>m<sup>3</sup> | Results | Sig.<br>Level<br><sup>4</sup> | Incl.<br>in<br>BD<br>MM<br><sup>5</sup> |
| Kim et al.,<br>2017 [55] | PTSD: post-traumatic stress disorder<br>HC: healthy controls | PTSD: 126<br>HC: 124 | PTSD: 100<br>HC: 100 | PTSD: 63.2±3.5<br>HC: 62.9±4.4 | Korean | NR | blood: L | C | Pyro | Age, education, marital status, SES, combat exposure, harmful alcohol drinking | N | Higher mean DNAm in PTSD vs. HC; NS mediating effect of DNAm in the path from trauma exposure to PTSD status | $p<.05$ | Y |
| Guo et al.,<br>2018 [56] | PTSD: post-traumatic stress disorder<br>PTSDh: post-traumatic stress disorder, high trauma<br>PTSDl: post-traumatic stress disorder, low trauma<br>HC: healthy controls | PTSD: 322<br>PTSDh: 145<br>PTSDl: 177<br>HC: 215 | PTSD: 38.8<br>PTSDh : 42.9<br>PTSDl : 35.6<br>HC: 44.2 | PTSD: 39.3±7.2<br>PTSDh: 39.8±7.1<br>PTSDl: 38.8±7.2<br>HC: 38.7±7 | Han Chinese | NR | blood: WB | C | qPCR | None | N | CpG1-7, 9, and 12-18 were nominally hypomethylated in PTSD vs. HC<br>CpG1 and 18 were positively associated with aggressive violent trauma after multiple correction<br>CpG1, 6, 7, 10-12 were positively associated, and CpG9 negatively associated with traffic accident trauma (nominal); only CpG7 survived multiple correction | $p<.05$ and nonspecific “multiple correction” | Y |
| Voisey et al.,<br>2019 [57] | PTSD: post-traumatic stress disorder<br>HC: healthy controls | PTSD: 48<br>HC: 48 | PTSD: 100<br>HC: 100 | PTSD: 67.9±0.6<br>HC: 69.4±0.6 | NR | NR | blood: WB | C | EPIC | Age, BMI, Cell counts, alcohol history, psychotropic medication | Y | cg01546433, cg24650785, and cg002298481 hypomethylated in PTSD vs. HC and positively associated with increased PTSD symptom severity<br>cg04481212, cg01546433, and cg00298481 negatively associated with exercise<br>cg01546433 and cg24650785 correctly classified PTSD vs. HC after adjusting for covariates<br>rs6265 x DNAm NS | BH | Y |
| Yang et al.,<br>2021 [58] | PTSDph: post-traumatic stress disorder receiving psychotherapy and hydrocortisone treatment | PTSDph: 25<br>PTSDpp: 26 | PTSDph: 88<br>PTSDpp: 92 | PTSDph: 36.0±8.7<br>PTSDpp: 34.8±8.0 | PTSDph:44%<br>HispanicPTSDpp:38 | Baseline, 3, and 6 months after intervention | blood: WB | EWAS | 450K | Age, granulocyte percentage at time | N | NS | $p<.01$ | Y |
|  | PTSDpp: post-traumatic stress disorder receiving psychotherapy and placebo |  |  |  | .5% Hispanic |  |  |  |  | point 3, treatment types, gender |  |  |  |  |
| Guo et al., 2021 [59] | PTSD: post-traumatic stress disorder<br>HC: healthy controls | PTSD: 164<br>HC: 141 | PTSD: 51.8<br>HC: 51.1 | PTSD: 38.0±6.9<br>HC: 37.0±6.7 | Li and Han Chinese | NR | blood: WB | C | Methylation-Sensitive High-Resolution Melting | ethnicity, serum BDNF concentrations | N | DNAm positively associated with PTSD | $p<.05$ | N |
| Hossack et al., 2020 [60] | PTSD: post-traumatic stress disorder<br>HC: healthy controls | PTSD: 70<br>HC: 100 | PTSD: 75.7<br>HC: 87 | PTSD: 38.6<br>HC: 35.1 | PTSD: 30% White, 30% Black, 24.3% Hispanic, 5.7% Asian, 10% Other<br>HC: 50% White, 22% Black, 20% Hispanic, 4 % Asian, 4% Other | NR | blood: WB | C | Pyro | antidepressant use | N | Hypomethylation in BDNF_08 Pos2-4 and BDNF_10 Pos1-2 and 4-5 in PTSD vs. HC | $p<.05$ | Y |
| Santos et al., 2018 [61] | pregnant women | 147 | 0 | 27.6±6.4 | Latina | Gestational age = 28.8 ±1.3 weeks | blood: BC | C | Pyro | Maternal age, marital status, presence of a partner, education, household income, ethnicity, years living in the U.S, nativity<br>(US or<br>non-US-born),<br>infant sex,<br>depressive<br>symptoms | N | Pregnancy: CpG 6 and 7 negatively associated with perceived discrimination<br>Post-partum: CpG 5, 6, and 7 negatively associated with perceived discrimination | FDR and BH | Y |
| Kertes et al.,<br>2017 [62] | M: mothers<br>I: infants | M: 24<br>I: 24 | M: 0<br>I: 54 | M:<br>26.9±5.6<br>I:<br>newborns | Congolese | Shortly after<br>delivery | Blood:<br>WB;<br>cord<br>blood:<br>WB;<br>placent<br>a | C | 450K | infant sex<br>for cord<br>blood and<br>placenta<br>models | N | Maternal blood: cg2429411, cg16257091, cg04106006, cg06684850, cg12448003, cg25412831, cg26949694, cg11806762 negatively, and cg05189570 positively, associated with war trauma, all of which survived FDR; cg04106006 and cg06684850 were also nominally negatively associated with chronic stress<br>Placenta: cg10558494, cg15313332, cg25962210 positively associated with war trauma after FDR; cg25962210 and cg26949694 also nominally positively associated with chronic stress; cg16257091, cg26840770, and cg27193031 nominally positively associated with war trauma; cg09492354 nominally negatively associated with both stressor types.<br>Cord blood: cg17413943 negatively associated with war trauma and cg16257091 positively associated with chronic stress after FDR; cg01225698 and cg09492354 nominally positively associated, and cg07704699 nominally negatively associated, with unspecified stressor | *p*<.05<br>and<br>FDR | Y |
| Pilkay et al.,<br>2020 [63] | ITP: newborns of<br>mothers with history of<br>interpersonal trauma<br>NITP: newborns of<br>mothers with history of<br>non-interpersonal trauma<br>CAb: newborns of<br>mothers with history of<br>child abuse<br>NCAb: newborns of<br>mothers, no history of<br>child abuse | WS: 216;<br>group-<br>specific<br>sample<br>sizes NR | ITP: 0<br>NITP:<br>0<br>CAb: 0<br>NCAb:<br>0 | NR | 55%<br>African<br>American,<br>45%<br>White | Shortly after<br>delivery | cord<br>blood:<br>WB | C | 27K | Mediation<br>analyses:<br>newborn<br>sex,<br>newborn<br>race, cell<br>compositio<br>n | N | Lifetime fear associated with higher mean DNAm in male newborns | *p*<.05 | Y |
| Moser et al.,<br>2015 [64] | Mothers with and<br>without interpersonal<br>violence-related PTSD | 54 | 0 | NR | NR | NR | saliva | C | pyro | None | N | Mean DNAm and CpG1, 3, and 4 positively associated with severity of maternal anxiety<br>Maternal exposure to domestic violence as a child positively associated with CpG3<br>Mean DNAm positively associated with maternal brain activity in a cluster comprising the left ventromedial prefrontal cortex, orbitofrontal cortex, and the ventral anterior cingulate cortex, as well as with clusters in the right ventromedial prefrontal cortex and posterior cingulate cortex, when mothers watched own and unfamiliar children during scenes of separation vs. play | Bonf. | Y |
| Serpeloni et al., 2020 [65] | CDVa: childhood domestic violence, adolescents<br>NCa: no childhood domestic violence, adolescents<br>CDVm: childhood domestic violence, mothers<br>NCm: no childhood domestic violence, mothers<br>CDVg: childhood domestic violence, grandmothers<br>NCg: no childhood domestic violence, grandmothers | CDVa: 34<br>NCa: 90<br>CDVm: 38<br>NCm: 87<br>CDVg: 24<br>NCg: 102 | CDVa: 52.9<br>NCa: 44.5<br>CDVm: 0<br>NCm: 0<br>CDVg: 0<br>NCg: 0 | CDVa: 14.6±2.7<br>NCa: 13.4±2.4<br>CDVm: 37.3±6.4<br>NCm: 39.2±6.2<br>CDVg: 63.2±7.8<br>NCg: 65±8.3 | Brazilian | NR | saliva | E<br>W<br>A<br>S | 450K | Exposure to other traumatic events in all models; sex and age in adolescent model; age in grandmother model | N | Hypomethylation of cg06260077 with increased lifetime exposure to CDV in adolescents; cg06260077 association with exposure to CDV was also observed in the maternal but not the grandmaternal generation | FDR | Y |
| Marzi et al., 2018 [66] | childhood and adolescent polyvictimization | 1658 | 49 | 18 | England and Wales | Age 18 years | blood: BC | C<br>&<br>E<br>W<br>A<br>S | 450K | sex, methylation-array control-probe principal components indexing technical variation, cell-type proportion estimates, smoking status | N | EWAS: No BDNF probes were associated with adolescent polyvictimization, childhood victimization, cumulative victimization, or retrospective reports of childhood victimization after array-wide Bonferroni correction<br>C: cg20954537 was hypermethylated in association with retrospective report of childhood victimization after gene-wide Bonferroni correction | Bonf. | Y |
| Weder et al., 2014 [67] | M: maltreated children<br>HC: healthy controls | M: 94<br>HC: 96 | M: 42<br>HC: 42 | 10.2 (range 5-14) | 17% European/American, 38% Hispanic, 30% African- | M: within six months of an incident of maltreatment of sufficient severity to warrant out-of-home placement | blood: WB | C<br>&<br>E<br>W<br>A<br>S | 450K and Sequenom MassARRAY for validation | age, sex, race | N | EWAS: BDNF CpG sites were NS in association with depression scores after epigenome-wide correction for multiple comparisons<br>Candidate, maltreated vs HC: cg00298481, cg21010859, cg01225698, cg06816235, cg22043168, cg03747251, cg15710245, cg24650785, cg23947039 differentially methylated after Bonferroni correction; hyper vs. hypomethylation information not reported | EWA<br>S: significance threshold set to | Y |
|  |  |  |  |  | American,<br>15%<br>biracial |  |  |  | n of 7<br>CpG<br>sites |  |  | Candidate, association with depression: all CpGs NS after Bonferroni correction | 5.0 x<br>10<sup>-7</sup><br>C:<br>Bonf. |  |
| Unternaehrer<br>et al., 2015<br>[68] | L: low childhood<br>maternal care<br>H: high childhood<br>maternal care | L: 45<br>H: 40 | L: 24.4<br>H:<br>17.5 | L: 30.3<br>(range<br>27.3-33.2)<br>H: 24.3<br>(range<br>22.7-25.9) | European | NR | blood:<br>WB | C | qPCR | batch<br>number,<br>sex, age,<br>BMI,<br>income,<br>mental<br>disorders | N | Higher mean DNAm in L vs. H | *p*<.05 | Y |
| Smith et al.,<br>2017 [69] | Atherosclerosis cohort | 1226 | 48 | 69.6<br>(range 55-95) | 47% non-Hispanic<br>white;<br>22%<br>African<br>American;<br>31%<br>Hispanic | 10 years after<br>enrolling in parent<br>study | blood:<br>MC | C<br>&<br>E<br>W<br>A<br>S | 450K | age, sex,<br>race/ethnici<br>ty,<br>childhood<br>SES, adult<br>SES, and<br>enrichment<br>scores for<br>each of 4<br>major<br>blood cell<br>types<br>(neutrophil<br>s, B cells, T<br>cells, and<br>natural<br>killer cells) | N | C: Neighborhood socioeconomic disadvantage NS<br>C: Worse neighborhood social environment associated with hypermethylation of non-<br>promoter CpG sites; specific sites NR<br>EWAS: cg06979684, cg18595174, cg20108357, cg00298481, cg22043168,<br>cg02527472, cg11718030, and cg01583131 in top 5% of sites associated with<br>neighborhood socioeconomic disadvantage or neighborhood social environment | Bonf.<br>and<br>FDR | Y |
| Thaler et al.,<br>2014 [70] | B: bulimia nervosa<br>NB: no eating disorders<br>Bp: bulimia nervosa,<br>physical abuse<br>Bnp: bulimia nervosa, no<br>physical abuse<br>Bs: bulimia nervosa,<br>sexual abuse<br>Bns: bulimia nervosa, no<br>sexual abuse<br>Bb: bulimia nervosa,<br>bipolar disorder | B: 64<br>NB: 62<br>Bp: 29<br>Bnp: 34<br>Bs: 15<br>Bns: 49<br>Bb: 14<br>Bnb: 47 | B: 0<br>NB: 0<br>Bp: 0<br>Bnp: 0<br>Bs: 0<br>Bns: 0<br>Bb: 0<br>Bnb: 0 | B:<br>26.1±6.6<br>NB:<br>23.7±5.7<br>Bp: NR<br>Bnp: NR<br>Bs: NR<br>Bns: NR<br>Bb: NR<br>Bnb: NR | Canadian | NR | blood:<br>lymph<br>ocytes | C | MassAR<br>RAY | None | N | Hypermethylation in CpGs 2, 17, 19, and 25 in B vs. HC<br>Hypermethylation in CpGs 15, 17, 19, 25, and 28 in Bp vs. HC<br>Hypermethylation in CpGs 2 and 19 in Bnp vs. HC<br>Hypermethylation in CpG19 in Bs, and at CpG2 in Bns, vs. HC<br>Hypermethylation in CpGs 2, 16, 19, and 26 in Bb vs. HC<br>Hypermethylation at CpGs 15, 17, 19, 25, and 28 in Bnb vs. HC | *p*<.05 | Y |
|  | Bnb: bulimia, no bipolar disorder |  |  |  |  |  |  |  |  |  |  | Hypermethylation in CpG11 in HC vs. B and subsamples of Bp, Bnp, Bns, Bnb |  |  |
| Untermaehrer et al., 2012 [71] | Healthy individuals | 76 | 43.4 | 64.1±1.7 | Germany | Prior to and 10 and 90 minutes after stress test | blood: WB | C | MassARRAY | Plate, cell type proportions | N | In unadjusted and adjusted analyses, no differences in mean DNAm across time<br>In unadjusted and adjusted analyses, CpG4/5 decreased from pre- to post-stress time points and slight increase from pre-stress to follow-up | *p*<.05 | Y |
| 27K = Illumina Infinium 27K Human DNA Methylation BeadChip; 450K = Illumina Infinium HumanMethylation 450 BeadChip; BA = Brodmann area; BC = buffy coat; BDNF = brain-derived neurotrophic factor; BH = Benjamini-Hochberg; Bisul-seq = targeted bisulfite sequencing that is not pyrosequencing; BMI = body mass index; C = candidate gene approach; CSF = cerebrospinal fluid; CpG = cytosine-phosphate-guanine; DNAm = DNA methylation; EPIC = Illumina Infinium MethylationEPIC BeadChip; EWAS = epigenome wide association study; FDR = false discovery rate; L= leukocytes; MALDI-TOF = matrix-assisted laser desorption/ionization-time-of-flight; MC = mononuclear cells; Met = methionine; N = no; NR = not reported; NS = non-significant; Pyro = pyrosequencing; qPCR = real-time quantitative polymerase chain reaction; SES = socioeconomic status; Val = valine; WB = whole blood; WS = whole sample; Y = yes<br><sup>1</sup>Sample sizes are reported here for all main groups and subgroups; thus, sample sizes cannot always be summed for the total study sample size<br><sup>2</sup>Mean ± standard deviation in years unless otherwise indicated<br><sup>3</sup>Whether the study examined DNAm in association with *BDNF* genotype<br><sup>4</sup>Statistical significance threshold specified by study authors; *p*<.05 means nominal statistical significance with no correction for multiple comparisons; other entries indicate multiple comparison correction approaches used<br><sup>5</sup>Whether the study is included in the online *BDNF* DNA Methylation Map |  |  |  |  |  |  |  |  |  |  |  |  |  |  |

##### 3.3.2.7. Psychopathology: other

Eight studies examined *BDNF* DNAm in association with other psychopathology phenotypes (Table 8), including borderline personality disorder (BPD) [72, 75], obsessive-compulsive disorder (OCD) [79, 78], first episode of psychosis (FEP) [77], burnout [76], emotion processing [73], and neuroticism [74]. In all studies, DNA was extracted from blood and in one, DNA extracted from saliva was also included. All studies used a candidate gene approach. Six studies adjusted for covariates and two studies corrected for multiple comparisons. Because this is a heterogenous collection of phenotypes, patterns of significant CpG sites are not described (see online *BDNF* DNA Methylation Map for positions of CpG examined in individual studies).

**Table 8.**
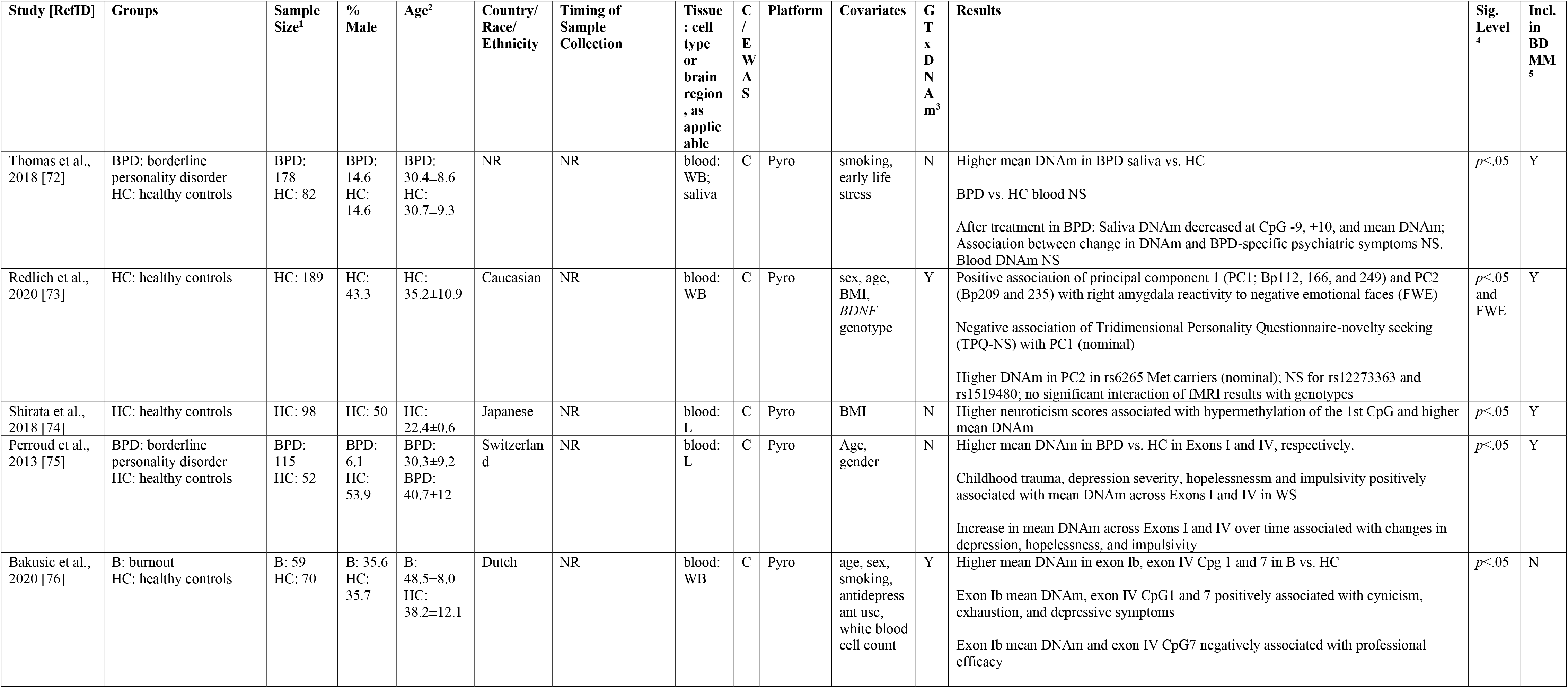
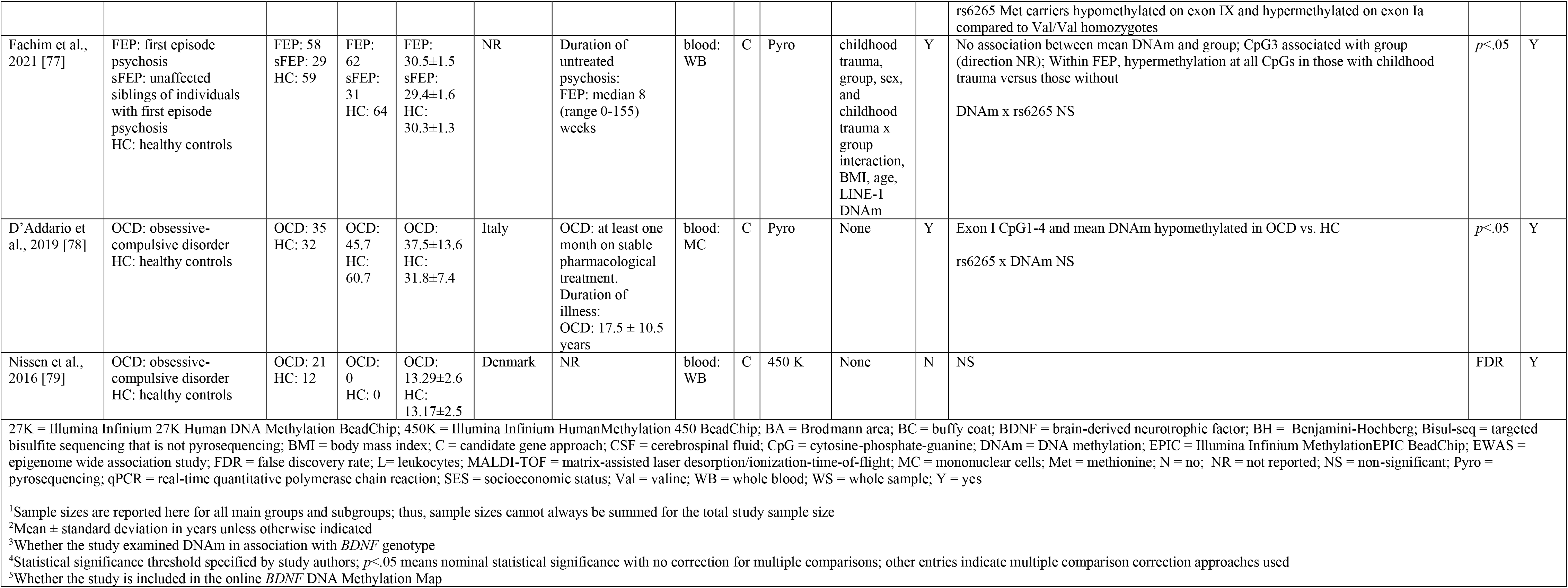
DNAm Studies. Psychopathology: Other.

##### 3.3.2.8. Neurological injury or disorder: dementia and mild cognitive impairment

Eleven studies examined *BDNF* DNAm in association with dementia and mild cognitive impairment (MCI; Table 9), including Alzheimer’s disease (AD) [82, 87, 84, 83, 81, 85, 88], dementia [90, 91], mild cognitive impairment [87, 86, 85, 88], Huntington’s disease [89], and amnesia [87]. Two studies [89 and 88] appear to report results from the same cohort of individuals. In nine studies DNA was extracted from blood, in two from brain, and in one from both blood and buccal tissue. All studies used a candidate approach. Five studies adjusted for covariates and four corrected for multiple comparisons. Of studies for which positions could be identified, examined CpG sites were located across all exon/promoter regions with relative enrichment in exons/promoters I, IV, and VI. Significant individual CpG sites and region-based mean DNAm were generally evenly distributed across examined regions.

**Table 9.**
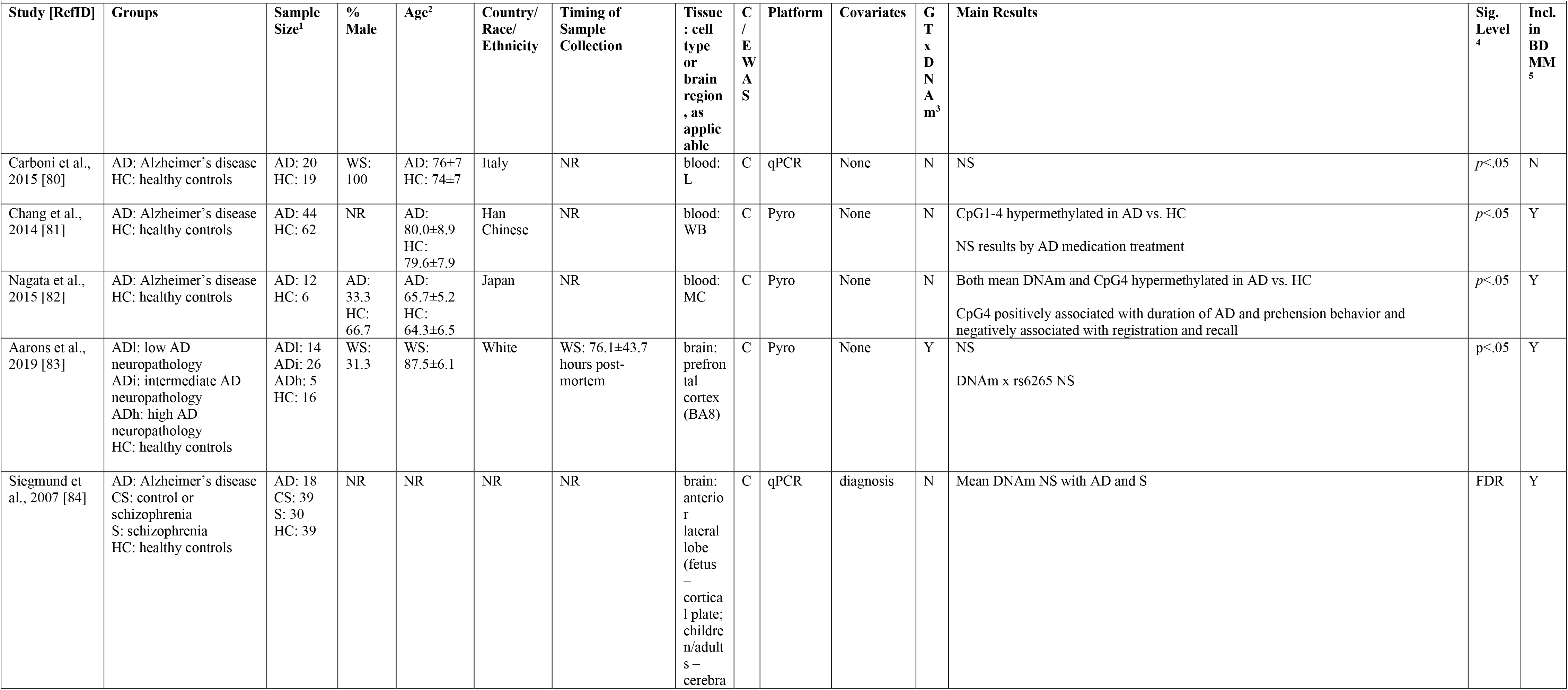
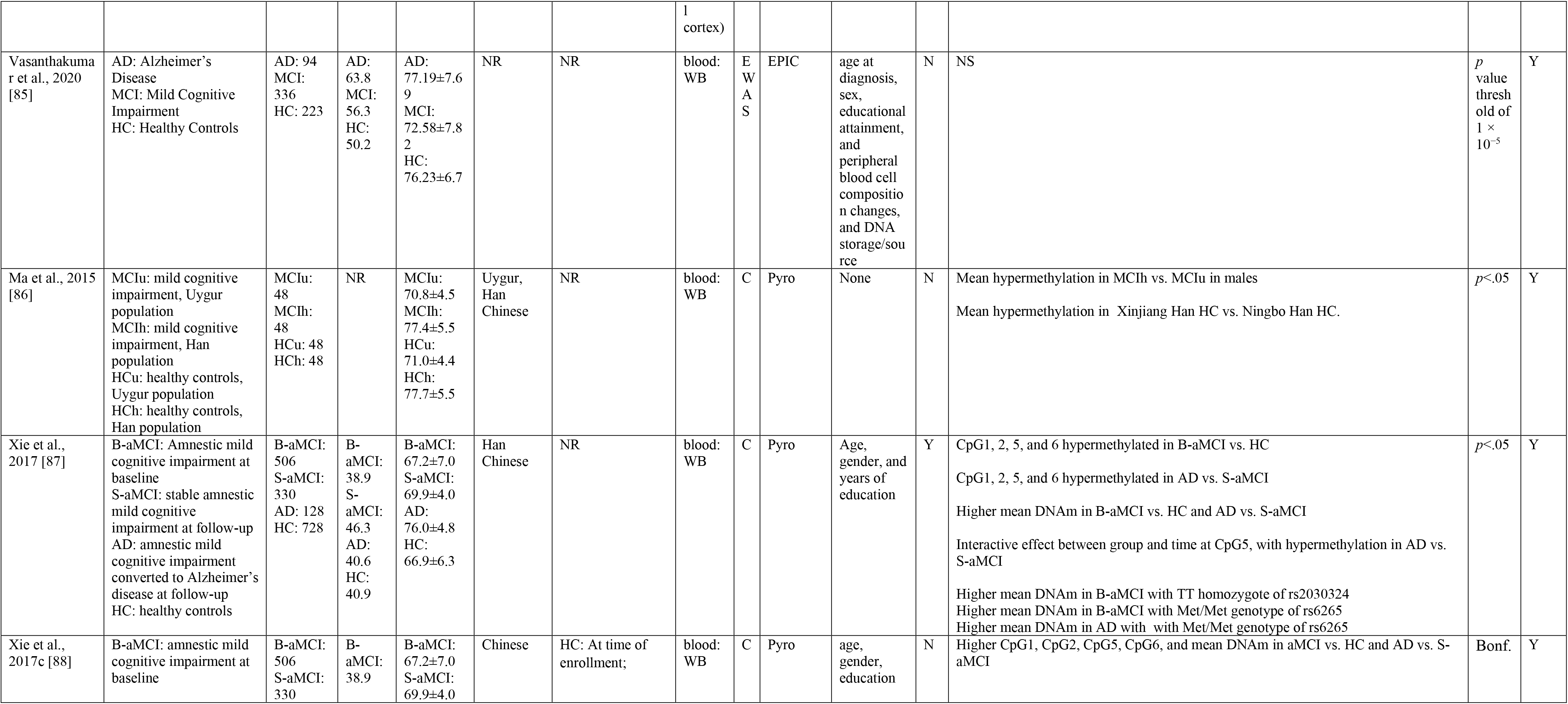
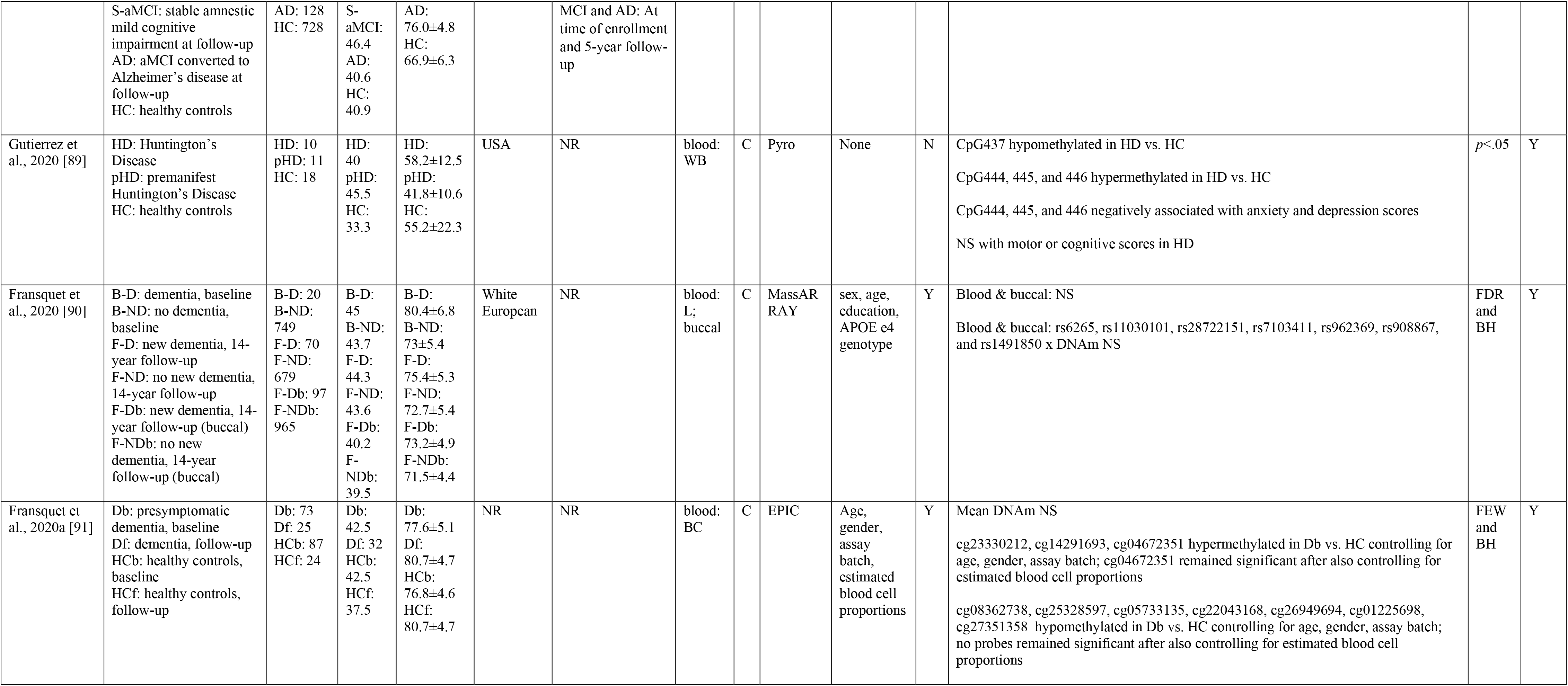
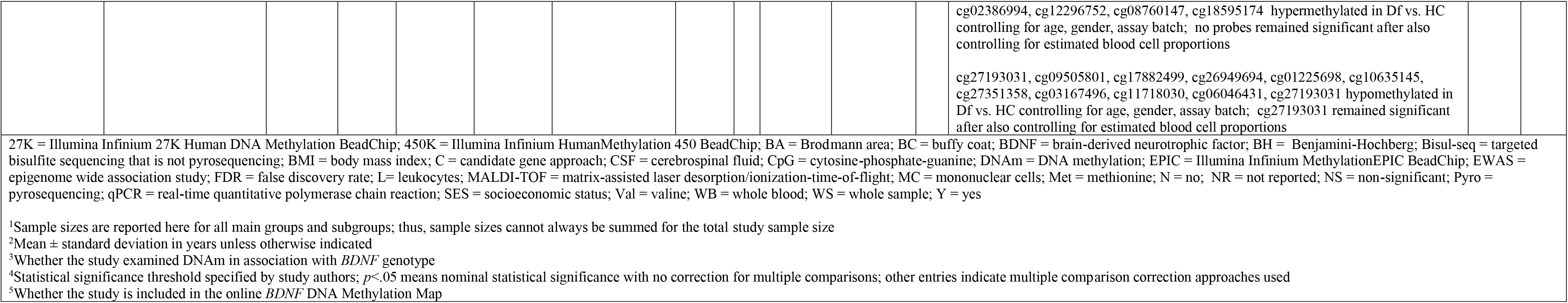
DNAm Studies. Neurological injury or disorder: Dementia & Mild Cognitive Impairment.

##### 3.3.2.9. Neurological injury or disorder: stroke

Five studies examined *BDNF* DNAm in association with stroke outcomes, including global outcome, physical disability, cognitive dysfunction, anxiety, depression, and response to aerobic exercise training (Table 10). Of these, three studies [92, 94, 93] appear to report results from the same cohort of individuals. In four studies DNA was extracted from blood and in one from muscle. All studies used a candidate approach. Four studies adjusted for covariates and one corrected for multiple comparisons. Of studies for which positions could be identified, examined CpG sites were located primarily in exon VI, in addition to four other CpG sites in promoter VII, downstream of exon VII, and in exon IX, respectively. All the studies of exon VI were from the same cohort [92, 93, 94] and reported several significant individual CpG sites and region-based mean DNAm. The other four CpG sites examined in the other cohorts and genomic regions were also significant.

**Table 10.**
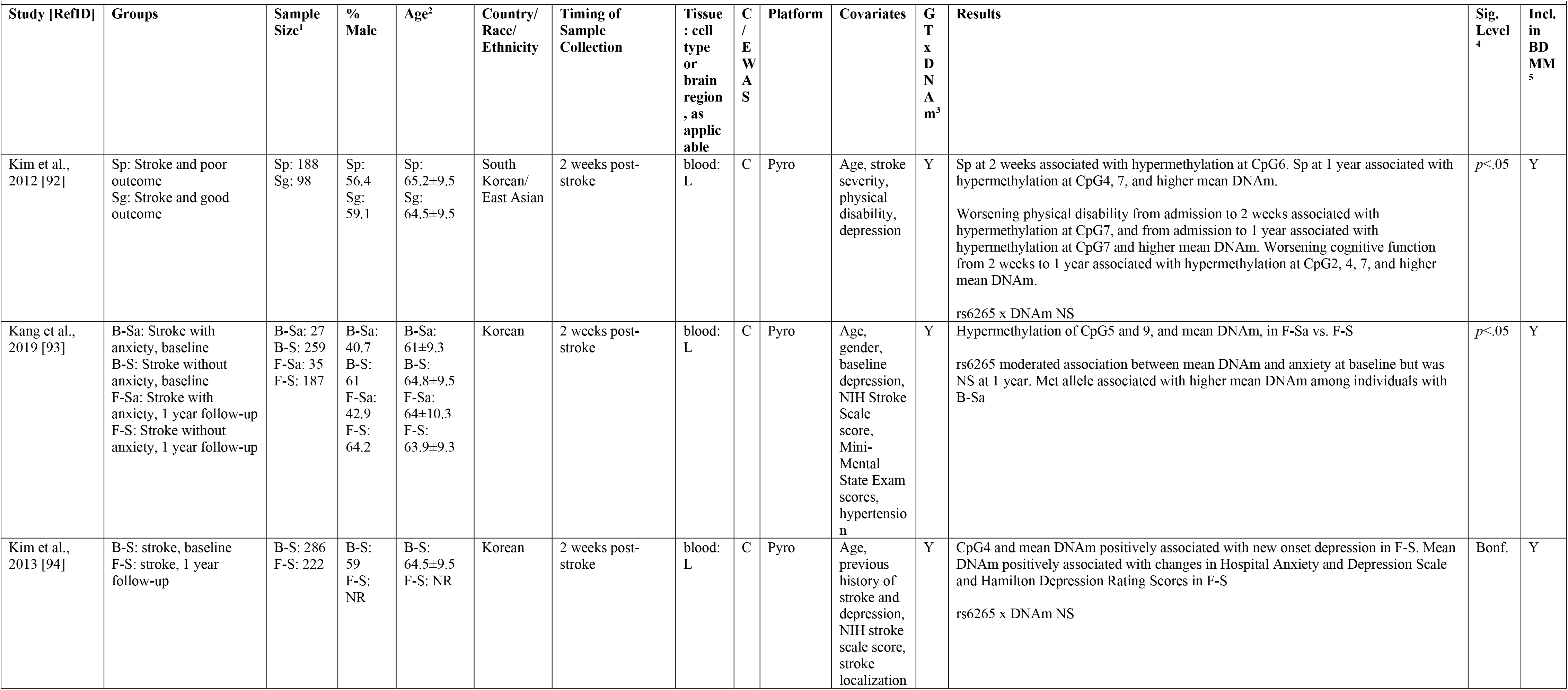
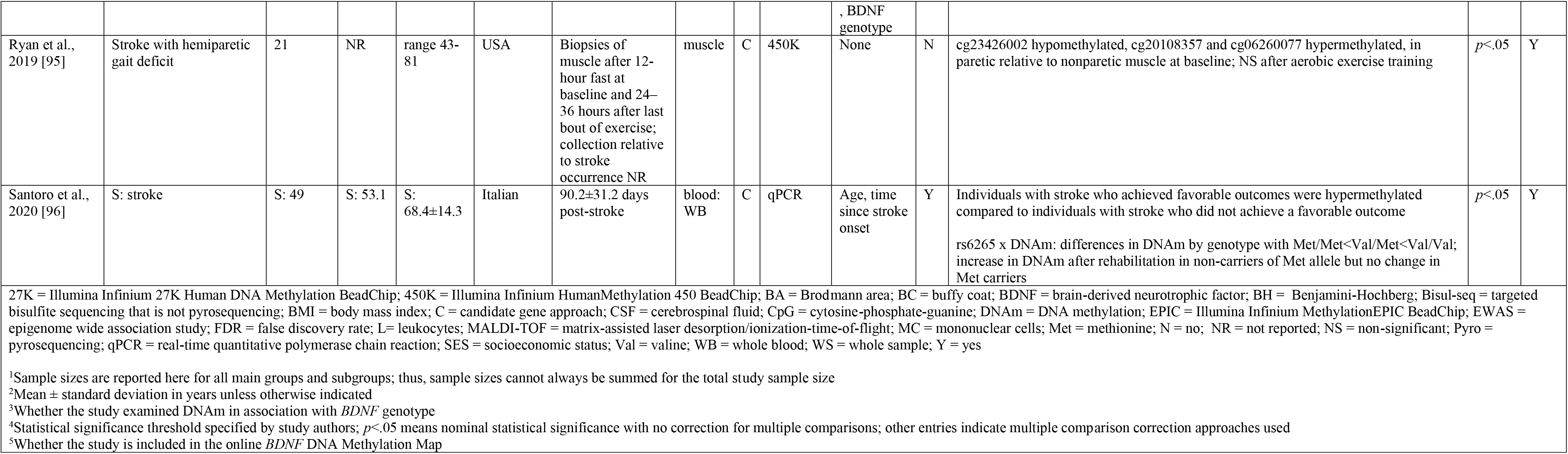
DNAm Studies. Neurological injury or disorder: Stroke.

##### 3.3.2.10. Neurological injury or disorder: other

Seven studies examined *BDNF* DNAm in association with other neurological injury or disorder phenotypes (Table 11). Two were studies of seizures/epilepsy, one in adults [97] and one at birth, during childhood, and during adolescence [98]. Study [99] examined adults with multiple sclerosis. Study [100] examined adults with severe traumatic brain injury (TBI) and was notably the only study included in the present review to examine trajectories of DNAm over time, in this case over the first 5 days post-TBI. Three studies focused on pediatric conditions; study [101] examined children with symptoms of attention-deficit/hyperactivity disorder, study [102] examined newborns with extremely low gestational age, and study [103] examined children with a history of fetal growth restriction with or without brain sparing.

Tissues for DNA extraction were varied, including brain, blood, cerebrospinal fluid, buccal, and placenta. Study [98] was one of only four EWASs included in the present review with significant results in *BDNF*, with Illumina probe cg13974632 hypermethylated in blood of adolescents with seizures vs. those without. The other six studies used a candidate approach. One of the two studies in the present review that tested for replication was in this phenotype [98]. All but one study adjusted for covariates. Two studies corrected for multiple comparisons. Because this is a heterogenous collection of phenotypes, patterns of significant CpG sites are not described (see online BDNF DNA Methylation Map for positions of CpG examined in individual studies).

**Table 11.**
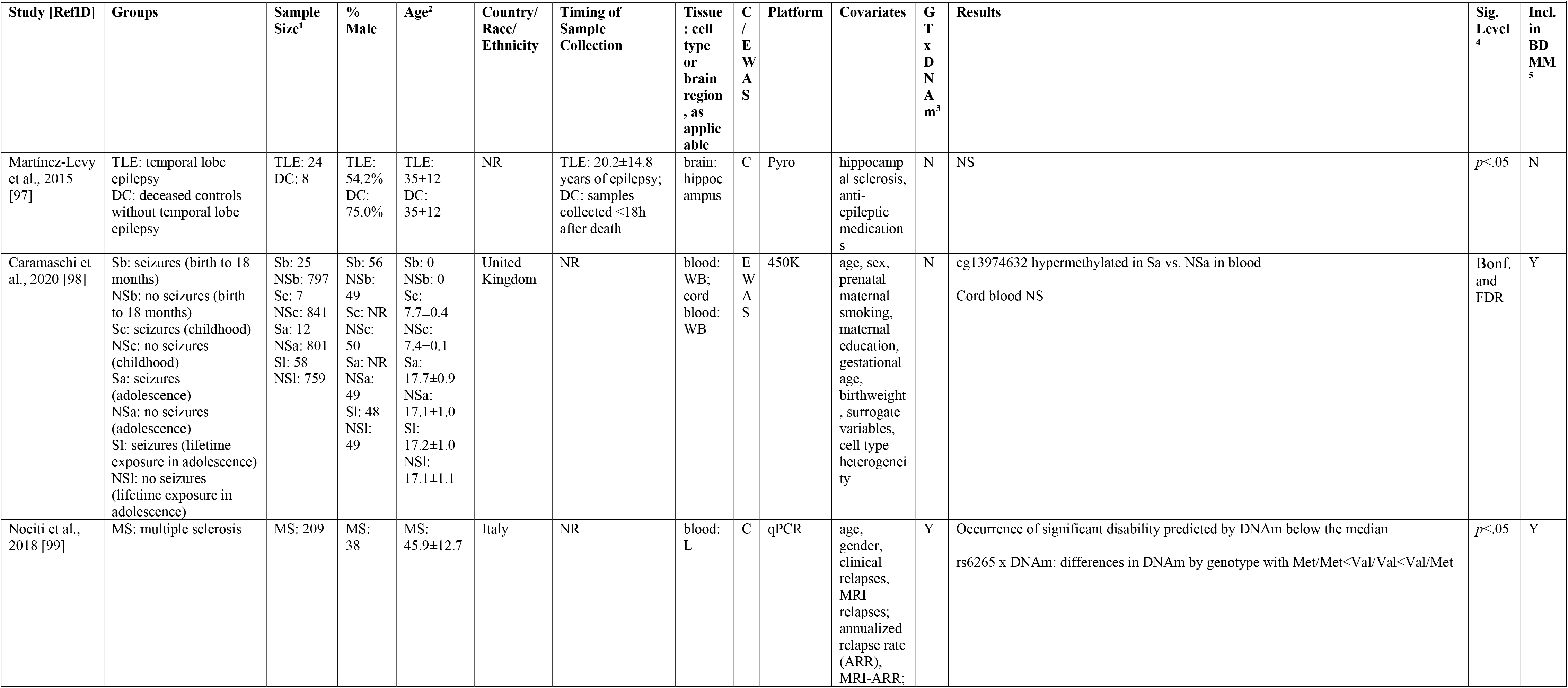
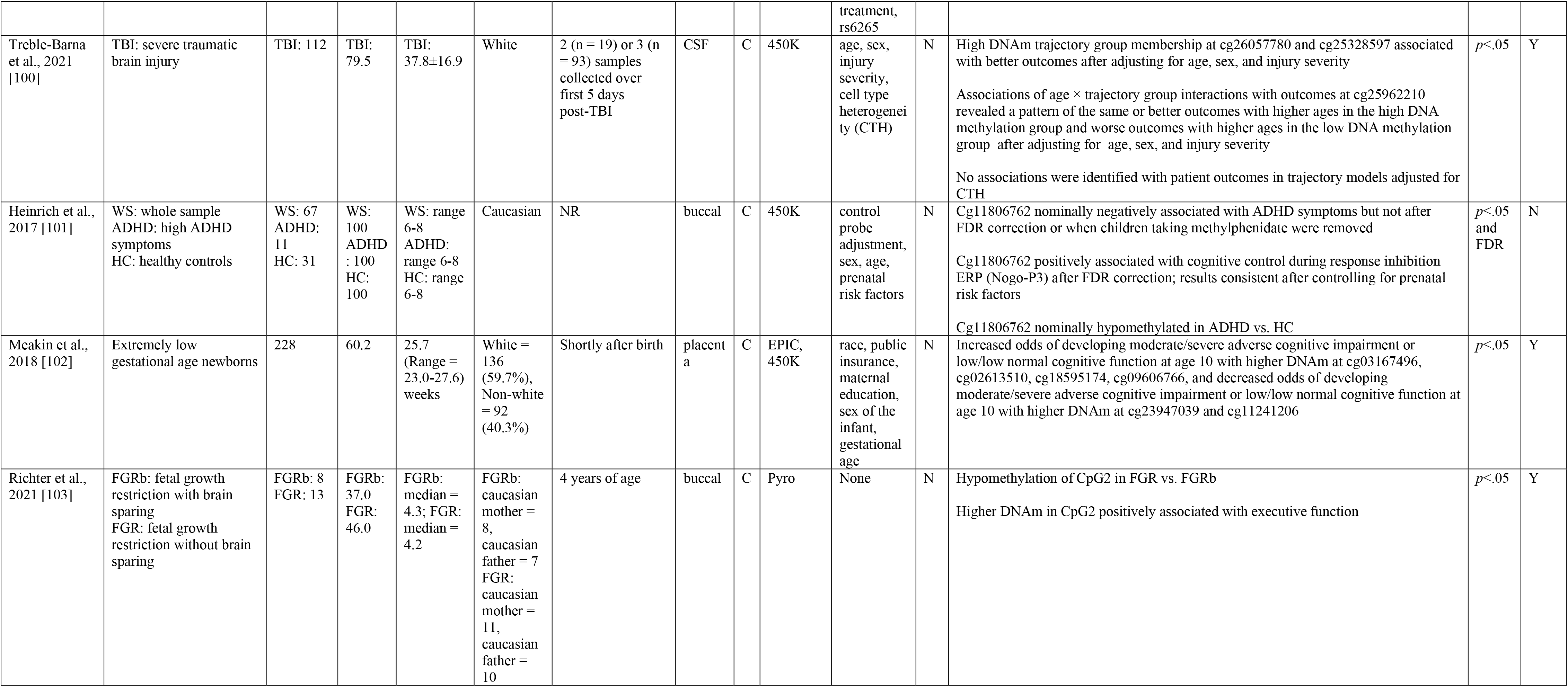
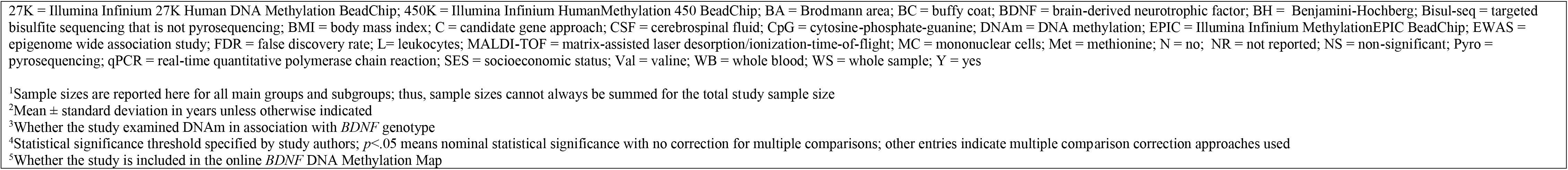
DNAm Studies. Neurological injury or disorder: Other.

###### 3.3.2.11. Chronic pain/fatigue

Three studies examined *BDNF* DNAm in association with chronic musculoskeletal pain, fibromyalgia, and chronic fatigue syndrome (Table 12). In all studies DNA was extracted DNA from blood. Two were candidate gene studies [104, 106] and one used an EWAS approach [105]. This EWAS study [105] was one of only four EWASs included in the present review with significant results in *BDNF*, with Illumina probe cg04481212 differentially methylated between individuals with fibromyalgia and healthy controls. One study adjusted for covariates and one study corrected for multiple comparisons. Of studies for which positions could be identified, examined CpG sites were located across all exon/promoter regions. Significant CpG sites were located in exon I promoter, downstream of exon III, and in exon IX.

**Table 12.** DNAm Studies. Chronic Pain/Fatigue.

| Table 12. DNAm Studies. Chronic Pain/Fatigue |  |  |  |  |  |  |  |  |  |  |  |  |  |  |
| --- | --- | --- | --- | --- | --- | --- | --- | --- | --- | --- | --- | --- | --- | --- |
| Study [RefID] | Groups | Sample Size<sup>1</sup> | % Male | Age<sup>2</sup> | Country/<br>Race/<br>Ethnicity | Timing of<br>Sample<br>Collection | Tissue<br>: cell<br>type<br>or<br>brain<br>region<br>, as<br>applic<br>able | C<br>/<br>E<br>W<br>A<br>S | Platform | Covariates | G<br>T<br>x<br>D<br>N<br>A<br>m<sup>3</sup> | Results | Sig.<br>Level<br><sup>4</sup> | Incl.<br>in<br>BD<br>MM<br><sup>5</sup> |
| Paoloni-Giacobino et<br>al., 2020 [104] | CMSP: chronic<br>musculoskeletal pain<br>HC: healthy controls | CMSP:<br>58<br>HC: 18 | CMSP:<br>79.0<br>HC:<br>61.0 | CMSP:<br>41±13<br>HC:<br>40±11 | NR | CMSP: Median<br>(range) time since<br>injury 436 (39-5331) days | blood:<br>WB | C | Pyro | age, sex,<br>level of<br>comorbiditi<br>es | N | Higher mean DNAm in CMSP vs. HC<br>Negative association between biopsychosocial complexity and mean DNAm, but NS<br>with severity of pain, degree of anxiety, depression, kinesiophobia, catastrophism | *p*<.05 | Y |
| Menzies et al.,<br>2013 [105] | F: fibromyalgia<br>HC: healthy controls | F: 10<br>HC: 8 | F: 0<br>HC: 0 | F:<br>48.2±6.7<br>HC: NR | F: 5<br>White; 4<br>Black; 1<br>"other"<br>race<br>category | F: Years since<br>diagnosis: range =<br>1- 19 years | blood:<br>WB | E<br>W<br>A<br>S | 450K | None | N | cg04481212 differentially methylated between F vs. HC; direction NR | FDR | Y |
| Polli et al.,<br>2020 [106] | CFS/FM: chronic fatigue<br>syndrome/fibromyalgia<br>HC: healthy controls | CFS/FM:<br>28<br>HC: 26 | CFS/F<br>M: 0<br>HC: 0 | CFS/FM:<br>48.0±9.4<br>HC:<br>52.0±10.9 | NR | Before and 3 days<br>after assessment<br>CFS/FM:<br>Duration of<br>illness: 9.15 ± 6.5<br>years | blood:<br>WB | C | Pyro | None | Y | Lower mean DNAm in exon 9 and 9b in CFS/FM vs. HC<br>rs6265 associated with mean DNAm in exon 9 across groups (higher for Val vs. Met<br>allele) | *p*<.05 | Y |
| 27K = Illumina Infinium 27K Human DNA Methylation BeadChip; 450K = Illumina Infinium HumanMethylation 450 BeadChip; BA = Brodmann area; BC = buffy coat; BDNF = brain-derived neurotrophic factor; BH = Benjamini-Hochberg; Bisul-seq = targeted bisulfite sequencing that is not pyrosequencing; BMI = body mass index; C = candidate gene approach; CSF = cerebrospinal fluid; CpG = cytosine-phosphate-guanine; DNAm = DNA methylation; EPIC = Illumina Infinium MethylationEPIC BeadChip; EWAS = epigenome wide association study; FDR = false discovery rate; L= leukocytes; MALDI-TOF = matrix-assisted laser desorption/ionization-time-of-flight; MC = mononuclear cells; Met = methionine; N = no; NR = not reported; NS = non-significant; Pyro = pyrosequencing; qPCR = real-time quantitative polymerase chain reaction; SES = socioeconomic status; Val = valine; WB = whole blood; WS = whole sample; Y = yes |  |  |  |  |  |  |  |  |  |  |  |  |  |  |
| <sup>1</sup>Sample sizes are reported here for all main groups and subgroups; thus, sample sizes cannot always be summed for the total study sample size |  |  |  |  |  |  |  |  |  |  |  |  |  |  |
| <sup>2</sup>Mean ± standard deviation in years unless otherwise indicated |  |  |  |  |  |  |  |  |  |  |  |  |  |  |
| <sup>3</sup>Whether the study examined DNAm in association with *BDNF* genotype |  |  |  |  |  |  |  |  |  |  |  |  |  |  |
| <sup>4</sup>Statistical significance threshold specified by study authors; *p*<.05 means nominal statistical significance with no correction for multiple comparisons; other entries indicate multiple comparison correction approaches used |  |  |  |  |  |  |  |  |  |  |  |  |  |  |
| <sup>5</sup>Whether the study is included in the online *BDNF* DNA Methylation Map |  |  |  |  |  |  |  |  |  |  |  |  |  |  |

###### 3.3.2.12. Pediatric cognition or behavior

Three studies examined *BDNF* DNAm in association with pediatric cognition or behavior (Table 13). Study [107] examined cognitive performance in adolescents at risk for congenital adrenal hyperplasia prenatally administered dexamethasone [107]. Study [108] examined early onset conduct problems and prenatal environmental exposures. Study [109] examined childhood BPA exposure and behavioral function in adolescent boys. In all three studies DNA was extracted from blood. Two studies used both EWAS and candidate approaches, with significant *BDNF* results in the candidate analyses only; the third study used a candidate approach. All studies adjusted for covariates and two studies corrected for multiple comparisons. Of studies for which positions could be identified, examined CpG sites were located across all exon/promoter regions. Significant CpG sites were located in exon II promoter, exon/promoter IV, exon/promoter V, exon VI promoter, exon VII promoter, and slightly upstream of exon VIII.

**Table 13.**
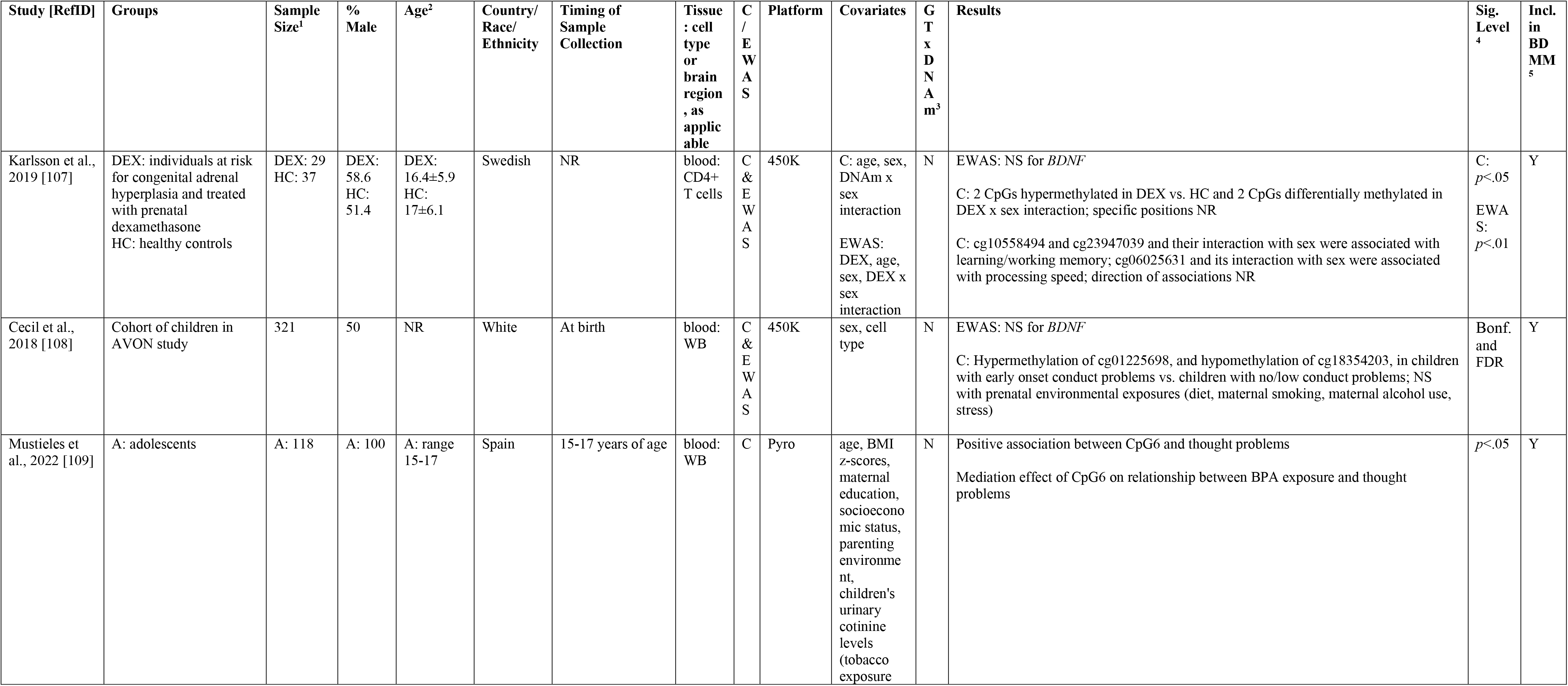
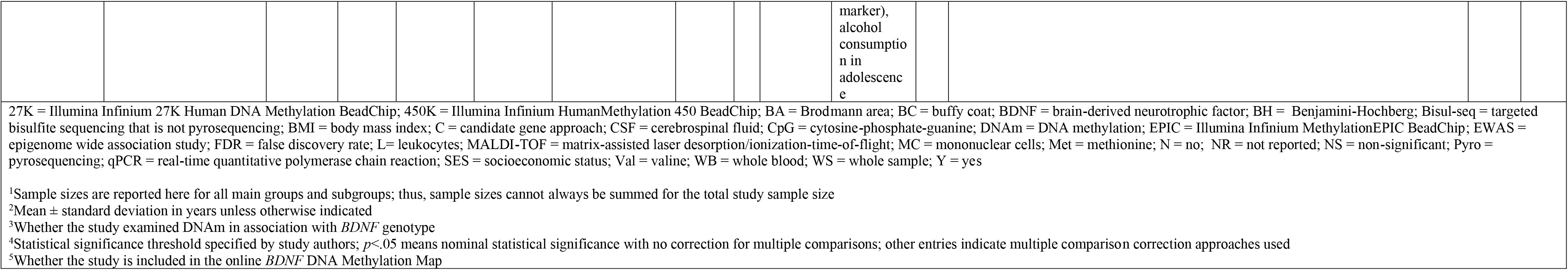
DNAm Studies. Pediatric Cognition or Behavior.

###### 3.3.2.13. Aging

Three studies examined *BDNF* DNAm in association with aging (Table 14). DNA was extracted from brain [110], blood [111], and both blood and brain [112]. All three studies used a candidate gene approach. One study adjusted for covariates and two studies corrected for multiple comparisons. Of studies for which positions could be identified, examined CpG sites were located across all exon/promoter regions. Significant individual CpG sites and region-based mean DNAm were located in exon/promoter I, exon/promoter II, exon III and slightly downstream from exon III, and exon VI promoter.

**Table 14.**
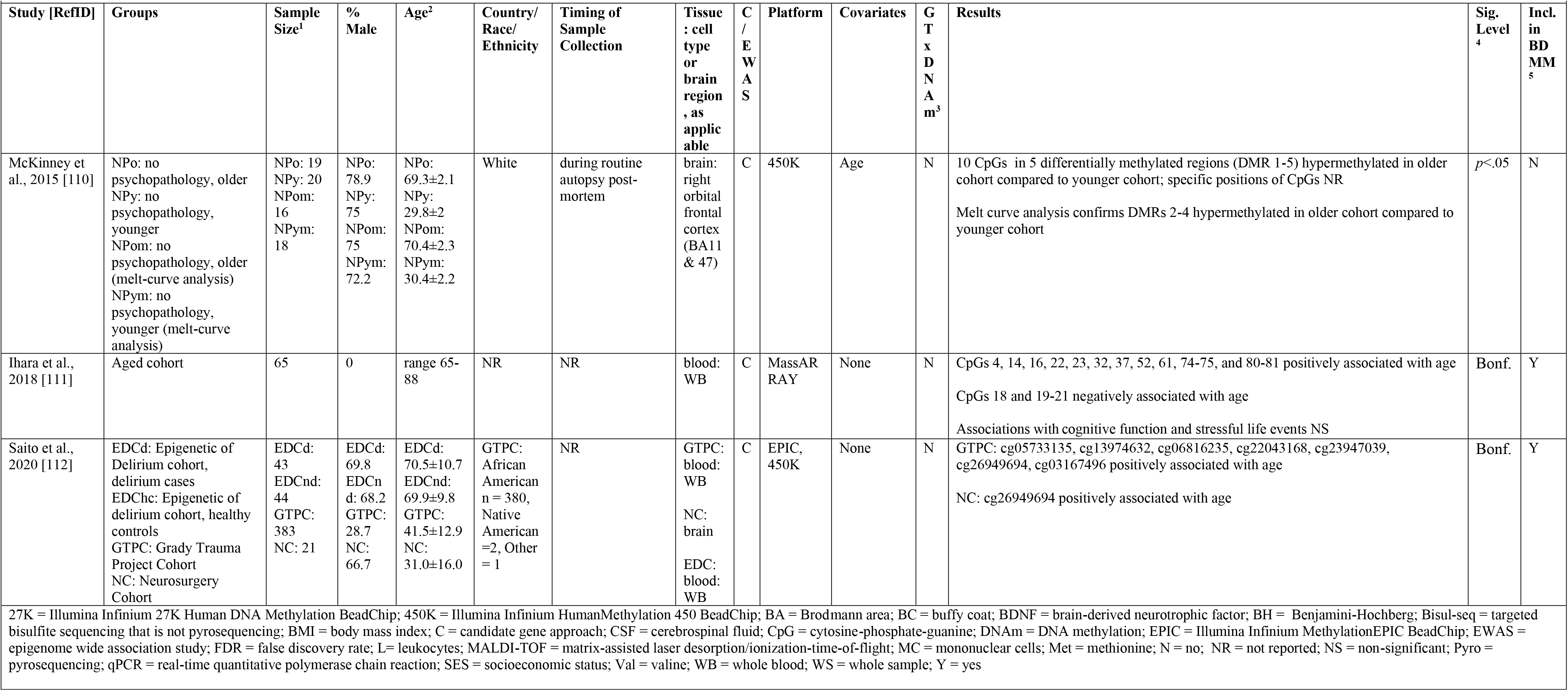
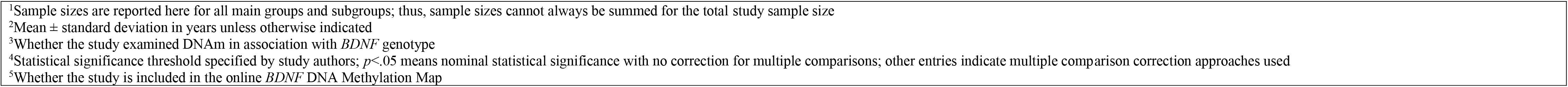
DNAm Studies. Aging.

| Study [RefID] | Groups | Sample Size<sup>1</sup> | % Male | Age<sup>2</sup> | Country/<br>Race/<br>Ethnicity | Timing of<br>Sample<br>Collection | Tissue<br>: cell<br>type<br>or<br>brain<br>region<br>, as<br>applic<br>able | C<br>/<br>E<br>W<br>A<br>S | Platform | Covariates | G<br>T<br>x<br>D<br>N<br>A<br>m<sup>3</sup> | Results | Sig.<br>Level<br><sup>4</sup> | Incl.<br>in<br>BD<br>MM<br><sup>5</sup> |
| --- | --- | --- | --- | --- | --- | --- | --- | --- | --- | --- | --- | --- | --- | --- |
| McKinney et al., 2015 [110] | NPo: no psychopathology, older<br>NPY: no psychopathology, younger<br>NPom: no psychopathology, older (melt-curve analysis)<br>NPym: no psychopathology, younger (melt-curve analysis) | NPo: 19<br>NPY: 20<br>NPom: 16<br>NPym: 18 | NPo: 78.9<br>NPY: 75<br>NPom: 75<br>NPym: 72.2 | NPo: 69.3±2.1<br>NPY: 29.8±2<br>NPom: 70.4±2.3<br>NPym: 30.4±2.2 | White | during routine autopsy post-mortem | brain: right orbital frontal cortex (BA11 & 47) | C | 450K | Age | N | 10 CpGs in 5 differentially methylated regions (DMR 1-5) hypermethylated in older cohort compared to younger cohort; specific positions of CpGs NR<br>Melt curve analysis confirms DMRs 2-4 hypermethylated in older cohort compared to younger cohort | *p*<.05 | N |
| Ihara et al., 2018 [111] | Aged cohort | 65 | 0 | range 65-88 | NR | NR | blood: WB | C | MassARRAY | None | N | CpGs 4, 14, 16, 22, 23, 32, 37, 52, 61, 74-75, and 80-81 positively associated with age<br>CpGs 18 and 19-21 negatively associated with age<br>Associations with cognitive function and stressful life events NS | Bonf. | Y |
| Saito et al., 2020 [112] | EDCd: Epigenetic of Delirium cohort, delirium cases<br>EDChc: Epigenetic of delirium cohort, healthy controls<br>GTPC: Grady Trauma Project Cohort<br>NC: Neurosurgery Cohort | EDCd: 43<br>EDCnd: 44<br>GTPC: 383<br>NC: 21 | EDCd: 69.8<br>EDCnd: 68.2<br>GTPC: 28.7<br>NC: 66.7 | EDCd: 70.5±10.7<br>EDCnd: 69.9±9.8<br>GTPC: 41.5±12.9<br>NC: 31.0±16.0 | GTPC: African American n = 380, Native American =2, Other = 1 | NR | GTPC: blood: WB<br>NC: brain<br>EDC: blood: WB | C | EPIC, 450K | None | N | GTPC: cg05733135, cg13974632, cg06816235, cg22043168, cg23947039, cg26949694, cg03167496 positively associated with age<br>NC: cg26949694 positively associated with age | Bonf. | Y |

27K = Illumina Infinium 27K Human DNA Methylation BeadChip; 450K = Illumina Infinium HumanMethylation 450 BeadChip; BA = Brodmann area; BC = buffy coat; BDNF = brain-derived neurotrophic factor; BH = Benjamini-Hochberg; Bisul-seq = targeted bisulfite sequencing that is not pyrosequencing; BMI = body mass index; C = candidate gene approach; CSF = cerebrospinal fluid; CpG = cytosine-phosphate-guanine; DNAm = DNA methylation; EPIC = Illumina Infinium MethylationEPIC BeadChip; EWAS = epigenome wide association study; FDR = false discovery rate; L= leukocytes; MALDI-TOF = matrix-assisted laser desorption/ionization-time-of-flight; MC = mononuclear cells; Met = methionine; N = no; NR = not reported; NS = non-significant; Pyro = pyrosequencing; qPCR = real-time quantitative polymerase chain reaction; SES = socioeconomic status; Val = valine; WB = whole blood; WS = whole sample; Y = yes
<sup>1</sup>Sample sizes are reported here for all main groups and subgroups; thus, sample sizes cannot always be summed for the total study sample size
<sup>2</sup>Mean ± standard deviation in years unless otherwise indicated
<sup>3</sup>Whether the study examined DNAm in association with *BDNF* genotype
<sup>4</sup>Statistical significance threshold specified by study authors; $p<.05$ means nominal statistical significance with no correction for multiple comparisons; other entries indicate multiple comparison correction approaches used
<sup>5</sup>Whether the study is included in the online *BDNF* DNA Methylation Map

#### 3.3.3 Transdiagnostic overview of non-coding RNA studies

##### 3.3.3.1. Summary table

Details of each non-coding RNA study are summarized and grouped by broad phenotype in Table 15. Consistent with the DNAm summary tables, the directions of statistically significant associations (e.g. higher miRNA; positive or negative association) are specified, non-significant associations are reported as “NS”, results are summarized after covariate adjustment when applicable (see “Covariates” column), and statistical significance is consistent with the study-specific approach and authors’ definition of “significance”—regardless of adjustment for multiple comparisons.

**Table 15.**
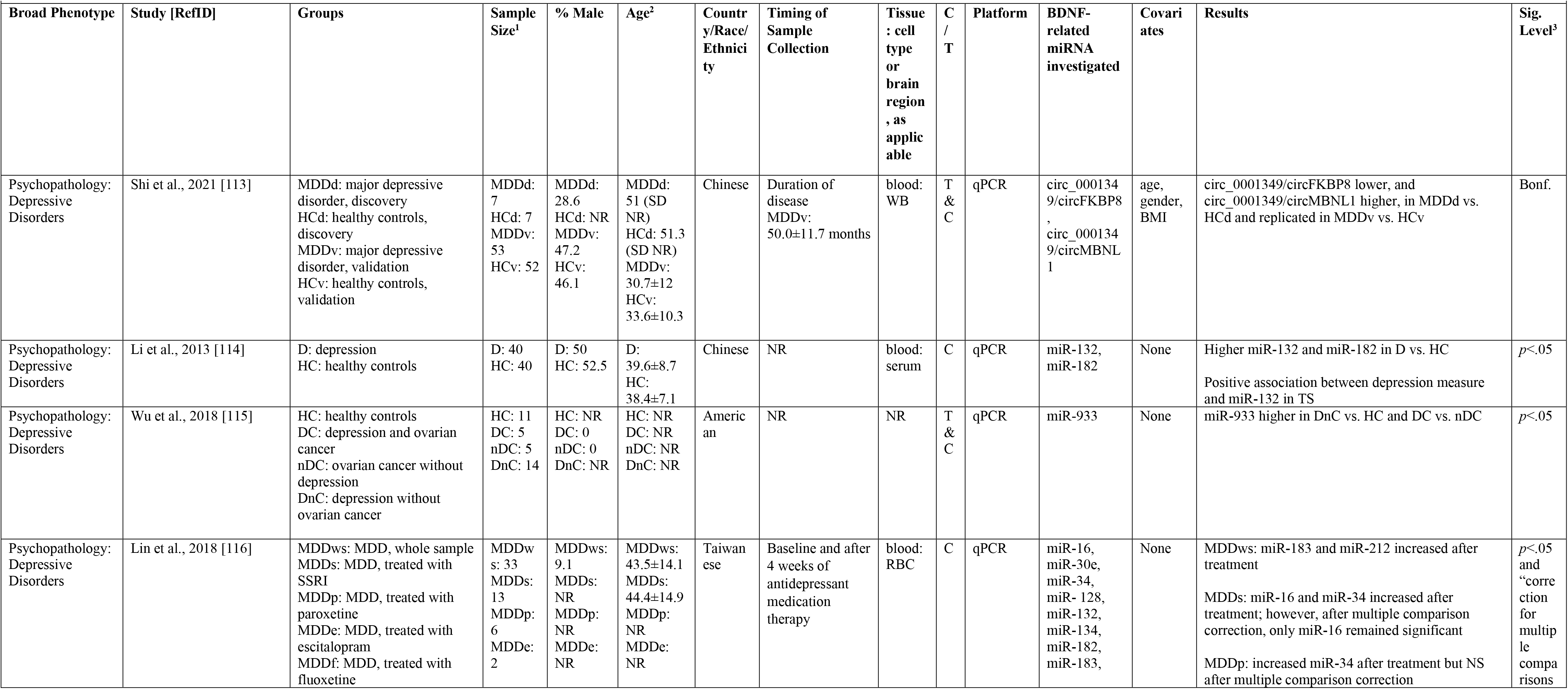
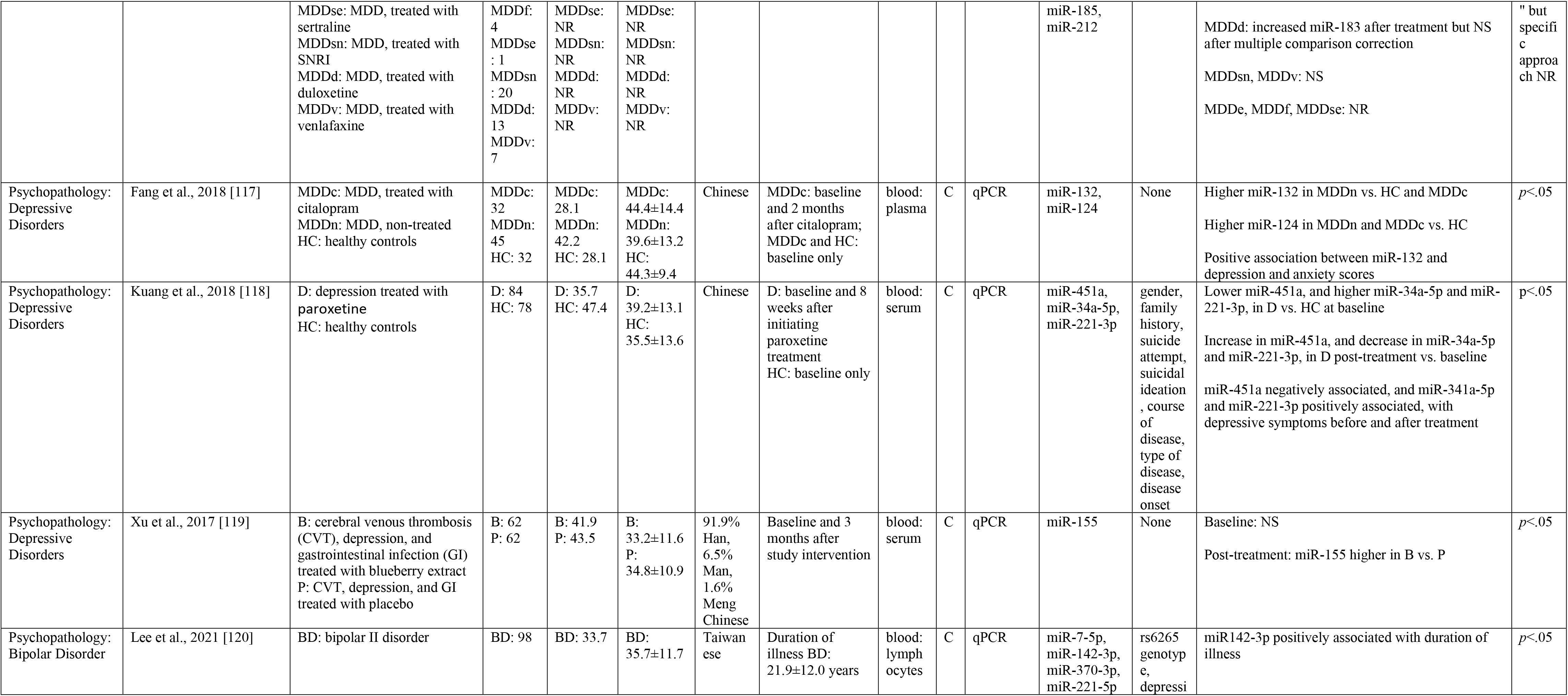
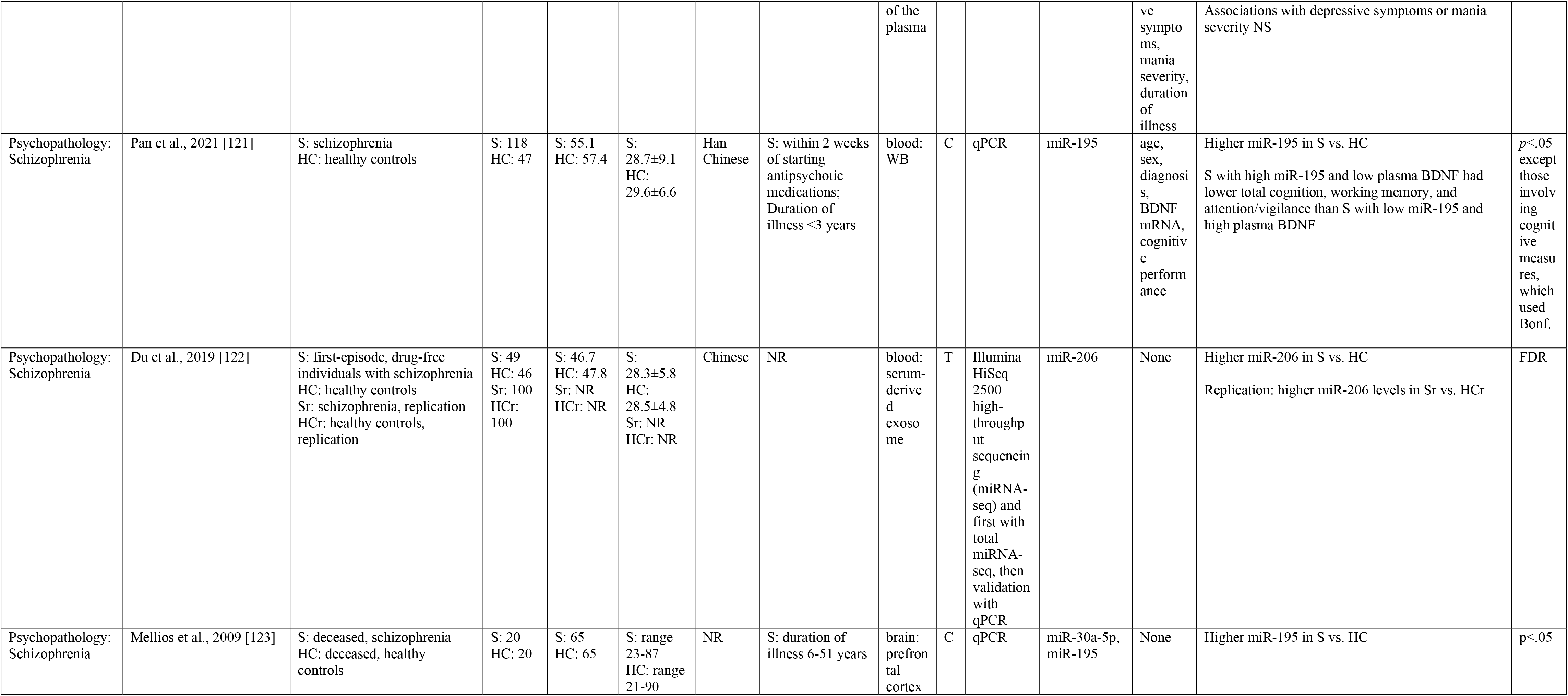
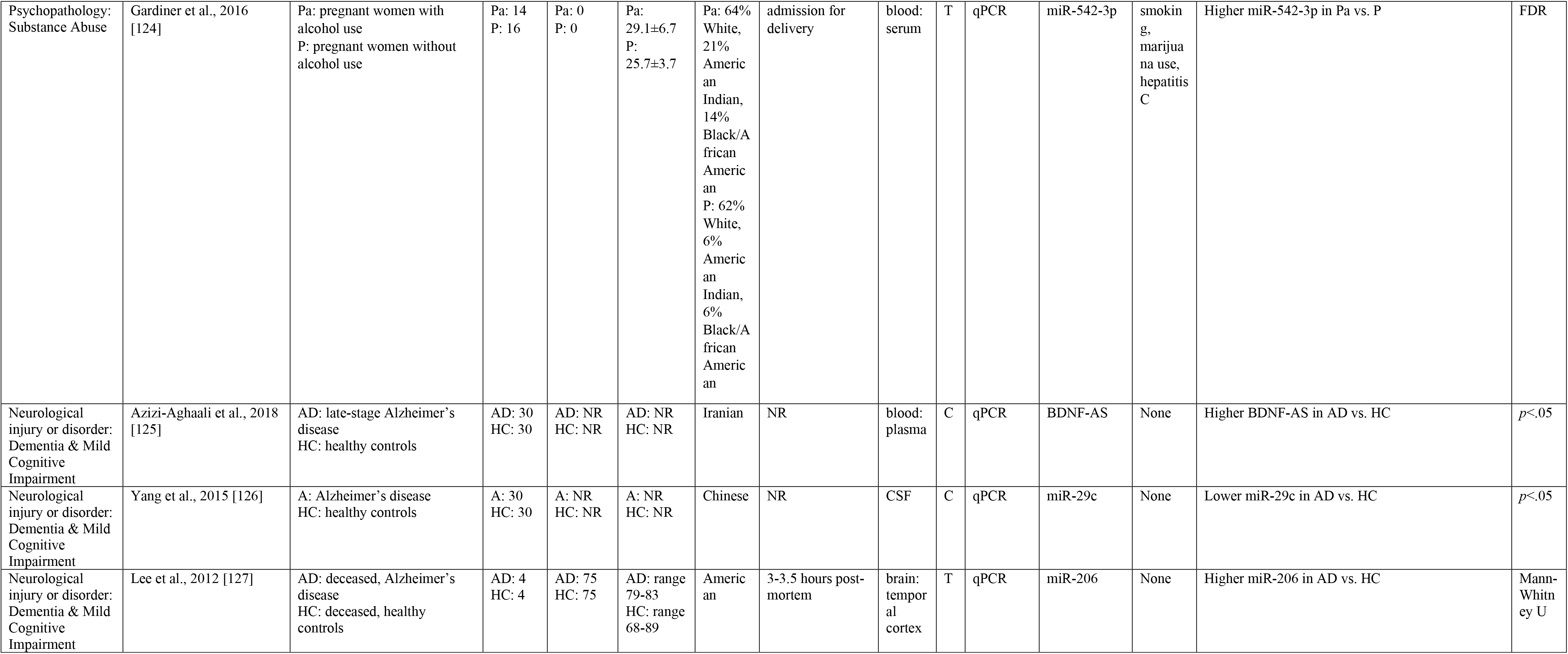
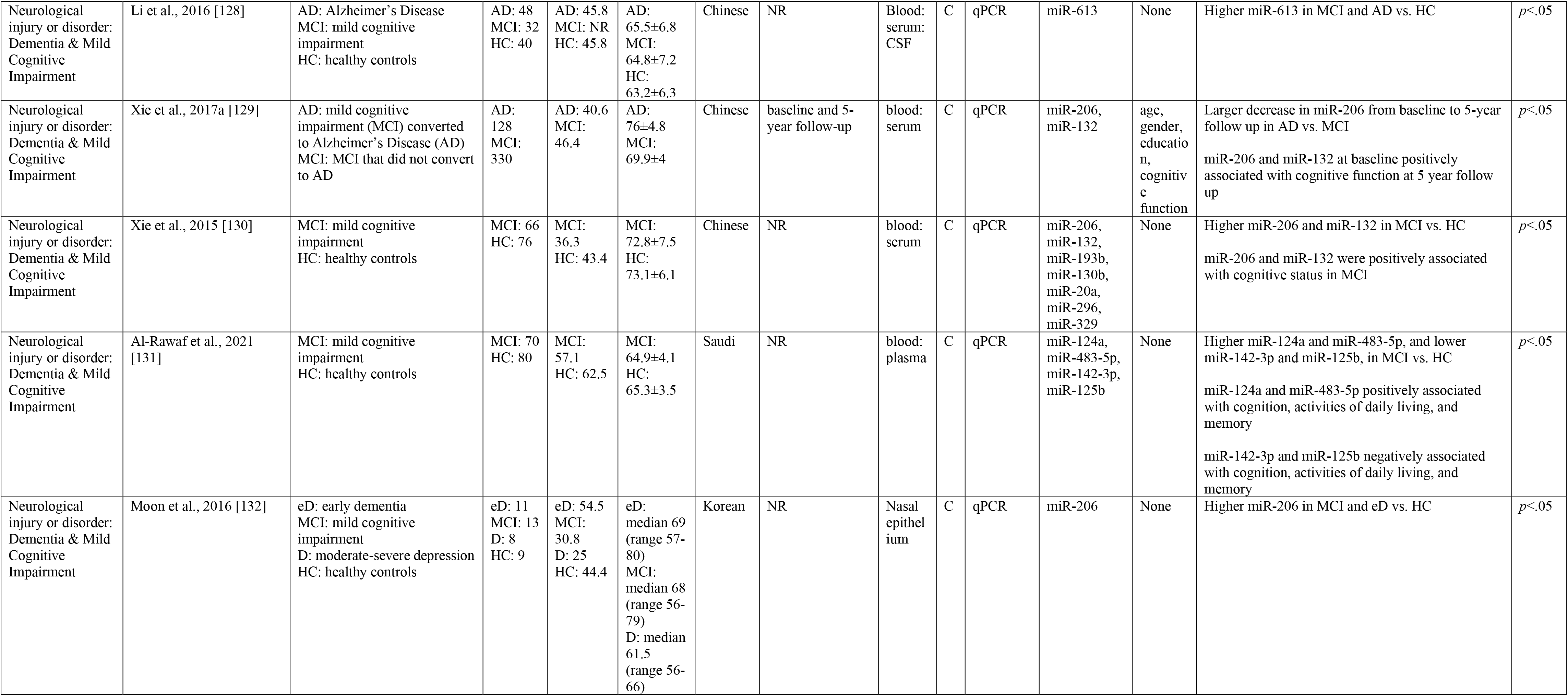
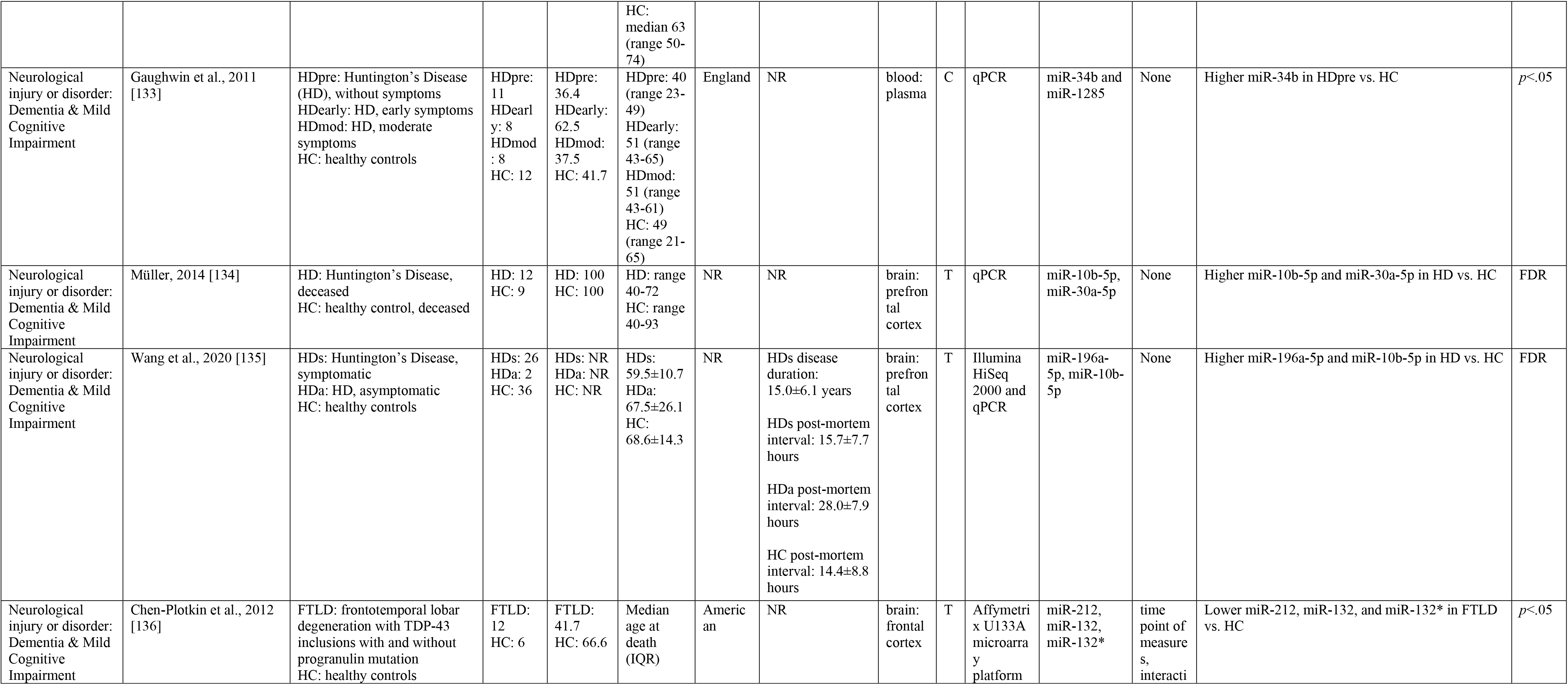
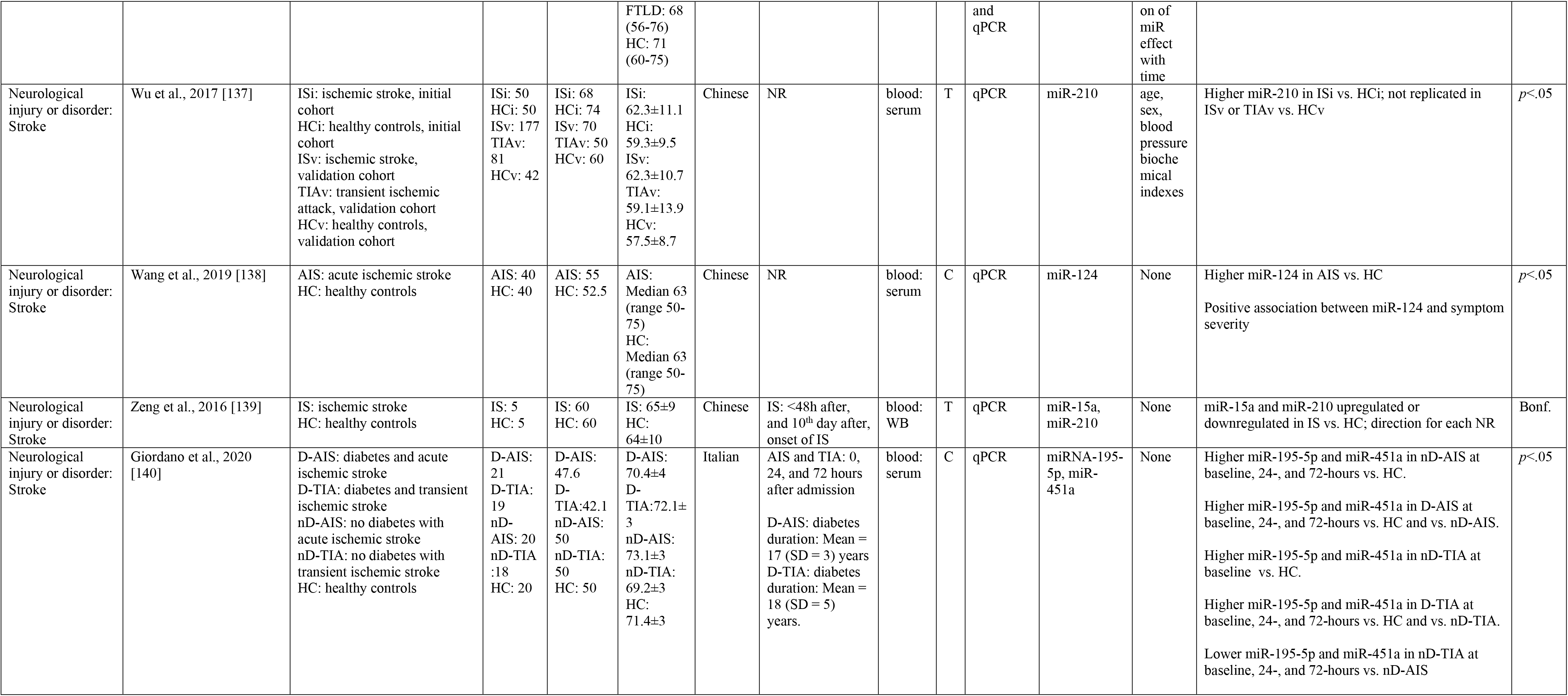
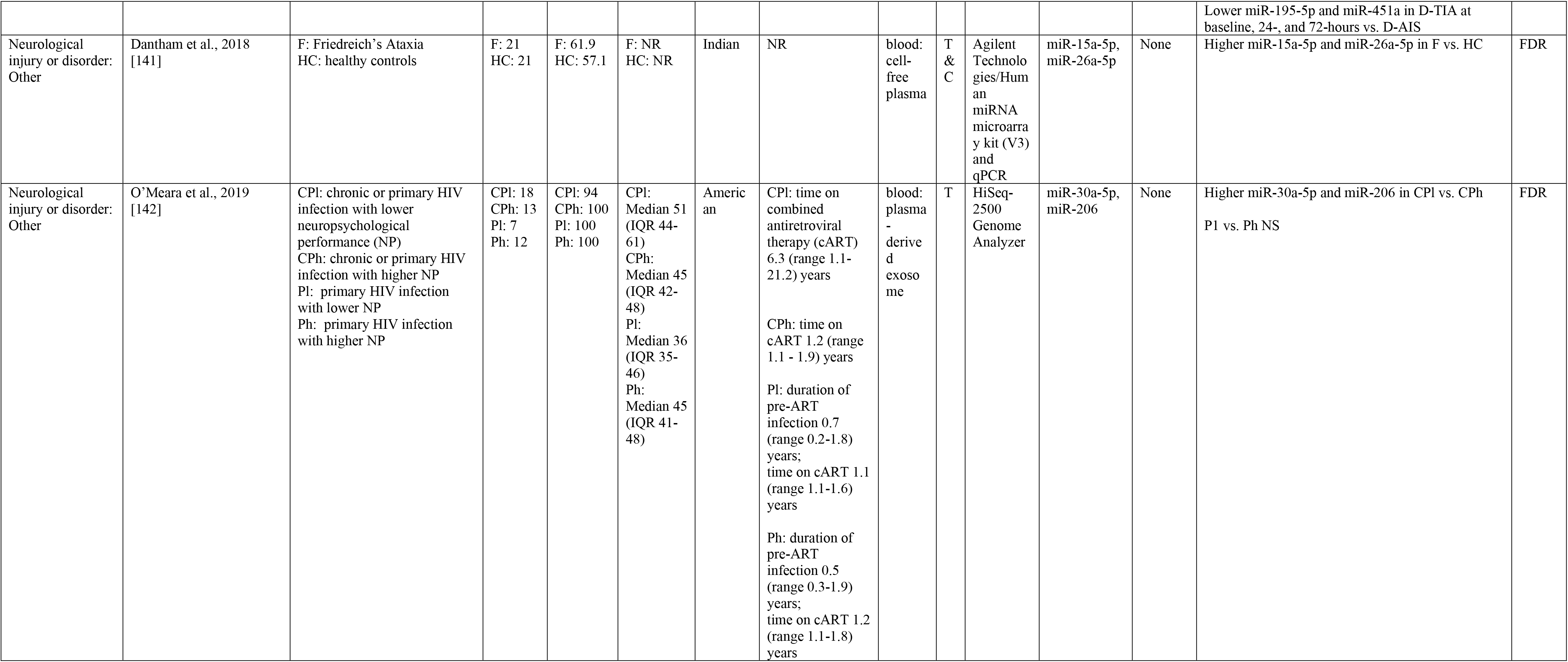
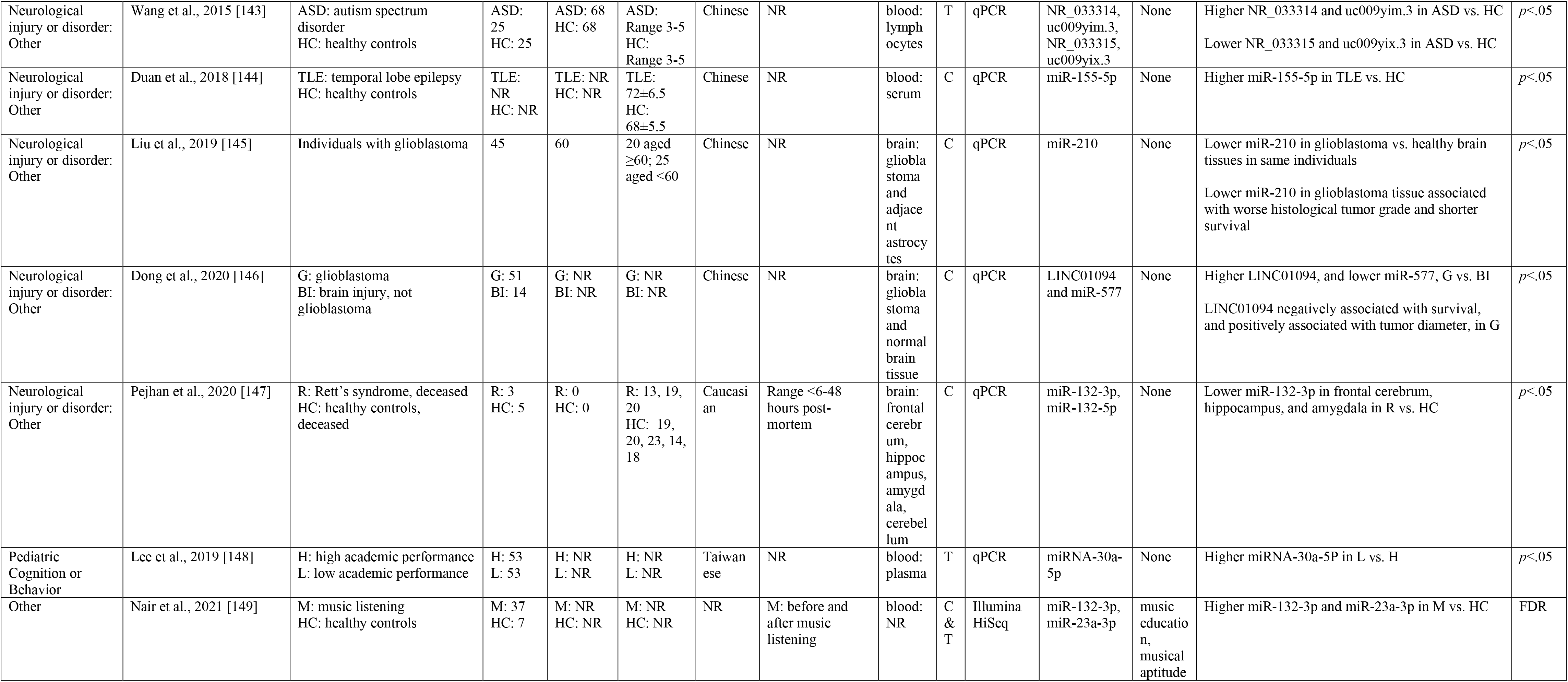
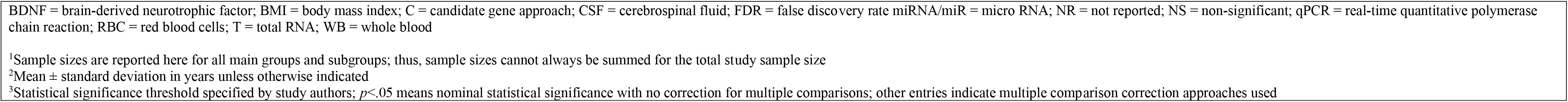
Non-Coding RNA Studies.

Non-coding RNAs exert their influence on the expression of another gene(s), and a single non-coding RNA can affect expression of hundreds to thousands of different genes (Selbach et al., 2008). Thus, only non-coding RNA studies that specifically mentioned BDNF in titles, abstracts, keywords, or MeSH terms would have been captured by our literature search. In addition, we only included non-coding RNA in our summary tables that were specifically mentioned by the authors to be related to *BDNF* expression. Therefore, the non-coding RNA results presented here include only a snapshot of the non-coding RNA literature that specifically “called out” *BDNF* and are likely less comprehensive than the DNAm and histone modification results.

##### 3.3.3.2. Methodological approaches

The total sample size across all non-coding RNA studies ranged from 8 to 458 participants, with one study [144] that did not report sample size. Mean sample size was 94 and median was 62. Sixty-eight percent of the 37 studies (n = 25) had a total sample size (groups summed) of < 100 participants.

The reporting of the timing of biosample collection and tissue type was variable across studies. Data on timing of biosample collection were not reported in approximately 54% of studies. Non-coding RNA were evaluated in blood in most studies (n = 26; 70%), followed by brain (n = 8; 22%), cerebrospinal fluid (n = 2; 5%), and nasal epithelium (n=1; 3%). Nearly half of the studies that used blood further specified serum (n = 12; 46%). Other specifications reported less frequently were plasma (n = 7; 27%), whole blood (n = 3; 12%), lymphocytes (n = 2; 8%), red blood cells (n = 1; 4%), and not reported (n = 1; 4%). All studies that extracted DNA from brain tissue specified the brain region from which tissue was collected.

Quantification of non-coding RNA was performed using real-time quantitative polymerase chain reaction (qPCR) in all but two studies. Four of the studies used a second platform (microarray and methyl-sequencing) alongside qPCR [Agilent Technologies/Human miRNA microarray kit V3, Affymetrix U133A microarray, Illumina Hiseq 2000, and Illumina Hiseq 2500 high-throughput sequencing (miRNA-seq)]. The two studies that did not use qPCR used a methyl-sequencing approach (Illumina HiSeq and HiSeq-2500 Genome Analyzer). Thirty percent of studies reported results for total RNA (n = 11), 59% (n = 22) reported results for only candidate non-coding RNA, and 11% (n = 4) reported results for both total RNA and candidate non-coding RNA analyses. Ninety-two percent of studies examined miRNA; one study examined circular RNA; one study examined BDNF antisense, and two studies examined long non-coding RNA.

Twenty-four percent of studies (n = 9) included covariate adjustment, with the number of covariates ranging from 2 (n = 2) to 7 (n = 1). Thirty percent of studies (n = 11) applied any method to correct for multiple comparisons. Three studies [113, 122, 137] tested replication of their non-coding RNA findings relevant to BDNF in independent samples.

##### 3.3.3.3. Patterns of non-Coding RNA results

All studies reported at least one statistically significant (per author definition) association between at least one non-coding RNA related to *BDNF* expression and a brain-related phenotype. The studies reflected a mix of those reporting higher vs. lower non-coding RNA in patients relative to healthy controls or in association with disease/disordered phenotypes, with around 70% of studies reporting higher non-coding RNA associated with disease/disorder phenotypes and smaller numbers of studies reporting lower or both higher and lower non-coding RNA with disease/disordered phenotypes. Certain non-coding RNA were examined across multiple studies: miR-206 (n = 6; 16%), miR-132 (n = 6; 16%), miR30a-5p (n = 4; 11%), miR-210 (n = 3; 8%), miR-182 (n = 2; 5%), miR-212 (n = 2; 5%), miR-124 (n = 2; 5%), miR142-3p (n = 2; 5%), and miR-10b-5p (n = 2; 5%).

#### 3.3.4. Non-coding RNA studies by broad brain-belated phenotype

Results are summarized below for broad phenotypes with at least two non-coding RNA studies within them.

##### 3.3.4.1. Psychopathology: depressive disorders

Seven studies examined non-coding RNA in association with depressive disorders. Of these, three were drug treatment studies [116-119]. Study [119] examined cerebral venous thrombosis, depression, and gastrointestinal infection treated with blueberry extract. Study [115] included comparisons of groups with differing combinations of ovarian cancer and depression, and studies [113 and 114] examined individuals with depression compared to healthy controls. Five studies [113, 114, 116, 117, 118, 119] examined blood, including whole blood, serum, red blood cells, or plasma. Tissue type was not reported in one study [115]. All studies used a candidate approach, while studies [113 and 115] used a total RNA approach as well. Two studies adjusted for covariates [113 and 118] and two studies corrected for multiple comparisons [113 and 116]. miRNA-132 was higher in individuals with depression vs. healthy controls across two different studies [114 and 117].

##### 3.3.4.2. Psychopathology: schizophrenia

Three studies examined non-coding RNA in association with schizophrenia. In two studies non-coding RNA was examined in blood, including whole blood [121] and serum [122] and in one study in brain [123]. Two studies [121 and 123] used a candidate approach, while study [122] used a total RNA approach. One study [121] adjusted for covariates and two studies [121 and 122] corrected for multiple comparisons. miRNA-125 was higher in individuals with schizophrenia vs. healthy controls across two different studies [121 and 123].

##### 3.3.4.3. Neurological injury or disorder: dementia and mild cognitive impairment

Twelve studies examined non-coding RNA in association with dementia and mild cognitive impairment. There was a wide variation of phenotypes among the studies, including AD, Huntington’s disease, frontotemporal lobar degeneration, and MCI. Studies examined non-coding RNA in blood serum or plasma [125, 128, 129, 130, 131, 133] and brain [127, 134, 135, 136]. Two studies examined CSF [126 and 128] and one examined nasal epithelium [132]. Two thirds of the studies used a candidate approach [125, 126, 128, 129, 130, 131, 132, 133] and the other third used a total RNA approach [127, 134, 135, 136]. Two studies adjusted for covariates [129 and 136] and three studies corrected for multiple comparisons [127, 134, 135]. miRNA-206 and miRNA-132 showed mixed higher vs. lower results in disease/disordered phenotypes across several studies. miRNA-10b-5p was higher in individuals with Huntington’s disease vs. healthy controls across two studies [134 and 135].

##### 3.3.4.4. Neurological injury or disorder: stroke

Four studies examined non-coding RNA in association with stroke. All studies examined non-coding RNA from blood; studies [137, 138, 140] analyzed serum and study [139] analyzed whole blood. Two used a candidate approach [138 and 140] and two used a total RNA approach [137 and 139]. One study [137] adjusted for covariates and one study [139] corrected for multiple comparisons.

##### 3.3.4.5. Neurological injury or disorder: other

Seven studies examined non-coding RNA in association with other neurological injuries or disorders. Phenotypes across the studies ranged from Friedrich’s Ataxia [141], chronic or primary HIV with higher or lower neuropsychological performance [142], autism spectrum disorder [143], temporal lobe epilepsy [144], glioblastoma [145 and 146], and Rett’s syndrome [147]. In four studies [141, 142, 143, 144] non-coding RNA was examined in blood, including plasma, lymphocytes, and serum, and in three studies [145, 146, 147] in brain. Four studies used a candidate approach [144, 145, 146, 147], two studies used a total RNA approach [142, 143], and one study used both candidate and total RNA approaches [141]. None of the studies adjusted for covariates and three corrected for multiple comparisons [141, 142, 149]. None of the non-coding RNA examined had significant results across multiple studies.

#### 3.3.5 Transdiagnostic overview of histone modification studies

##### 3.3.5.1. Summary table

Details of the four histone modification studies are summarized in Table 16. The directions of statistically significant associations (e.g., lower H3 acetylation; positive or negative association) are specified, non-significant associations are reported as “NS”, results are summarized after covariate adjustment when applicable (see “Covariates” column), and statistical significance is consistent with the study-specific approach and authors’ definition of “significance”—regardless of adjustment for multiple comparisons.

**Table 16:**
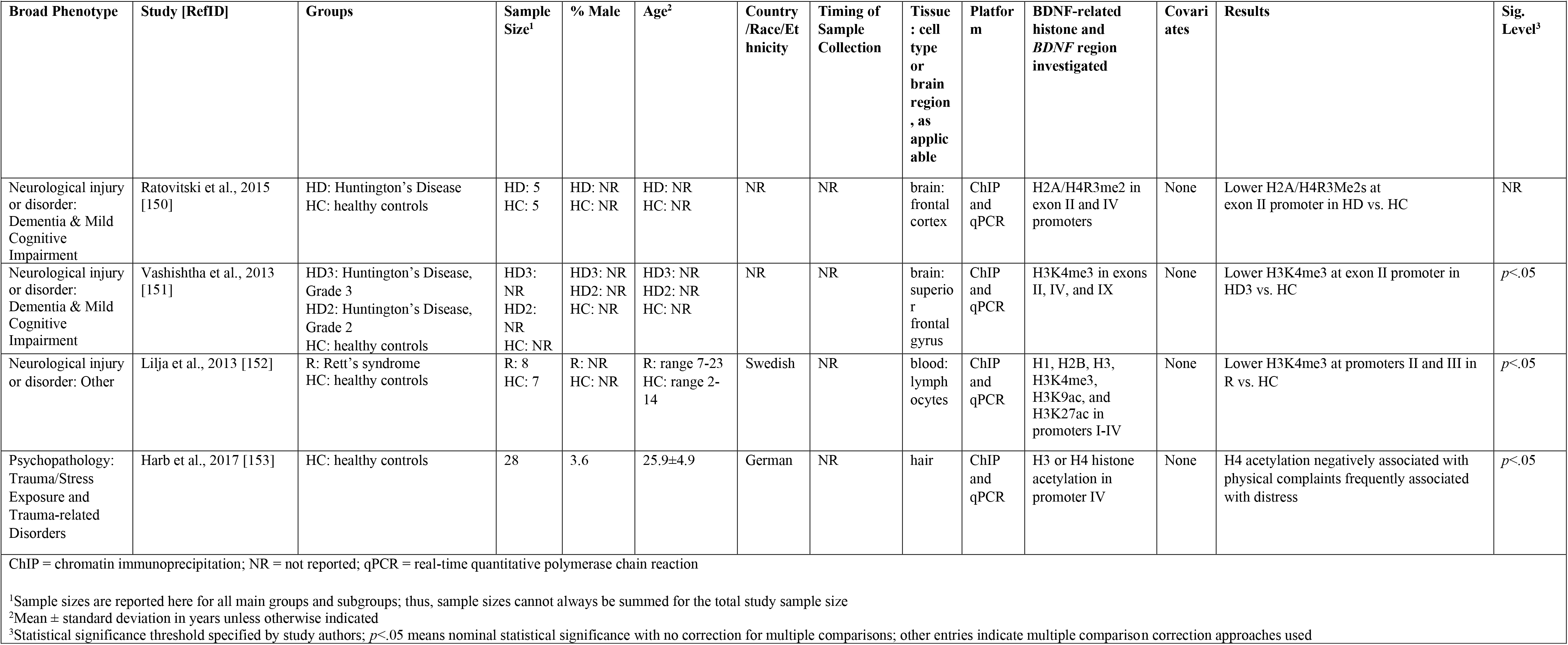
Histone Modification Studies.

| Broad Phenotype | Study [RefID] | Groups | Sample Size<sup>1</sup> | % Male | Age<sup>2</sup> | Country /Race/Ethnicity | Timing of Sample Collection | Tissue : cell type or brain region , as applicable | Platform | BDNF-related histone and *BDNF* region investigated | Covariates | Results | Sig. Level<sup>3</sup> |
| --- | --- | --- | --- | --- | --- | --- | --- | --- | --- | --- | --- | --- | --- |
| Neurological injury or disorder: Dementia & Mild Cognitive Impairment | Ratovitski et al., 2015 [150] | HD: Huntington's Disease<br>HC: healthy controls | HD: 5<br>HC: 5 | HD: NR<br>HC: NR | HD: NR<br>HC: NR | NR | NR | brain: frontal cortex | ChIP and qPCR | H2A/H4R3me2 in exon II and IV promoters | None | Lower H2A/H4R3Me2s at exon II promoter in HD vs. HC | NR |
| Neurological injury or disorder: Dementia & Mild Cognitive Impairment | Vashishtha et al., 2013 [151] | HD3: Huntington's Disease, Grade 3<br>HD2: Huntington's Disease, Grade 2<br>HC: healthy controls | HD3: NR<br>HD2: NR<br>HC: NR | HD3: NR<br>HD2: NR<br>HC: NR | HD3: NR<br>HD2: NR<br>HC: NR | NR | NR | brain: superior frontal gyrus | ChIP and qPCR | H3K4me3 in exons II, IV, and IX | None | Lower H3K4me3 at exon II promoter in HD3 vs. HC | $p<.05$ |
| Neurological injury or disorder: Other | Lilja et al., 2013 [152] | R: Rett's syndrome<br>HC: healthy controls | R: 8<br>HC: 7 | R: NR<br>HC: NR | R: range 7-23<br>HC: range 2-14 | Swedish | NR | blood: lymphocytes | ChIP and qPCR | H1, H2B, H3, H3K4me3, H3K9ac, and H3K27ac in promoters I-IV | None | Lower H3K4me3 at promoters II and III in R vs. HC | $p<.05$ |
| Psychopathology: Trauma/Stress Exposure and Trauma-related Disorders | Harb et al., 2017 [153] | HC: healthy controls | 28 | 3.6 | 25.9±4.9 | German | NR | hair | ChIP and qPCR | H3 or H4 histone acetylation in promoter IV | None | H4 acetylation negatively associated with physical complaints frequently associated with distress | $p<.05$ |

ChIP = chromatin immunoprecipitation; NR = not reported; qPCR = real-time quantitative polymerase chain reaction
<sup>1</sup>Sample sizes are reported here for all main groups and subgroups; thus, sample sizes cannot always be summed for the total study sample size
<sup>2</sup>Mean ± standard deviation in years unless otherwise indicated
<sup>3</sup>Statistical significance threshold specified by study authors; $p < .05$ means nominal statistical significance with no correction for multiple comparisons; other entries indicate multiple comparison correction approaches used

##### 3.3.5.2. Methodological approaches

The total sample size ranged from 10 to 28 participants, with one study [151] that did not report sample size. Mean sample size was 18 and median was 15. Limited demographic information about the samples was reported. Timing of biosample collection was not reported in any studies. Histone modifications were evaluated in brain in two studies and in lymphocytes and hair in the other studies, respectively. Quantification of histone modifications was performed using chromatin immunoprecipitation (ChIP) and qPCR in all studies. No studies included covariate adjustment. One study [150] did not specify their statistical analysis and the other three did not correct for multiple comparisons. No studies tested for replication in independent samples.

##### 3.3.5.3. Patterns of Histone Modification results

All four studies reported at least one statistically significant (per author definition) association between at least one histone modification and a brain-related phenotype. All four studies reported lower histone modification occupancy in association with disease/disorder phenotypes. Consistent with the DNAm studies, histone modifications in the exon IV promoter were examined most frequently.

## 4. Discussion

The present work aimed to systematically review studies investigating *BDNF* epigenomic modifications in association with brain-related phenotypes in humans. Our transdiagnostic approach, comprehensive review of methodological approaches used, and creation of an open access web-based application allowing interactive visualization of CpG sites investigated across all DNAm studies for which these data were available, will inform the design of future epigenomic studies of *BDNF*.

### 4.1. Key methodological approach findings and recommendations

A significant contribution of this systematic review is the extraction and summary of methodological details from all studies. We now summarize key findings related to methodological approach and provide recommendations for future epigenomic research, acknowledging that many of these recommendations have been provided previously and in much greater detail by others, cited below.

#### 4.1.1. Sample size and statistical power

Sample size was variable by epigenomic modification examined, with mean and median total sample sizes as follows: DNAm studies, mean = 283, median = 131; non-coding RNA studies, mean = 94, median = 62; histone modification studies, mean = 18, median = 15. Multiple epigenomic modifications were examined in the vast majority of studies; for example, looking only at DNAm studies captured in the *BDNF* DNA Methylation Map, the number of comparisons ranged from 1 to 231 in candidate gene studies and >450,000 in EWASs, creating a major multiple testing burden in some cases. Adequate statistical power is paramount in epigenomic study design, but epigenomic power calculations can be complex as they depend on hypothetical clinically or scientifically meaningful differences that can be difficult to predict and might vary by the specific epigenomic modification, tissue, and phenotype of interest. In general, however, complex phenotype-epigenomic modification associations are expected to involve small effect sizes (e.g., mean difference of 2-5%) (Heinsberg et al., 2021c). While adequate sample sizes for candidate gene studies can vary greatly based on the number of targets examined, the latest literature on the Infinium EPIC chip recommends at least 500 cases and 500 controls to achieve >80% power at 81% of CpGs sites to detect a mean DNAm difference of 2% at a genome-wide significance threshold of 9×10^-8^ (Mansell et al., 2019). While these sample sizes are more attainable than ever before as the cost of epigenome-wide data collection has decreased over recent years, only two of the twelve EWASs (16.7%) identified by this systematic review had a sample size >1000 [66, 69]. While post hoc power calculations are not meaningful (Heinsberg and Weeks, 2022), we believe that most studies captured by this systematic review were underpowered. Paired with the lack of replication of findings, the results should be interpreted with caution. The complexities of epigenomic study power calculations are discussed in detail elsewhere (Mansell et al., 2019; Rakyan et al., 2011; Tsai and Bell, 2015).

#### 4.1.2. Timing of biosample collection

Because epigenomic modifications are dynamic, the timing of biosample collection is a key consideration in interpreting epigenomic results in brain-related phenotypes. For example, there are likely differences in epigenomic modifications by chronicity for evolving conditions such as dementia and stroke, time since exposure (e.g., trauma, clinical event), and time relative to treatment (e.g., medication initiation). Despite its importance, data on timing of biosample collection were not reported in approximately 40% of DNAm studies, 54% of the non-coding RNA studies, and all histone modification studies. In future studies, we strongly recommend providing data regarding timing of biosample collection in relation to condition onset or duration, exposure (e.g., trauma), and/or intervention (e.g., psychotropic medication initiation), as applicable.

#### 4.1.3. Tissue type and cell type heterogeneity

Among the DNAm studies captured here, 79% extracted DNA from blood, with brain, saliva, buccal cells, umbilical cord blood, placenta, muscle, spermatozoa, and cerebrospinal fluid examined in a small number of studies. More than half of DNAm studies that extracted DNA from blood used whole blood, whereas a minority extracted it from specified cell types within blood (e.g. leukocytes, mononuclear cells). Among DNAm studies in which DNA was extracted from mixed cell types from any tissue, only about 15% adjusted for cell type heterogeneity (i.e., cell type proportion differences).

Cell type heterogeneity is an important consideration in epigenomic studies, and particularly so in DNAm studies. Specifically, because DNAm is cell type (and therefore tissue) specific, it can confound associations between a CpG site and phenotype if unaccounted for (e.g., researchers may think that there is a DNAm-disease association when, in reality, there is a cell type proportion-disease association as observed elsewhere (Liu et al., 2013)). While alterations in cell type proportion may be potentially quite clinically useful (assuming it is truly specific to a disease), these associations do not reflect disease-associated alterations of DNAm, creating the potential for misguided studies of bio-mechanism. Therefore, in line with recent papers (Heinsberg et al., 2021c, 2021a, 2021b; Qi and Teschendorff, 2022; Treble-Barna et al., 2021), we recommend that, when possible, analyses be performed both *adjusted* (to gain a greater biological understanding of the disease) and *unadjusted* (to identify potential clinical biomarkers for the disease) for cell type heterogeneity. This is because both approaches hold great potential value but using one without the other fails to provide a complete understanding of the data.

While we acknowledge that currently this cannot be readily accomplished in candidate gene studies, opportunities for deconvolution of cell types using pyrosequencing techniques (which would be more amenable in candidate gene studies) have been proposed in a recent proof of concept paper (Schmidt et al., 2020). While tissue and cell type may be less critical for non-coding RNA studies given that miRNA, the most studied non-coding RNA in included studies, demonstrates low cell-type specificity (Landgraf et al., 2007; Yuan et al., 2018), additional evidence is needed to define the correlation between circulating non-coding RNA and non-coding RNA in brain (Yuan et al., 2018).

Finally, because epigenomic modifications are tissue-specific and reflective of the local environment of each cell type, and examination of brain tissue is not feasible in many brain-related phenotypes, determining which types of tissue biosamples serve as effective proxies for the brain environment is an important methodological consideration for epigenomic studies. Therefore, examining the concordance between peripheral tissue and brain DNAm profiles (Hannon et al., 2015), prioritizing examination of CpGs within biological systems that are known to associate with variation in post-mortem brain samples (Barker et al., 2019; Hannon et al., 2015), and targeting CpGs with underlying genetic influence (i.e., methylation quantitative trait loci [mQTLs]) may increase the relevance of DNAm studies in peripheral tissues to brain-related phenotypes.

#### 4.1.4. Candidate gene/non-coding RNA vs. EWAS/total RNA approaches

Eighty-nine percent of DNAm studies used a candidate gene approach, while the remaining studies used EWAS or combined EWAS and candidate approaches. This preponderance of candidate studies, however, may be biased by the fact that EWASs that, by definition, examined the *BDNF* gene but did not call out BDNF in titles, abstracts, keywords, or MeSH terms would not have been identified by our literature search strategy and, therefore, were not included in the present review. Nevertheless, the much larger number of studies using a candidate gene approach is in line with the historical popularity of this approach in psychiatry and neuroscience. Unfortunately, candidate gene studies are now recognized to often be vastly underpowered, likely contributing in part to overestimates of effect size and low reproducibility of results (Button et al., 2013; Hyman and Krystal, 2018). Recent large-scale genome-wide association studies have failed to replicate some of the *BDNF* genetic associations from earlier candidate gene studies (Border et al., 2019; Johnson et al., 2017). Although EWASs are several years behind, emerging evidence indicates similar replication issues for candidate DNAm studies in brain-related phenotypes (Shields et al., 2021; Sumner et al., 2022; Wang et al., 2021). Consistent with the recent recommendations of the National Institute of Mental Health Council Workgroup on Genomics (Hyman and Krystal, 2018), we recommend that candidate DNAm studies should be abandoned for EWAS approaches in most cases. When sample sizes for EWAS are insufficient, the rigor of candidate gene DNAm studies can be promoted by following ours and others’ recommendations discussed here relevant to sample size, cell type heterogeneity, statistical analysis, transparency of scientific reporting, and – perhaps most importantly – replication of findings in an independent test sample (see section 4.1.8.). Adding additional levels of analysis, such as protein concentrations or gene expression, for the gene of interest can also strengthen the validity of candidate gene DNAm results. The examination of DNAm for sets of genes enriched within pertinent biological pathways via systems biology approaches may also bolster the impact of candidate approaches.

Regarding non-coding RNA studies, about a third of studies reported results for total RNA, two thirds for only candidate non-coding RNA, and a handful reported results for both total RNA and candidate non-coding RNA analyses. A similar call has been made to extend studies of single candidate non-coding RNA to the examination of molecular pathways associated with miRNA of interest and analysis of total RNA expression profiles (Issler and Chen, 2015; Yuan et al., 2018).

#### 4.1.5. Platform

Quantification of DNAm was performed using Illumina Infinium BeadChips for all but one EWAS, and candidate studies used pyrosequencing most often. Non-coding RNA studies used qPCR in all but two studies and all histone modification studies used chromatin immunoprecipitation (ChIP) and qPCR. We do not offer recommendations regarding specific platforms for quantification of epigenomic modifications other than to note that for DNAm studies, obtaining data using an EWAS platform allows for adjustment for cell type heterogeneity, even if a candidate analysis approach is used (see section 4.1.4.). Additionally, technology and costs to generate these data evolves over time; therefore, we recommend that investigators evaluate all platforms available at the time of data collection.

#### 4.1.6. Covariate adjustment

Sixty-two percent of DNAm studies, 24% of non-coding RNA studies, and no histone modification studies included covariate adjustment. For DNAm studies in particular, evidence indicates that DNAm is associated with several demographic and lifestyle factors, including age (Jones et al., 2015), pubertal status (Thompson et al., 2018), sex (Maschietto et al., 2017), BMI (Dick et al., 2014), and smoking (Joehanes et al., 2016). Failure to adjust for these important covariates could result in model confounding. Our recommendation is to collect and adjust for demographic and lifestyle factors that have been shown to impact the epigenomic modifications under investigation, as well as to present both adjusted and non-adjusted results for cell type heterogeneity when possible as discussed above (see section 4.1.3).

#### 4.1.7. Multiple comparison correction

Fewer than half of the DNAm studies, about one third of non-coding RNA studies, and none of the histone modification studies applied any method to correct for multiple comparisons while nearly all studies included multiple tests of epigenomic modifications (e.g., multiple CpG sites, etc.). As with the issue of insufficient sample size, failure to correct for multiple comparisons increases the likelihood of Type I error, which has been implicated in the scientific replication crisis (Goodman et al., 2018; Rubin, 2021). This methodological issue makes it particularly challenging to glean what are the most robust *BDNF* epigenomic modification effects associated with brain-related phenotypes since published definitions of statistical significance can vary so widely from nominal (e.g., any *p* < .05 regardless of multiple testing) to stringent at *p* = 5.0 × 10−7. We recommend careful consideration of the many factors related to significance testing, for which there are many outstanding papers to provide guidance(Mudge et al., 2012; Rakyan et al., 2011; Rubin, 2021).

#### 4.1.8. Replication

Only two DNAm studies, three non-coding RNA studies, and no histone modification studies attempted replication of results in independent samples. Identifying results that replicate across studies is made challenging by the heterogeneity in methods discussed here but is facilitated by the *BDNF* DNA Methylation Map for DNAm studies. We recommend that replication in independent samples be attempted to increase confidence in the reproducibility of results, particularly for candidate gene or candidate non-coding RNA approaches.

#### 4.1.9. DNAm study reporting of CpG positions

One of the most surprising findings in conducting the present systematic review was the lack of transparency in scientific reporting of information pertaining to which CpG sites were investigated within each study. No consensus appears to exist regarding position information and/or nomenclature for referring to specific CpG sites within *BDNF*. Aside from when Illumina cg probe names are used, the most common approach to date appears to be labelling CpG sites as CpG1, CpG2, etc., which lacks meaning for the reader. We were able to identify exact positions of examined CpG sites for 84% of included DNAm studies; in more cases than not, however, this required significant effort searching for the pieces of information provided (e.g., DNA sequences of CpG sites and their surrounding regions, genome position ranges of CpG sites, and primer sequences) using various bioinformatic tools (e.g., UCSC Genome Browser, NCBI nucleotide blast, and CLC Genomics Workbench). The lack of transparency in reporting CpG position information further exacerbates the difficulty in comparing results between studies or attempting replication of findings. We recommend that CpG sites be referred to by their Illumina cg probe name (when applicable) and their genomic position (e.g. chr11:27,721,905) with the essential caveat that the authors specify the genome build to which the position is aligned (e.g. Genome Reference Consortium Human Build 38).

### 4.2. Key patterns of results

Despite our comprehensive literature search approach and detailed data extraction effort, the methodological heterogeneity discussed above made it challenging to identify clear patterns of robust or replicated *BDNF* epigenomic modifications in association with brain-related phenotypes. Meaningful meta-analysis was not possible given extreme heterogeneity in phenotype, tissue type, and (for DNAm studies) specific CpG sites examined; pooling across studies would have only been possible for 2-3 studies within one or two phenotypes. This inconclusive state of the literature is quite notable given that *BDNF* is, to our knowledge, the most frequently examined gene in the field of epigenomics and brain-related phenotypes and given the substantial number of articles that were included in the present review.

Though we cannot point to specific CpG sites, non-coding RNAs, or histone modifications in *BDNF* as promising biomarkers for further interrogation, we summarize below several more general patterns of results gleaned from the present review.

#### 4.2.1. DNAm studies

Eighty-five percent of DNAm studies reported at least one statistically significant (per author definition) association between *BDNF* DNAm and a brain-related phenotype. A mix of studies reported higher vs. lower DNAm in association with disease/disordered phenotypes, with a slightly higher number of results reporting higher DNAm associated with disease/disorder phenotypes. The mixed direction of the findings could reflect potential differences in gene expression by CpG location, with lower DNAm in promoter regions, but higher DNAm in the gene body, most often associated with higher gene expression (Ball et al., 2009). Also, DNAm in promoter or gene body regions that are not active in a specific tissue will not affect expression in that tissue. The slight preponderance of higher DNAm associated with disease/disorder phenotypes and many studies targeting promoter region DNAm might point to lower *BDNF* expression as a potential causal mechanism in, or consequence of, brain pathology in disordered brain-related phenotypes. Alternatively, lower *BDNF* DNAm in promoter regions, suggesting higher *BDNF* expression associated with disordered brain-related phenotypes, could be postulated to reflect an adaptive response by the brain to promote neuroplasticity or repair in response to brain pathology.

Our creation of the *BDNF* DNA Methylation Map made it possible to interactively visualize the specific positions of BDNF DNAm CpG sites investigated across all studies for which these data were available. In doing so, we observed that studies have examined CpG sites in and around all 11 *BDNF* exons and their associated promoter regions, as well as Illumina probes outside of these regions. Aside from the Illumina probes, the most frequently examined CpG sites were positioned around exon/promoter I and exons/promoters IV and VI, as well as a smaller cluster of studies around the same region in exon IX.

DNAm studies have focused on these specific regions of the *BDNF* gene for several reasons. Higher DNAm in CpG sites of promoter regions (Davidowitz, 2009; Moore et al., 2013), and in the first exon of genes (Brenet et al., 2011), is associated with repression of transcription and lower gene expression. Thus, both Illumina beadchip probes and studies of targeted CpG sites are enriched in these promoter and first exon regions. The examination of brain-related phenotypes and DNAm of exon IV, in particular, is supported by studies of cultured cortical neurons showing activity-dependent transcription of *BDNF* in exon IV (Tabuchi et al., 2000; Tao et al., 1998; Zheng and Wang, 2009). Rodent models have also shown higher DNAm of exon IV in association with lower BDNF expression in the forebrain and hippocampus (Dennis and Levitt, 2005; Lubin et al., 2008; Roth et al., 2011). As evident in the online *BDNF* DNA Methylation Map, the concentration of studies examining CpG sites in exon VI were published almost exclusively by one group of authors [13, 15-17, 24, 26-28, 92-94], though in data comprised of several cohorts of individuals with varying phenotypes. The small cluster of studies that examined CpG sites around the same region in exon IX aimed at examining DNAm potentially altered by the common rs6265 SNP, which is located at hg38 chr11:27,658,369 in exon IX. Finally, as expected, new DNAm studies often examine the same CpG sites or regions that were significantly associated with similar phenotypes in prior studies, and some of the earliest studies included in this review reported significant CpG sites in exon/promoters I [1] and IV [30].

The *BDNF* DNA Methylation Map also allowed us to reveal several “hot spots” wherein investigated CpG sites were most often significantly associated with brain-related phenotypes. The most prominent “hot spots” across all brain-related phenotypes and tissue types were in the following hg38 approximate regions: (1) chr11:27,721,905-27,723,016 in exon/promoter I; (2) chr11:27,701,549-27,701,744 in exon/promoter IV; and (3) chr11:27,700,516-27,700,187 in exon/promoter VI. Importantly, each of these “hot spots” contains between 2-5 transcription start sites, providing potential corroborating evidence for their functional relevance for gene transcription. Filtering the *BDNF* DNA Methylation Map by tissue type to view the much less frequently examined tissues than blood, a clear “hot spot” remained in exon IV, which was repeated across brain, saliva, placenta, and buccal tissues. Coloring the *BDNF* DNA Methylation Map by tissue type or broad phenotype did not reveal any clearly appreciable regions of particular significance for a certain tissue type or broad phenotype.

Finally, of the 12 EWASs, four reported significant probes in *BDNF* after epigenome-wide multiple comparison correction. Unfortunately, none of the 11 significant probes across the four studies overlapped. While they tended to cluster in exon and promoter regions, no enrichment was clearly apparent near any specific exon.

One third of studies examined DNAm in association with *BDNF* genotype, with the vast majority focused on the rs6265 SNP. Results were mixed regarding both significance and the direction of associations, providing equivocal evidence regarding whether rs6265 functions as an mQTL.

#### 4.2.2. Non-coding RNA studies

Because non-coding RNAs exert their influence on the expression of another gene(s) and single non-coding RNA can affect expression of hundreds to thousands of different genes (Selbach et al., 2008), the non-coding RNA studies included in the present review likely represent only a snapshot of the non-coding RNA literature related to *BDNF*. All studies reported at least one statistically significant (per author definition) association between at least one non-coding RNA and a brain-related phenotype, with a significant majority reporting higher non-coding RNA in disease/disordered phenotypes. The predominance of studies reporting higher *BDNF*-related non-coding RNA might suggest suppressed post-transcriptional *BDNF* expression as a potential mechanism underlying disease/disordered brain-related phenotypes.

#### 4.2.3. Histone modification studies

All four studies reported at least one statistically significant (per author definition) association between at least one histone modification and a brain-related phenotype, all of which reported lower histone modification occupancy in disease/disorder phenotypes. Histone modifications in the exon IV promoter were examined most frequently. Interestingly, the specific histone modifications negatively associated with disease/disorder phenotypes in these four studies are all associated with lower gene expression (Bannister and Kouzarides, 2011; Vashishtha et al., 2013). Thus, lower histone modification occupancy and resultant lower *BDNF* expression could be a potential mechanism underlying disease/disordered brain-related phenotypes.

### 4.3. The BDNF DNA Methylation Map as a tool for researchers

The *BDNF* DNA Methylation Map not only provides a visual synthesis of data curated through this systematic review, but it also represents a tool for the scientific community that can support the design of DNAm studies of *BDNF*. Specifically, because users can customize the figure based on their interests or access to existing data/biosamples, they can more easily see what has been examined previously, identify sites that look promising for future research, or even look up results for specific CpG sites by position or Illumina probe ID. Finally, given the cognitive benefits of interactive data visualization, we anticipate this tool will also encourage more innovative and dynamic approaches in reporting epigenomic data and scientific results (Heinsberg et al., 2022).

### 4.4. Limitations of the present review

Our literature search strategy had several limitations. The search approach may have failed to include some articles examining *BDNF* epigenomic modifications in brain-related phenotypes that were not identified by our phenotype search terms. Further, EWASs that, by definition, examined the *BDNF* gene but did not call out *BDNF* in searched terms would not have been identified by our literature search strategy and, therefore, were not included in the present review. To capture such studies, the literature search strategy would need to be inclusive of all EWAS and brain-related phenotype studies and investigators would need to manually review each study for *BDNF*-specific results. This would likely be an informative undertaking but was outside of the scope of the present review. Finally, while we are confident in the comprehensiveness of the included DNAm and histone modification studies (aside from the limitations above), in contrast, the non-coding RNA studies included likely represent only a snapshot of this literature for reasons discussed above (section 3.3.3.1.). The use of bioinformatics tools to identify all non-coding RNA that have been associated with *BDNF* expression would be required to perform a more comprehensive search.

Another notable limitation of this review is that we did not extract information from articles regarding laboratory or statistical quality control procedures beyond adjustment for cell type heterogeneity. While we considered extracting this information, reporting of information pertaining to quality control was extremely variable. Important considerations include batch effects and population stratification. Variation in laboratory and statistical quality control undoubtedly represents another significant source of heterogeneity in this field. Readers are referred to several other papers (Aryee et al., 2014; Barfield et al., 2014, 2012; Johnson et al., 2007; Michels et al., 2013; Titus et al., 2017) for guidance on these issues.

### 4.5. Conclusions

The evidence presented in this review suggests the possibility of an association between *BDNF* epigenomic modifications and brain-related phenotypes in humans. Conclusions based on the current evidence, however, are limited by extreme heterogeneity in methodological approach regarding phenotype, CpG positions examined, and tissue type, thereby limiting meta-analysis, the identification of the most robust or replicated findings, and even the direct comparison of results between studies. The literature appears to remain in its infancy. This area of study requires additional rigorous work, with consideration of the methodological recommendations we make above, to fulfill its potential to explain heterogeneity in *BDNF*-associated risk for brain-related conditions and improve our understanding of the molecular mechanisms underlying their pathogenesis.

## Data Availability

The full source code and data base for the BDNF DNA Methylation Map is available in Github: https://github.com/lwheinsberg/BDNF_DNAmMap

https://lwheinsberg.shinyapps.io/BDNF_DNAmMap/

## Registration and protocol

The protocol for this systematic review was registered with PROSPERO (CRD42020180494). The protocol may be retrieved from https://www.crd.york.ac.uk/prospero/display_record.php?RecordID=180494

## Declaration of Interest

Dr. Treble-Barna, Dr. Heinsberg, Dr. Weeks, Dr. Conley, Mr. Breazeale, and Ms. Davis reported receiving grants from the NIH during the conduct of the study. Dr. Kochanek reported receiving grants from the NIH, the U.S. Department of Defense, the Chuck Noll foundation, the Laerdal Foundation, and the Zoll Foundation during the conduct of the study and serves as an expert witness in cases related to neurocritical care and traumatic brain injury. Dr. Yeates reported receiving grant support from the Ronald and Irene Ward Chair in Pediatric Brain injury from Alberta Children’s Hospital Foundation.

## Funding

This study was supported by NICHD grant K01HD097030 (ATB) from the NIH. The content is solely the responsibility of the authors and does not necessarily represent the official views of the funder.

## Websites

Shiny app: https://lwheinsberg.shinyapps.io/BDNF_DNAmMap/

## Notes

### Clinical Protocols

https://www.crd.york.ac.uk/prospero/display_record.php?RecordID=180494

### Funding Statement

This study was supported by NICHD grant K01HD097030 (ATB) from NICHD of the NIH. The content is solely the responsibility of the authors and does not necessarily represent the official views of the funder.

### Author Declarations

The study used ONLY openly available human data that were previously published in peer-reviewed articles.

