## Appendices: Table 1a - Table 1d for "Brain-Derived Neurotrophic Factor (*BDNF*) Epigenomic Modifications and Brain-Related Phenotypes in Humans: A Systematic Review"

**Appendix A. Literature Search Strategy**

**Table 1a: PubMed search strategy**

| | |
|---|---|
| **Provider/Interface** | National Library of Medicine |
| **Database** | PubMed |
| **Date searched** | March 12, 2020; update September 17, 2021 |
| **Database update** | March 12, 2020; update September 17, 2021 |
| **Search developer(s)** | Helena M. VonVille; Andrea Ketchum; Amery Treble-Barna; Lacey W. Heinsberg; Yvette P. Conley |
| **Limit to English?** | Yes |
| **Date Range** | No limit by date |
| **Publication Types** | No limit by publication type |
| **Search filter source** | No search filter used |

| | |
|---|---|
| **Note:** All items were downloaded to EndNote. The last 3 search statements: 1) locate overlap items from both searches per the instructions of the PI; and 2) remove overlap items from each set of search results to avoid duplication. | |
| #1 | ("Brain-derived Neurotrophic Factor"[mesh:noexp] OR "Brain-Derived Neurotrophic Factor"[mesh:noexp] OR brain-derived neurotrophic factor[tiab] OR BDNF[tiab] OR val66met[tiab]) AND ((epigenetic[tiab] OR epigenetics[tiab] OR epigenomic[tiab] OR epigenomics[tiab] OR "micrornas"[mesh:noexp] OR "circulating microrna"[mesh:noexp] OR methylation[tiab] OR methylated[tiab] OR methylation[tiab] OR micro rna*[tiab] OR microrna*[tiab] OR mirna*[tiab] OR non-coding rna[tiab] OR noncoding rna[tiab]) OR (Histone*[tiab] AND (modification[tiab] OR acetylation[tiab] OR acetylated[tiab] OR deacetylation[tiab]))) NOT (("Animals"[Mesh] OR "dogs"[MeSH Terms:noexp] OR "mice"[MeSH Terms:noexp] OR "rats"[MeSH Terms:noexp] OR "zebrafish"[MeSH Terms:noexp] OR Canine[ti] OR dog[ti] OR dogs[ti] OR mice[ti] OR mouse[ti] OR rat[ti] OR rats[ti] OR zebrafish[ti] OR "Plant Proteins"[Mesh] OR plants[mesh] OR barley[ti] OR rice[ti]) NOT ("Animals"[Mesh] AND "Humans"[Mesh])) AND ("Central Nervous System Diseases"[mesh] OR "Cognitive Neuroscience"[mesh:exp] OR "Domestic Violence"[mesh] OR "Mental Disorders"[mesh] OR "Neuropsychological Tests"[mesh:exp] OR "Premature Birth"[mesh:exp] OR "Stress Disorders, Post-Traumatic"[mesh:exp] OR "Trauma, Nervous System"[mesh] OR abuse[tiab] OR abused[tiab] OR abuser[tiab] OR abusers[tiab] OR abusive[tiab] OR abusively[tiab] OR abusiveness[tiab] OR adhd[tiab] OR Adjustment Disorder*[tiab] OR adversities[tiab] OR adversity[tiab] OR Affective disorder*[tiab] OR |

| | |
|---|---|
| | aging[tiab] OR Agoraphobia[tiab] OR Alcoholism[tiab] OR alzheimer*[tiab] OR Amnesia[tiab] OR Anorexia[tiab] OR antisocial behavior[tiab] OR Anxiety[tiab] OR anxiousness[tiab] OR Aphasia[tiab] OR ASPD[tiab] OR Attachment disorder*[tiab] OR Attention Deficit [tiab] OR Avoidant Restrictive Food Intake[tiab] OR Behavior[tiab] OR Binge-Eating[tiab] OR Bipolar[tiab] OR Body Dysmorphi*[tiab] OR Borderline Personality[tiab] OR Brain cancer*[tiab] OR brain injury[tiab] OR Brain neoplasm*[tiab] OR Brain Stem cancer*[tiab] OR Brain Stem Neoplasm*[tiab] OR Brain Stem tumor*[tiab] OR Brain Stem tumour*[tiab] OR Brain tumor*[tiab] OR Brain tumour*[tiab] OR Bulimia[tiab] OR Capgras[tiab] OR Cerebellar cancer*[tiab] OR Cerebellar Neoplasm*[tiab] OR Cerebellar tumor*[tiab] OR Cerebellar tumour*[tiab] OR Cerebral Ventricle cancer*[tiab] OR Cerebral Ventricle Neoplasm*[tiab] OR Cerebral Ventricle tumor*[tiab] OR Cerebral Ventricle tumour*[tiab] OR Child Development Disorder*[tiab] OR Choroid Plexus cancer*[tiab] OR Choroid Plexus Neoplasm*[tiab] OR Choroid Plexus tumor*[tiab] OR Choroid Plexus tumour*[tiab] OR Cognition[tiab] OR cognitive[tiab] OR Communication Disorder*[tiab] OR Compulsive Personality [tiab] OR Conduct Disorder*[tiab] OR Consciousness Disorder*[tiab] OR Conversion disorder*[tiab] OR Creutzfeldt-Jakob[tiab] OR Cyclothymic[tiab] OR Delirium[tiab] OR Delusional Parasitosis[tiab] OR dementia[tiab] OR Dependent Personality[tiab] OR deployment[tiab] OR depression[tiab] OR depressive[tiab] OR deprivation[tiab] OR deprive[tiab] OR deprived[tiab] OR deprivement[tiab] OR Developmental Disabilities[tiab] OR Diabulimia[tiab] OR dissociation[tiab] OR Dissociative Disorder*[tiab] OR Dissociative Identity Disorder*[tiab] OR domestic violence[tiab] OR drug abuse[tiab] OR Dyslexia[tiab] OR dyslexic[tiab] OR Dyspareunia[tiab] OR Dyssomnias[tiab] OR early-life stress[tiab] OR Eating disorder*[tiab] OR Elimination Disorder*[tiab] OR ELS[tiab] OR emotion regulation[tiab] OR emotional abuse[tiab] OR emotional neglect[tiab] OR emotionally abused[tiab] OR emotionally neglected[tiab] OR Encopresis[tiab] OR Enuresis[tiab] OR epilepsy[tiab] OR Epilepsy[tiab] OR Epileptic[tiab] OR Erectile Dysfunction[tiab] OR Exhibitionism[tiab] OR Factitious disorder*[tiab] OR family violence[tiab] OR fetal alcohol syndrom[tiab] OR Fetishism [tiab] OR Firesetting[tiab] OR Food Addiction[tiab] OR Frontotemporal[tiab] OR Gambling[tiab] OR Gender Disorder*[tiab] OR Gender Dysphoria[tiab] OR Glioma*[tiab] OR Huntington*[tiab] OR Hypochondriasis[tiab] OR Infratentorial cancer*[tiab] OR Infratentorial Neoplasm*[tiab] OR Infratentorial tumor*[tiab] OR Infratentorial tumour*[tiab] OR Intellectual Disability[tiab] OR ischemia[tiab] OR Kluver-Bucy*[tiab] OR Learning Disabilities[tiab] OR Lewy Body*[tiab] OR maltreat[tiab] OR maltreated[tiab] OR maltreatment[tiab] OR Masochism[tiab] OR mdd[tiab] OR mental disorder*[tiab] OR mental illness[tiab] OR minDD[tiab] OR Mood disorder*[tiab] OR Morgellon*[tiab] OR Motor disorder*[tiab] OR Motor Skills |

| | |
|---|---|
| | disorder*[tiab] OR Mutism[tiab] OR negative experiences[tiab] OR neglected[tiab] OR Neonatal Abstinence Syndrome[tiab] OR Neurasthenia[tiab] OR Neurocirculatory Asthenia[tiab] OR Neurocognitive disorder*[tiab] OR Neurocytoma[tiab] OR Neurodevelopmental disorder*[tiab] OR neurological disorder*[tiab] OR neuropsychological[tiab] OR neuroses[tiab] OR Neurotic[tiab] OR Night Eating[tiab] OR Obsessive-Compulsive[tiab] OR ocd[tiab] OR Panic[tiab] OR paranoia[tiab] OR Paranoid[tiab] OR Paraphilic [tiab] OR Parasomnias[tiab] OR Parkinson*[tiab] OR Pedophilia[tiab] OR Personality Disorder*[tiab] OR phobia*[tiab] OR Phobic[tiab] OR Pica[tiab] OR Pinealoma[tiab] OR posttraumatic[tiab] OR post-traumatic[tiab] OR Premature Ejaculation[tiab] OR preterm birth[tiab] OR psychiatric[tiab] OR psychological[tiab] OR psychopathology[tiab] OR Psychoses [tiab] OR psychosis[tiab] OR psychosocial[tiab] OR psychotic[tiab] OR PTSD[tiab] OR Rumination Syndrome[tiab] OR Sadism[tiab] OR Schizoid[tiab] OR Schizophrenia[tiab] OR schizophrenic[tiab] OR sexual abuse[tiab] OR Sexual disorder*[tiab] OR Sexual Dysfunction*[tiab] OR Sleep Disorder*[tiab] OR Somatoform[tiab] OR Stereotypic Movement disorder*[tiab] OR stress[tiab] OR stressful[tiab] OR stressor[tiab] OR stroke[tiab] OR Substance Abuse[tiab] OR suicidal[tiab] OR suicide[tiab] OR Tic[tiab] OR tics[tiab] OR trauma*[tiab] OR Trichotillomania[tiab] OR Vaginismus[tiab] OR Voyeurism[tiab]) AND english[la] |
| | Update September 17, 2021 |
| #2 | ("Brain-derived Neurotrophic Factor"[mesh:noexp] OR "Brain-Derived Neurotrophic Factor"[mesh:noexp] OR brain-derived neurotrophic factor[tiab] OR BDNF[tiab] OR val66met[tiab]) AND ((epigenetic[tiab] OR epigenetics[tiab] OR epigenomic[tiab] OR epigenomics[tiab] OR "micrornas"[mesh:noexp] OR "circulating microrna"[mesh:noexp] OR methylation[tiab] OR methylated[tiab] OR methylation[tiab] OR micro rna*[tiab] OR microrna*[tiab] OR mirna*[tiab] OR non-coding rna[tiab] OR noncoding rna[tiab]) OR (Histone*[tiab] AND (modification[tiab] OR acetylation[tiab] OR acetylated[tiab] OR deacetylation[tiab]))) NOT (("Animals"[Mesh] OR "dogs"[MeSH Terms:noexp] OR "mice"[MeSH Terms:noexp] OR "rats"[MeSH Terms:noexp] OR "zebrafish"[MeSH Terms:noexp] OR Canine[ti] OR dog[ti] OR dogs[ti] OR mice[ti] OR mouse[ti] OR rat[ti] OR rats[ti] OR zebrafish[ti] OR "Plant Proteins"[Mesh] OR plants[mesh] OR barley[ti] OR rice[ti]) NOT ("Animals"[Mesh] AND "Humans"[Mesh])) AND ("Central Nervous System Diseases"[mesh] OR "Cognitive Neuroscience"[mesh:exp] OR "Domestic Violence"[mesh] OR "Mental Disorders"[mesh] OR "Neuropsychological Tests"[mesh:exp] OR "Premature Birth"[mesh:exp] OR "Stress Disorders, Post-Traumatic"[mesh:exp] OR "Trauma, Nervous System"[mesh] OR abuse[tiab] OR abused[tiab] OR abuser[tiab] OR abusers[tiab] OR |

| | |
|---|---|
| | abusive[tiab] OR abusively[tiab] OR abusiveness[tiab] OR adhd[tiab] OR Adjustment Disorder*[tiab] OR adversities[tiab] OR adversity[tiab] OR Affective disorder*[tiab] OR aging[tiab] OR Agoraphobia[tiab] OR Alcoholism[tiab] OR alzheimer*[tiab] OR Amnesia[tiab] OR Anorexia[tiab] OR antisocial behavior[tiab] OR Anxiety[tiab] OR anxiousness[tiab] OR Aphasia[tiab] OR ASPD[tiab] OR Attachment disorder*[tiab] OR Attention Deficit [tiab] OR Avoidant Restrictive Food Intake[tiab] OR Behavior[tiab] OR Binge-Eating[tiab] OR Bipolar[tiab] OR Body Dysmorphi*[tiab] OR Borderline Personality[tiab] OR Brain cancer*[tiab] OR brain injury[tiab] OR Brain neoplasm*[tiab] OR Brain Stem cancer*[tiab] OR Brain Stem Neoplasm*[tiab] OR Brain Stem tumor*[tiab] OR Brain Stem tumour*[tiab] OR Brain tumor*[tiab] OR Brain tumour*[tiab] OR Bulimia[tiab] OR Capgras[tiab] OR Cerebellar cancer*[tiab] OR Cerebellar Neoplasm*[tiab] OR Cerebellar tumor*[tiab] OR Cerebellar tumour*[tiab] OR Cerebral Ventricle cancer*[tiab] OR Cerebral Ventricle Neoplasm*[tiab] OR Cerebral Ventricle tumor*[tiab] OR Cerebral Ventricle tumour*[tiab] OR Child Development Disorder*[tiab] OR Choroid Plexus cancer*[tiab] OR Choroid Plexus Neoplasm*[tiab] OR Choroid Plexus tumor*[tiab] OR Choroid Plexus tumour*[tiab] OR Cognition[tiab] OR cognitive[tiab] OR Communication Disorder*[tiab] OR Compulsive Personality [tiab] OR Conduct Disorder*[tiab] OR Consciousness Disorder*[tiab] OR Conversion disorder*[tiab] OR Creutzfeldt-Jakob[tiab] OR Cyclothymic[tiab] OR Delirium[tiab] OR Delusional Parasitosis[tiab] OR dementia[tiab] OR Dependent Personality[tiab] OR deployment[tiab] OR depression[tiab] OR depressive[tiab] OR deprivation[tiab] OR deprive[tiab] OR deprived[tiab] OR deprivement[tiab] OR Developmental Disabilities[tiab] OR Diabulimia[tiab] OR dissociation[tiab] OR Dissociative Disorder*[tiab] OR Dissociative Identity Disorder*[tiab] OR domestic violence[tiab] OR drug abuse[tiab] OR Dyslexia[tiab] OR dyslexic[tiab] OR Dyspareunia[tiab] OR Dyssomnias[tiab] OR early-life stress[tiab] OR Eating disorder*[tiab] OR Elimination Disorder*[tiab] OR ELS[tiab] OR emotion regulation[tiab] OR emotional abuse[tiab] OR emotional neglect[tiab] OR emotionally abused[tiab] OR emotionally neglected[tiab] OR Encopresis[tiab] OR Enuresis[tiab] OR epilepsy[tiab] OR Epilepsy[tiab] OR Epileptic[tiab] OR Erectile Dysfunction[tiab] OR Exhibitionism[tiab] OR Factitious disorder*[tiab] OR family violence[tiab] OR fetal alcohol syndrom[tiab] OR Fetishism [tiab] OR Firesetting[tiab] OR Food Addiction[tiab] OR Frontotemporal[tiab] OR Gambling[tiab] OR Gender Disorder*[tiab] OR Gender Dysphoria[tiab] OR Glioma*[tiab] OR Huntington*[tiab] OR Hypochondriasis[tiab] OR Infratentorial cancer*[tiab] OR Infratentorial Neoplasm*[tiab] OR Infratentorial tumor*[tiab] OR Infratentorial tumour*[tiab] OR Intellectual Disability[tiab] OR ischemia[tiab] OR Kluver-Bucy*[tiab] OR Learning Disabilities[tiab] OR Lewy Body*[tiab] OR maltreat[tiab] OR maltreated[tiab] OR maltreatment[tiab] OR Masochism[tiab] OR |

| | |
|---|---|
| | mdd[tiab] OR mental disorder*[tiab] OR mental illness[tiab] OR minDD[tiab] OR Mood disorder*[tiab] OR Morgellon*[tiab] OR Motor disorder*[tiab] OR Motor Skills disorder*[tiab] OR Mutism[tiab] OR negative experiences[tiab] OR neglected[tiab] OR Neonatal Abstinence Syndrome[tiab] OR Neurasthenia[tiab] OR Neurocirculatory Asthenia[tiab] OR Neurocognitive disorder*[tiab] OR Neurocytoma[tiab] OR Neurodevelopmental disorder*[tiab] OR neurological disorder*[tiab] OR neuropsychological[tiab] OR neuroses[tiab] OR Neurotic[tiab] OR Night Eating[tiab] OR Obsessive-Compulsive[tiab] OR ocd[tiab] OR Panic[tiab] OR paranoia[tiab] OR Paranoid[tiab] OR Paraphilic [tiab] OR Parasomnias[tiab] OR Parkinson*[tiab] OR Pedophilia[tiab] OR Personality Disorder*[tiab] OR phobia*[tiab] OR Phobic[tiab] OR Pica[tiab] OR Pinealoma[tiab] OR posttraumatic[tiab] OR post-traumatic[tiab] OR Premature Ejaculation[tiab] OR preterm birth[tiab] OR psychiatric[tiab] OR psychological[tiab] OR psychopathology[tiab] OR Psychoses [tiab] OR psychosis[tiab] OR psychosocial[tiab] OR psychotic[tiab] OR PTSD[tiab] OR Rumination Syndrome[tiab] OR Sadism[tiab] OR Schizoid[tiab] OR Schizophrenia[tiab] OR schizophrenic[tiab] OR sexual abuse[tiab] OR Sexual disorder*[tiab] OR Sexual Dysfunction*[tiab] OR Sleep Disorder*[tiab] OR Somatoform[tiab] OR Stereotypic Movement disorder*[tiab] OR stress[tiab] OR stressful[tiab] OR stressor[tiab] OR stroke[tiab] OR Substance Abuse[tiab] OR suicidal[tiab] OR suicide[tiab] OR Tic[tiab] OR tics[tiab] OR trauma*[tiab] OR Trichotillomania[tiab] OR Vaginismus[tiab] OR Voyeurism[tiab]) AND english[la] |

**Table 1b: Embase search strategy**

| | |
|---|---|
| **Provider/Interface** | Elsevier |
| **Database** | Embase® |
| **Date searched** | April 27, 2020; update September 27, 2021 |
| **Database update** | April 27, 2020; update September 27, 2021 |
| **Search developer(s)** | Helena M. VonVille; Andrea Ketchum |
| **Limit to English?** | Yes |
| **Date Range** | No limit by date |
| **Publication Types** | In the original search: Conference abstracts, conference reviews, and conference papers were deleted post-search from EndNote library. In the updated search: limited to articles, articles in press, data papers, letters, and reviews |
| **Search filter source** | No search filter used |

| | |
|---|---|
| **Note:** In the original search, all items were downloaded to EndNote and duplicates removed. In the updated search, duplicates were removed by excluding items found in previous searches. | |
| # 1 | brain derived neurotrophic factor'/exp OR 'brain derived neurotrophic factor receptor':de OR 'brain-derived neurotrophic factor':ti,ab,kw OR 'bdnf':ti,ab,kw OR 'val66met':ti,ab,kw |
| # 2 | ('epigenetic':ti,ab,kw OR 'epigenetics':ti,ab,kw OR 'epigenomic':ti,ab,kw OR 'epigenomics':ti,ab,kw OR 'hypermethylated':ti,ab,kw OR 'hypermethylation':ti,ab,kw OR 'methylated':ti,ab,kw OR 'methylation':ti,ab,kw OR 'microrna'/exp OR 'microrna':de OR 'micro rna*':ti,ab,kw OR 'microrna*':ti,ab,kw OR 'mirna*':ti,ab,kw OR 'non-coding rna':ti,ab,kw OR 'noncoding rna':ti,ab,kw OR (('histone*':de OR 'histone*':ti,ab,kw) AND ('modification':ti,ab,kw OR 'acetylation':ti,ab,kw OR 'acetylated':ti,ab,kw OR 'deacetylation':ti,ab,kw))) |
| # 3 | #1 AND #2 |
| # 4 | #3 NOT ('animal'/exp NOT ('animal'/exp AND 'human'/exp)) |
| # 5 | ('central nervous system disease'/exp OR 'cognitive neuroscience'/exp OR 'domestic violence'/exp OR 'mental disease'/exp OR 'neuropsychological test'/exp OR 'prematurity'/exp OR 'nervous system injury'/exp OR 'abuse':ti,ab,kw OR 'abused':ti,ab,kw OR 'abuser':ti,ab,kw OR 'abusers':ti,ab,kw OR 'abusive':ti,ab,kw OR 'abusively':ti,ab,kw OR 'abusiveness':ti,ab,kw OR 'adhd':ti,ab,kw OR 'adjustment disorder*':ti,ab,kw OR 'adversities':ti,ab,kw OR 'adversity':ti,ab,kw OR 'affective disorder*':ti,ab,kw OR 'aging':ti,ab,kw OR 'agoraphobia':ti,ab,kw OR 'alcoholism':ti,ab,kw OR 'alzheimer*':ti,ab,kw OR 'amnesia':ti,ab,kw OR 'anorexia':ti,ab,kw OR 'antisocial behavior':ti,ab,kw OR 'anxiety':ti,ab,kw OR 'anxiousness':ti,ab,kw OR 'aphasia':ti,ab,kw OR 'aspd':ti,ab,kw OR 'attachment disorder*':ti,ab,kw OR 'attention deficit':ti,ab,kw OR 'avoidant restrictive food intake':ti,ab,kw OR 'behavior':ti,ab,kw OR 'binge-eating':ti,ab,kw OR 'bipolar':ti,ab,kw OR 'body dysmorphi*':ti,ab,kw OR 'borderline personality':ti,ab,kw OR 'brain cancer*':ti,ab,kw OR |

| | |
|---|---|
| | 'brain injury':ti,ab,kw OR 'brain neoplasm*':ti,ab,kw OR 'brain stem cancer*':ti,ab,kw OR 'brain stem neoplasm*':ti,ab,kw OR 'brain stem tumor*':ti,ab,kw OR 'brain stem tumour*':ti,ab,kw OR 'brain tumor*':ti,ab,kw OR 'brain tumour*':ti,ab,kw OR 'bulimia':ti,ab,kw OR 'capgras':ti,ab,kw OR 'cerebellar cancer*':ti,ab,kw OR 'cerebellar neoplasm*':ti,ab,kw OR 'cerebellar tumor*':ti,ab,kw OR 'cerebellar tumour*':ti,ab,kw OR 'cerebral ventricle cancer*':ti,ab,kw OR 'cerebral ventricle neoplasm*':ti,ab,kw OR 'cerebral ventricle tumor*':ti,ab,kw OR 'cerebral ventricle tumour*':ti,ab,kw OR 'child development disorder*':ti,ab,kw OR 'choroid plexus cancer*':ti,ab,kw OR 'choroid plexus neoplasm*':ti,ab,kw OR 'choroid plexus tumor*':ti,ab,kw OR 'choroid plexus tumour*':ti,ab,kw OR 'cognition':ti,ab,kw OR 'cognitive':ti,ab,kw OR 'communication disorder*':ti,ab,kw OR 'compulsive personality':ti,ab,kw OR 'conduct disorder*':ti,ab,kw OR 'consciousness disorder*':ti,ab,kw OR 'conversion disorder*':ti,ab,kw OR 'creutzfeldt-jakob':ti,ab,kw OR 'cyclothymic':ti,ab,kw OR 'delirium':ti,ab,kw OR 'delusional parasitosis':ti,ab,kw OR 'dementia':ti,ab,kw OR 'dependent personality':ti,ab,kw OR 'deployment':ti,ab,kw OR 'depression':ti,ab,kw OR 'depressive':ti,ab,kw OR 'deprivation':ti,ab,kw OR 'deprive':ti,ab,kw OR 'deprived':ti,ab,kw OR 'deprivement':ti,ab,kw OR 'developmental disabilities':ti,ab,kw OR 'diabulimia':ti,ab,kw OR 'dissociation':ti,ab,kw OR 'dissociative disorder*':ti,ab,kw OR 'dissociative identity disorder*':ti,ab,kw OR 'domestic violence':ti,ab,kw OR 'drug abuse':ti,ab,kw OR 'dyslexia':ti,ab,kw OR 'dyslexic':ti,ab,kw OR 'dyspareunia':ti,ab,kw OR 'dyssomnias':ti,ab,kw OR 'early-life stress':ti,ab,kw OR 'eating disorder*':ti,ab,kw OR 'elimination disorder*':ti,ab,kw OR 'els':ti,ab,kw OR 'emotion regulation':ti,ab,kw OR 'emotional abuse':ti,ab,kw OR 'emotional neglect':ti,ab,kw OR 'emotionally abused':ti,ab,kw OR 'emotionally neglected':ti,ab,kw OR 'encopresis':ti,ab,kw OR 'enuresis':ti,ab,kw OR 'epilepsy':ti,ab,kw OR 'epileptic':ti,ab,kw OR 'erectile dysfunction':ti,ab,kw OR 'exhibitionism':ti,ab,kw OR 'factitious disorder*':ti,ab,kw OR 'family violence':ti,ab,kw OR 'fetal alcohol syndrom':ti,ab,kw OR 'fetishism':ti,ab,kw OR 'firesetting':ti,ab,kw OR 'food addiction':ti,ab,kw OR 'frontotemporal':ti,ab,kw OR 'gambling':ti,ab,kw OR 'gender disorder*':ti,ab,kw OR 'gender dysphoria':ti,ab,kw OR 'glioma*':ti,ab,kw OR 'huntington*':ti,ab,kw OR 'hypochondriasis':ti,ab,kw OR 'infratentorial cancer*':ti,ab,kw OR 'infratentorial neoplasm*':ti,ab,kw OR 'infratentorial tumor*':ti,ab,kw OR 'infratentorial tumour*':ti,ab,kw OR 'intellectual disability':ti,ab,kw OR 'ischemia':ti,ab,kw OR 'kluver-bucy*':ti,ab,kw OR 'learning disabilities':ti,ab,kw OR 'lewy body*':ti,ab,kw OR 'maltreat':ti,ab,kw OR 'maltreated':ti,ab,kw OR 'maltreatment':ti,ab,kw OR 'masochism':ti,ab,kw OR 'mdd':ti,ab,kw OR 'mental disorder*':ti,ab,kw OR 'mental illness':ti,ab,kw OR 'mindd':ti,ab,kw OR 'mood disorder*':ti,ab,kw OR 'morgellon*':ti,ab,kw OR 'motor disorder*':ti,ab,kw OR 'motor skills disorder*':ti,ab,kw OR 'mutism':ti,ab,kw OR 'negative experiences':ti,ab,kw OR 'neglected':ti,ab,kw OR 'neonatal abstinence syndrome':ti,ab,kw OR 'neurasthenia':ti,ab,kw OR 'neurocirculatory asthenia':ti,ab,kw OR 'neurocognitive disorder*':ti,ab,kw OR 'neurocytoma':ti,ab,kw OR 'neurodevelopmental disorder*':ti,ab,kw OR 'neurological disorder*':ti,ab,kw OR 'neuropsychological':ti,ab,kw OR 'neuroses':ti,ab,kw OR 'neurotic':ti,ab,kw OR 'night eating':ti,ab,kw OR 'obsessive-compulsive':ti,ab,kw OR 'ocd':ti,ab,kw OR 'panic':ti,ab,kw OR 'paranoia':ti,ab,kw OR 'paranoid':ti,ab,kw OR 'paraphilic':ti,ab,kw OR 'parasomnias':ti,ab,kw OR 'parkinson*':ti,ab,kw OR 'pedophilia':ti,ab,kw OR 'personality disorder*':ti,ab,kw OR 'phobia*':ti,ab,kw OR 'phobic':ti,ab,kw OR 'pica':ti,ab,kw OR 'pinealoma':ti,ab,kw OR 'posttraumatic':ti,ab,kw OR 'post-traumatic':ti,ab,kw OR 'premature ejaculation':ti,ab,kw OR 'preterm birth':ti,ab,kw OR 'psychiatric':ti,ab,kw OR 'psychological':ti,ab,kw OR 'psychopathology':ti,ab,kw OR 'psychoses':ti,ab,kw OR 'psychosis':ti,ab,kw OR 'psychosocial':ti,ab,kw OR 'psychotic':ti,ab,kw OR 'ptsd':ti,ab,kw OR 'rumination syndrome':ti,ab,kw OR 'sadism':ti,ab,kw OR 'schizoid':ti,ab,kw OR 'schizophrenia':ti,ab,kw OR 'schizophrenic':ti,ab,kw OR 'sexual abuse':ti,ab,kw OR 'sexual disorder*':ti,ab,kw OR 'sexual dysfunction*':ti,ab,kw OR 'sleep disorder*':ti,ab,kw OR 'somatoform':ti,ab,kw OR 'stereotypic movement disorder*':ti,ab,kw OR 'stress':ti,ab,kw OR 'stressful':ti,ab,kw OR |

| | |
|---|---|
| | 'stressor':ti,ab,kw OR 'stroke':ti,ab,kw OR 'substance abuse':ti,ab,kw OR 'suicidal':ti,ab,kw OR 'suicide':ti,ab,kw OR 'tic':ti,ab,kw OR 'tics':ti,ab,kw OR 'trauma*':ti,ab,kw OR 'trichotillomania':ti,ab,kw OR 'vaginismus':ti,ab,kw OR 'voyeurism':ti,ab,kw) |
| #6 | #4 AND #5 |
| #7 | #6 AND [english]/lim |
| #8 | ('genome-wide association study'/de OR 'EWA':ti,ab,kw OR 'EWAS':ti,ab,kw OR 'epigenome-wide':ti,ab,kw OR 'genome-wide':ti,ab,kw OR 'gwa':ti,ab,kw OR 'gwas':ti,ab,kw) |
| #9 | ('circulating microRNA'/de OR 'epigenetic':ti,ab,kw OR 'epigenetics':ti,ab,kw OR 'hypermethylated':ti,ab,kw OR 'hypermethylation':ti,ab,kw OR 'methylated':ti,ab,kw OR 'methylation':ti,ab,kw OR 'micro rna*':ti,ab,kw OR 'microRNA'/exp OR 'microrna*':ti,ab,kw OR 'mirna*':ti,ab,kw OR 'methylated':ti,ab,kw OR 'methylation':ti,ab,kw OR (('histone*':de OR 'histone*':ti,ab,kw) AND ('modification':ti,ab,kw OR 'acetylation':ti,ab,kw OR 'acetylated':ti,ab,kw OR 'deacetylation':ti,ab,kw))) |
| #10 | #8 AND #9 |
| #11 | #10 NOT ('animal'/exp NOT ('animal'/exp AND 'human'/exp)) |
| #12 | ('central nervous system disease'/exp OR 'cognitive neuroscience'/exp OR 'domestic violence'/exp OR 'mental disease'/exp OR 'neuropsychological test'/exp OR 'prematurity'/exp OR 'nervous system injury'/exp OR 'abuse':ti,ab,kw OR 'abused':ti,ab,kw OR 'abuser':ti,ab,kw OR 'abusers':ti,ab,kw OR 'abusive':ti,ab,kw OR 'abusively':ti,ab,kw OR 'abusiveness':ti,ab,kw OR 'adhd':ti,ab,kw OR 'adjustment disorder*':ti,ab,kw OR 'adversities':ti,ab,kw OR 'adversity':ti,ab,kw OR 'affective disorder*':ti,ab,kw OR 'aging':ti,ab,kw OR 'agoraphobia':ti,ab,kw OR 'alcoholism':ti,ab,kw OR 'alzheimer*':ti,ab,kw OR 'amnesia':ti,ab,kw OR 'anorexia':ti,ab,kw OR 'antisocial behavior':ti,ab,kw OR 'anxiety':ti,ab,kw OR 'anxiousness':ti,ab,kw OR 'aphasia':ti,ab,kw OR 'aspd':ti,ab,kw OR 'attachment disorder*':ti,ab,kw OR 'attention deficit':ti,ab,kw OR 'avoidant restrictive food intake':ti,ab,kw OR 'behavior':ti,ab,kw OR 'binge-eating':ti,ab,kw OR 'bipolar':ti,ab,kw OR 'body dysmorphi*':ti,ab,kw OR 'borderline personality':ti,ab,kw OR 'brain cancer*':ti,ab,kw OR 'brain injury':ti,ab,kw OR 'brain neoplasm*':ti,ab,kw OR 'brain stem cancer*':ti,ab,kw OR 'brain stem neoplasm*':ti,ab,kw OR 'brain stem tumor*':ti,ab,kw OR 'brain stem tumour*':ti,ab,kw OR 'brain tumor*':ti,ab,kw OR 'brain tumour*':ti,ab,kw OR 'bulimia':ti,ab,kw OR 'capgras':ti,ab,kw OR 'cerebellar cancer*':ti,ab,kw OR 'cerebellar neoplasm*':ti,ab,kw OR 'cerebellar tumor*':ti,ab,kw OR 'cerebellar tumour*':ti,ab,kw OR 'cerebral ventricle cancer*':ti,ab,kw OR 'cerebral ventricle neoplasm*':ti,ab,kw OR 'cerebral ventricle tumor*':ti,ab,kw OR 'cerebral ventricle tumour*':ti,ab,kw OR 'child development disorder*':ti,ab,kw OR 'choroid plexus cancer*':ti,ab,kw OR 'choroid plexus neoplasm*':ti,ab,kw OR 'choroid plexus tumor*':ti,ab,kw OR 'choroid plexus tumour*':ti,ab,kw OR 'cognition':ti,ab,kw OR 'cognitive':ti,ab,kw OR 'communication disorder*':ti,ab,kw OR 'compulsive personality':ti,ab,kw OR 'conduct disorder*':ti,ab,kw OR 'consciousness disorder*':ti,ab,kw OR 'conversion disorder*':ti,ab,kw OR 'creutzfeldt-jakob':ti,ab,kw OR 'cyclothymic':ti,ab,kw OR 'delirium':ti,ab,kw OR 'delusional parasitosis':ti,ab,kw OR 'dementia':ti,ab,kw OR 'dependent personality':ti,ab,kw OR 'deployment':ti,ab,kw OR 'depression':ti,ab,kw OR 'depressive':ti,ab,kw OR 'deprivation':ti,ab,kw OR |

| | |
|---|---|
| | 'deprive':ti,ab,kw OR 'deprived':ti,ab,kw OR 'deprivement':ti,ab,kw OR 'developmental disabilities':ti,ab,kw OR 'diabulimia':ti,ab,kw OR 'dissociation':ti,ab,kw OR 'dissociative disorder*':ti,ab,kw OR 'dissociative identity disorder*':ti,ab,kw OR 'domestic violence':ti,ab,kw OR 'drug abuse':ti,ab,kw OR 'dyslexia':ti,ab,kw OR 'dyslexic':ti,ab,kw OR 'dyspareunia':ti,ab,kw OR 'dyssomnias':ti,ab,kw OR 'early-life stress':ti,ab,kw OR 'eating disorder*':ti,ab,kw OR 'elimination disorder*':ti,ab,kw OR 'els':ti,ab,kw OR 'emotion regulation':ti,ab,kw OR 'emotional abuse':ti,ab,kw OR 'emotional neglect':ti,ab,kw OR 'emotionally abused':ti,ab,kw OR 'emotionally neglected':ti,ab,kw OR 'encopresis':ti,ab,kw OR 'enuresis':ti,ab,kw OR 'epilepsy':ti,ab,kw OR 'epileptic':ti,ab,kw OR 'erectile dysfunction':ti,ab,kw OR 'exhibitionism':ti,ab,kw OR 'factitious disorder*':ti,ab,kw OR 'family violence':ti,ab,kw OR 'fetal alcohol syndrom':ti,ab,kw OR 'fetishism':ti,ab,kw OR 'firesetting':ti,ab,kw OR 'food addiction':ti,ab,kw OR 'frontotemporal':ti,ab,kw OR 'gambling':ti,ab,kw OR 'gender disorder*':ti,ab,kw OR 'gender dysphoria':ti,ab,kw OR 'glioma*':ti,ab,kw OR 'huntington*':ti,ab,kw OR 'hypochondriasis':ti,ab,kw OR 'infratentorial cancer*':ti,ab,kw OR 'infratentorial neoplasm*':ti,ab,kw OR 'infratentorial tumor*':ti,ab,kw OR 'infratentorial tumour*':ti,ab,kw OR 'intellectual disability':ti,ab,kw OR 'ischemia':ti,ab,kw OR 'kluver-bucy*':ti,ab,kw OR 'learning disabilities':ti,ab,kw OR 'lewy body*':ti,ab,kw OR 'maltreat':ti,ab,kw OR 'maltreated':ti,ab,kw OR 'maltreatment':ti,ab,kw OR 'masochism':ti,ab,kw OR 'mdd':ti,ab,kw OR 'mental disorder*':ti,ab,kw OR 'mental illness':ti,ab,kw OR 'mindd':ti,ab,kw OR 'mood disorder*':ti,ab,kw OR 'morgellon*':ti,ab,kw OR 'motor disorder*':ti,ab,kw OR 'motor skills disorder*':ti,ab,kw OR 'mutism':ti,ab,kw OR 'negative experiences':ti,ab,kw OR 'neglected':ti,ab,kw OR 'neonatal abstinence syndrome':ti,ab,kw OR 'neurasthenia':ti,ab,kw OR 'neurocirculatory asthenia':ti,ab,kw OR 'neurocognitive disorder*':ti,ab,kw OR 'neurocytoma':ti,ab,kw OR 'neurodevelopmental disorder*':ti,ab,kw OR 'neurological disorder*':ti,ab,kw OR 'neuropsychological':ti,ab,kw OR 'neuroses':ti,ab,kw OR 'neurotic':ti,ab,kw OR 'night eating':ti,ab,kw OR 'obsessive-compulsive':ti,ab,kw OR 'ocd':ti,ab,kw OR 'panic':ti,ab,kw OR 'paranoia':ti,ab,kw OR 'paranoid':ti,ab,kw OR 'paraphilic':ti,ab,kw OR 'parasomnias':ti,ab,kw OR 'parkinson*':ti,ab,kw OR 'pedophilia':ti,ab,kw OR 'personality disorder*':ti,ab,kw OR 'phobia*':ti,ab,kw OR 'phobic':ti,ab,kw OR 'pica':ti,ab,kw OR 'pinealoma':ti,ab,kw OR 'posttraumatic':ti,ab,kw OR 'post-traumatic':ti,ab,kw OR 'premature ejaculation':ti,ab,kw OR 'preterm birth':ti,ab,kw OR 'psychiatric':ti,ab,kw OR 'psychological':ti,ab,kw OR 'psychopathology':ti,ab,kw OR 'psychoses':ti,ab,kw OR 'psychosis':ti,ab,kw OR 'psychosocial':ti,ab,kw OR 'psychotic':ti,ab,kw OR 'ptsd':ti,ab,kw OR 'rumination syndrome':ti,ab,kw OR 'sadism':ti,ab,kw OR 'schizoid':ti,ab,kw OR 'schizophrenia':ti,ab,kw OR 'schizophrenic':ti,ab,kw OR 'sexual abuse':ti,ab,kw OR 'sexual disorder*':ti,ab,kw OR 'sexual dysfunction*':ti,ab,kw OR 'sleep disorder*':ti,ab,kw OR 'somatoform':ti,ab,kw OR 'stereotypic movement disorder*':ti,ab,kw OR 'stress':ti,ab,kw OR 'stressful':ti,ab,kw OR 'stressor':ti,ab,kw OR 'stroke':ti,ab,kw OR 'substance abuse':ti,ab,kw OR 'suicidal':ti,ab,kw OR 'suicide':ti,ab,kw OR 'tic':ti,ab,kw OR 'tics':ti,ab,kw OR 'trauma*':ti,ab,kw OR 'trichotillomania':ti,ab,kw OR 'vaginismus':ti,ab,kw OR 'voyeurism':ti,ab,kw) |
| #13 | #11 AND #12 |
| #14 | #13 AND [english]/lim |
| | **Update September 27, 2021** |

| | |
|---|---|
| #15 | #7 AND ([article]/lim OR [article in press]/lim OR [data papers]/lim OR [letter]/lim OR [review]/lim) |
| #16 | #14 AND ([article]/lim OR [article in press]/lim OR [data papers]/lim OR [letter]/lim OR [review]/lim) |
| #17 | 11787061:ui OR 11986135:ui OR 12135773:ui OR 12139615:ui OR 12163697:ui OR 12163698:ui OR 12399230:ui OR 12418965:ui OR 12759921:ui OR 12944423:ui OR 14501188:ui OR 14677079:ui OR 15151498:ui OR 15207239:ui OR 15247106:ui OR 15343058:ui OR 15389703:ui OR 15459182:ui OR 15485357:ui OR 15517021:ui OR 15533642:ui OR 15720206:ui OR 15867356:ui OR 15940292:ui OR 15952869:ui OR 16082431:ui OR 16170379:ui OR 16230102:ui OR 16570847:ui OR 16682435:ui OR 16728577:ui OR 16728645:ui OR 16804030:ui OR 16818640:ui OR 16839207:ui OR 16848714:ui OR 16951158:ui OR 16954426:ui OR 16972885:ui OR 17018599:ui OR 17088384:ui OR 17178896:ui OR 17268193:ui OR 17334394:ui OR 17362799:ui OR 17415710:ui OR 17460163:ui OR 17462974:ui OR 17726534:ui OR 17765733:ui OR 17850661:ui OR 17850924:ui OR 17971223:ui OR 17994099:ui OR 18022365:ui OR 18024567:ui OR 18029387:ui OR 18075316:ui OR 18084025:ui OR 18156153:ui OR 18212068:ui OR 18319075:ui OR 18370235:ui OR 18461137:ui OR 18471879:ui OR 18474104:ui OR 18485833:ui OR 18558867:ui OR 18639233:ui OR 18719369:ui OR 18759544:ui OR 18817811:ui OR 18922972:ui OR 18948693:ui OR 18987810:ui OR 19000991:ui OR 19047176:ui OR 19121517:ui OR 19124685:ui OR 19159391:ui OR 19208779:ui OR 19279142:ui OR 19279335:ui OR 19302768:ui OR 19303062:ui OR 19317649:ui OR 19322538:ui OR 19322547:ui OR 19351832:ui OR 19358877:ui OR 19412416:ui OR 19429483:ui OR 19465937:ui OR 19541429:ui OR 19550145:ui OR 19559755:ui OR 19560734:ui OR 19568875:ui OR 19571809:ui OR 19584087:ui OR 19584924:ui OR 19631266:ui OR 19633427:ui OR 19633696:ui OR 19640594:ui OR 19661838:ui OR 19684417:ui OR 19684418:ui OR 19703567:ui OR 19721723:ui OR 19741528:ui OR 19756388:ui OR 19776032:ui OR 19796384:ui OR 19807716:ui OR 19809479:ui OR 19833297:ui OR 19844740:ui OR 19845972:ui OR 19900619:ui OR 19909260:ui OR 19922134:ui OR 19923886:ui OR 19935738:ui OR 19936134:ui OR 20030625:ui OR 20034565:ui OR 20081370:ui OR 20082975:ui OR 20124482:ui OR 20126538:ui OR 20145203:ui OR 20157508:ui OR 20170730:ui OR 20188170:ui OR 20194826:ui OR 20201854:ui OR 20208569:ui OR 20224866:ui OR 20226637:ui OR 20300172:ui OR 20347265:ui OR 20351280:ui OR 20371351:ui OR 20380821:ui OR 20390474:ui OR 20404328:ui OR 20406073:ui OR 20410295:ui OR 20418067:ui OR 20451302:ui OR 20451875:ui OR 20457675:ui OR 20471694:ui OR 20505758:ui OR 20510393:ui OR 20512147:ui OR 20516477:ui OR 20553308:ui OR 20562745:ui OR 20564181:ui OR 20582289:ui OR 20583129:ui OR 20595593:ui OR 20600988:ui OR 20603005:ui OR 20614233:ui OR 20625490:ui OR 20629514:ui OR 20643939:ui OR 20644477:ui OR 20652507:ui OR 20657825:ui OR 20661424:ui OR 20686202:ui OR 20686705:ui OR 20711171:ui OR 20714900:ui OR 20725513:ui OR 20725930:ui OR 20731666:ui OR 20808944:ui OR 20818725:ui OR 20824079:ui OR 20836906:ui OR 20868653:ui OR 20874467:ui OR 20935674:ui OR 20961626:ui OR 20961626:ui OR 20964959:ui OR 20977922:ui OR 21062751:ui OR 21122584:ui OR 21141728:ui OR 21149808:ui OR 21168126:ui OR 21178384:ui OR 21179246:ui OR 21197653:ui OR 21203516:ui OR 21206489:ui OR 21210123:ui OR 21210123:ui OR 21217833:ui OR 21245904:ui OR 21251613:ui OR 21251613:ui OR 21253408:ui OR 21256147:ui OR 21261996:ui OR 21265934:ui OR 21306990:ui OR 21314941:ui OR 21320297:ui OR 21321605:ui OR 21343564:ui OR 21348783:ui OR 21358093:ui OR 21364872:ui OR 21402790:ui OR 21411439:ui OR 21427290:ui OR 21430268:ui OR 21453447:ui OR 21453505:ui OR 21455577:ui OR 21464311:ui OR 21483898:ui OR 21489983:ui OR 21497949:ui OR 21508512:ui OR 21512313:ui OR 21516289:ui OR 21525861:ui OR 21559064:ui OR 21606928:ui OR 21609203:ui OR 21615192:ui OR 21647149:ui OR |

| | |
|---|---|
| | 21677649:ui OR 21677773:ui OR 21683323:ui OR 21693556:ui OR 21706341:ui OR 21708825:ui OR 21712773:ui OR 21723895:ui OR 21727940:ui OR 21734494:ui OR 21738488:ui OR 21738743:ui OR 21802740:ui OR 21803283:ui OR 21807998:ui OR 21830278:ui OR 21867882:ui OR 21877259:ui OR 21892731:ui OR 21894111:ui OR 21894152:ui OR 21901086:ui OR 21901745:ui OR 21908516:ui OR 21912609:ui OR 21914791:ui OR 21915259:ui OR 21926974:ui OR 21928112:ui OR 21931317:ui OR 21937876:ui OR 21945389:ui OR 21946175:ui OR 21956205:ui OR 21968193:ui OR 22001921:ui OR 22002076:ui OR 22003227:ui OR 22003392:ui OR 22003392:ui OR 22012472:ui OR 22012472:ui OR 22025123:ui OR 22025602:ui OR 22032400:ui OR 22037496:ui OR 22048252:ui OR 22048458:ui OR 22054117:ui OR 22065254:ui OR 22080730:ui OR 22099692:ui OR 22102699:ui OR 22103807:ui OR 22126157:ui OR 22140101:ui OR 22161352:ui OR 22169120:ui OR 22169120:ui OR 22182936:ui OR 22184503:ui OR 22190067:ui OR 22194877:ui OR 22206861:ui OR 22207353:ui OR 22212596:ui OR 22222044:ui OR 22224195:ui OR 22232023:ui OR 22235027:ui OR 22238121:ui OR 22245218:ui OR 22247756:ui OR 22249356:ui OR 22258902:ui OR 22270829:ui OR 22270850:ui OR 22274923:ui OR 22274961:ui OR 22303378:ui OR 22343889:ui OR 22352436:ui OR 22353757:ui OR 22363459:ui OR 22367878:ui OR 22383710:ui OR 22391556:ui OR 22426120:ui OR 22451312:ui OR 22457398:ui OR 22483044:ui OR 22488895:ui OR 22497667:ui OR 22507743:ui OR 22511943:ui OR 22514556:ui OR 22533606:ui OR 22535415:ui OR 22541288:ui OR 22541296:ui OR 22543304:ui OR 22547115:ui OR 22548723:ui OR 22565234:ui OR 22567398:ui OR 22570426:ui OR 22589448:ui OR 22607505:ui OR 22614291:ui OR 22615490:ui OR 22617348:ui OR 22623900:ui OR 22632468:ui OR 22674182:ui OR 22675360:ui OR 22681167:ui OR 22687339:ui OR 22690659:ui OR 22707957:ui OR 22716958:ui OR 22722874:ui OR 22733126:ui OR 22745829:ui OR 22752237:ui OR 22752520:ui OR 22760556:ui OR 22772731:ui OR 22772731:ui OR 22776641:ui OR 22784618:ui OR 22784618:ui OR 22787434:ui OR 22792191:ui OR 22792191:ui OR 22792358:ui OR 22802923:ui OR 22823440:ui OR 22833211:ui OR 22850735:ui OR 22883350:ui OR 22887127:ui OR 22889921:ui OR 22892716:ui OR 22895706:ui OR 22899282:ui OR 22899776:ui OR 22901293:ui OR 22904366:ui OR 22909203:ui OR 22912562:ui OR 22915015:ui OR 22920183:ui OR 22923132:ui OR 22926857:ui OR 22957032:ui OR 22958471:ui OR 22961555:ui OR 22964638:ui OR 22964682:ui OR 22966037:ui OR 22982201:ui OR 22984602:ui OR 22992936:ui OR 23013629:ui OR 23016625:ui OR 23020045:ui OR 23042784:ui OR 23045325:ui OR 23062307:ui OR 23065263:ui OR 23073979:ui OR 23086283:ui OR 23086705:ui OR 23088889:ui OR 23095399:ui OR 23097332:ui OR 23098078:ui OR 23099559:ui OR 23109089:ui OR 23114813:ui OR 23125283:ui OR 23130832:ui OR 23130833:ui OR 23130836:ui OR 23134689:ui OR 23134689:ui OR 23139755:ui OR 23145961:ui OR 23149449:ui OR 23149449:ui OR 23152449:ui OR 23154643:ui OR 23157625:ui OR 23173617:ui OR 23175387:ui OR 23177740:ui OR 23182420:ui OR 23185133:ui OR 23185213:ui OR 23190615:ui OR 23201144:ui OR 23216574:ui OR 23222849:ui OR 23226047:ui OR 23236293:ui OR 23238284:ui OR 23239666:ui OR 23239946:ui OR 23240009:ui OR 23251804:ui OR 23259645:ui OR 23264780:ui OR 23264884:ui OR 23270201:ui OR 23272238:ui OR 23273101:ui OR 23276717:ui OR 23276979:ui OR 23277782:ui OR 23280964:ui OR 23288649:ui OR 23291739:ui OR 23299920:ui OR 23300739:ui OR 23314041:ui OR 23318130:ui OR 23318417:ui OR 23320132:ui OR 23326387:ui OR 23331212:ui OR 23332324:ui OR 23336521:ui OR 23341638:ui OR 23348913:ui OR 23349634:ui OR 23351786:ui OR 23351966:ui OR 23353823:ui OR 23355297:ui OR 23356558:ui OR 23356856:ui OR 23359110:ui OR 23360310:ui OR 23372024:ui OR 23372024:ui OR 23376737:ui OR 23379751:ui OR 23382145:ui OR 23387924:ui OR 23389607:ui OR 23399480:ui OR 23417712:ui OR 23417809:ui OR 23422032:ui OR 23422199:ui OR 23422958:ui OR 23428498:ui OR 23430141:ui OR 23432646:ui OR 23432717:ui OR 23447695:ui OR 23455440:ui OR 23459466:ui OR 23463236:ui OR 23467123:ui OR 23481915:ui OR 23487789:ui OR 23497937:ui OR 23498093:ui OR 23502341:ui OR 23503168:ui OR 23509941:ui OR 23511308:ui OR 23526956:ui OR 23533356:ui OR 23539621:ui OR 23547754:ui OR 23552953:ui OR 23557329:ui OR 23560301:ui OR 23565345:ui OR 23579937:ui OR 23584115:ui OR 23590724:ui OR 23593014:ui OR 23593017:ui OR 23598174:ui OR 23602987:ui OR |

| | |
|---|---|
| | 23603442:ui OR 23606027:ui OR 23608919:ui OR 23618224:ui OR 23629961:ui OR 23630272:ui OR 23631413:ui OR 23634256:ui OR 23637569:ui OR 23637704:ui OR 23639551:ui OR 23664828:ui OR 23665156:ui OR 23669348:ui OR 23670489:ui OR 23677056:ui OR 23683160:ui OR 23689617:ui OR 23689617:ui OR 23697939:ui OR 23701460:ui OR 23704927:ui OR 23707635:ui OR 23709714:ui OR 23711791:ui OR 23717324:ui OR 23720423:ui OR 23729812:ui OR 23739121:ui OR 23751776:ui OR 23752794:ui OR 23754783:ui OR 23756188:ui OR 23756378:ui OR 23756378:ui OR 23756379:ui OR 23756379:ui OR 23770587:ui OR 23774737:ui OR 23785159:ui OR 23786914:ui OR 23788696:ui OR 23796633:ui OR 23804593:ui OR 23806439:ui OR 23831425:ui OR 23834009:ui OR 23837554:ui OR 23847551:ui OR 23852853:ui OR 23857790:ui OR 23865462:ui OR 23872847:ui OR 23876774:ui OR 23884202:ui OR 23888970:ui OR 23889991:ui OR 23900692:ui OR 23904455:ui OR 23906214:ui OR 23907097:ui OR 23907119:ui OR 23910899:ui OR 23914177:ui OR 23916482:ui OR 23920159:ui OR 23925128:ui OR 23925498:ui OR 23930849:ui OR 23931339:ui OR 23934658:ui OR 23935365:ui OR 23938174:ui OR 23951366:ui OR 23951496:ui OR 23956119:ui OR 23958448:ui OR 23958917:ui OR 23964093:ui OR 23964094:ui OR 23968814:ui OR 23972692:ui OR 23973796:ui OR 23992681:ui OR 23995205:ui OR 23996309:ui OR 23999529:ui OR 24002086:ui OR 24002086:ui OR 24014427:ui OR 24026226:ui OR 24037088:ui OR 24038247:ui OR 24038819:ui OR 24041972:ui OR 24057672:ui OR 24058526:ui OR 24058735:ui OR 24071781:ui OR 24071828:ui OR 24076518:ui OR 24077665:ui OR 24078085:ui OR 24078902:ui OR 24080855:ui OR 24090360:ui OR 24101221:ui OR 24103155:ui OR 24107773:ui OR 24109432:ui OR 24109772:ui OR 24109772:ui OR 24112369:ui OR 24115114:ui OR 24115305:ui OR 24120360:ui OR 24129413:ui OR 24129413:ui OR 24132022:ui OR 24135035:ui OR 24145027:ui OR 24148570:ui OR 24151802:ui OR 24163133:ui OR 24163245:ui OR 24179125:ui OR 24196486:ui OR 24204800:ui OR 24211850:ui OR 24213247:ui OR 24214992:ui OR 24220673:ui OR 24234873:ui OR 24248060:ui OR 24252620:ui OR 24255876:ui OR 24264533:ui OR 24266366:ui OR 24283978:ui OR 24286259:ui OR 24286259:ui OR 24286388:ui OR 24290382:ui OR 24303168:ui OR 24304487:ui OR 24309249:ui OR 24323294:ui OR 24331467:ui OR 24333734:ui OR 24336737:ui OR 24337497:ui OR 24342564:ui OR 24347181:ui OR 24349040:ui OR 24354796:ui OR 24361663:ui OR 24367289:ui OR 24367640:ui OR 24371146:ui OR 24380442:ui OR 24380960:ui OR 24382160:ui OR 24382386:ui OR 24382740:ui OR 24387052:ui OR 24390222:ui OR 24399042:ui OR 24399042:ui OR 24402055:ui OR 24423287:ui OR 24434851:ui OR 24436253:ui OR 24452011:ui OR 24452678:ui OR 24464287:ui OR 24465609:ui OR 24475181:ui OR 24479385:ui OR 24491429:ui OR 24498348:ui OR 24498348:ui OR 24501229:ui OR 24516231:ui OR 24555763:ui OR 24556472:ui OR 24559763:ui OR 24566867:ui OR 24570342:ui OR 24577456:ui OR 24586208:ui OR 24586730:ui OR 24600647:ui OR 24603599:ui OR 24609082:ui OR 24612133:ui OR 24618023:ui OR 24625753:ui OR 24626631:ui OR 24636783:ui OR 24646280:ui OR 24651765:ui OR 24653661:ui OR 24655651:ui OR 24659497:ui OR 24662927:ui OR 24668484:ui OR 24670968:ui OR 24678875:ui OR 24678875:ui OR 24691403:ui OR 24694013:ui OR 24702539:ui OR 24704120:ui OR 24704572:ui OR 24705142:ui OR 24729694:ui OR 24731781:ui OR 24732804:ui OR 24737292:ui OR 24751725:ui OR 24751885:ui OR 24767015:ui OR 24767210:ui OR 24769290:ui OR 24781207:ui OR 24781529:ui OR 24788684:ui OR 24789080:ui OR 24789982:ui OR 24792117:ui OR 24795740:ui OR 24797517:ui OR 24801253:ui OR 24801751:ui OR 24804643:ui OR 24804834:ui OR 24807792:ui OR 24811782:ui OR 24812277:ui OR 24820543:ui OR 24822056:ui OR 24831017:ui OR 24833255:ui OR 24838203:ui OR 24842956:ui OR 24843883:ui OR 24865430:ui OR 24866554:ui OR 24875834:ui OR 24878765:ui OR 24880341:ui OR 24889829:ui OR 24893906:ui OR 24905038:ui OR 24910563:ui OR 24917881:ui OR 24919190:ui OR 24923441:ui OR 24929637:ui OR 24936775:ui OR 24939695:ui OR 24943344:ui OR 24945102:ui OR 24950403:ui OR 24952577:ui OR 24956270:ui OR 24968695:ui OR 24972302:ui OR 24973978:ui OR 24978894:ui OR 24980697:ui OR 24983395:ui OR 24984191:ui OR 24984191:ui OR 24994119:ui OR 24997139:ui OR 25002536:ui OR 25008768:ui OR 25008768:ui OR 25010727:ui OR 25012071:ui OR 25012449:ui OR 25016317:ui OR 25018147:ui OR 25027291:ui OR 25028501:ui OR 25029256:ui OR 25033457:ui OR |

| | |
|---|---|
| | 25034949:ui OR 25035344:ui OR 25039969:ui OR 25043306:ui OR 25043696:ui OR 25044277:ui OR 25052007:ui OR 25054579:ui OR 25054922:ui OR 25056913:ui OR 25059688:ui OR 25064009:ui OR 25065861:ui OR 25071529:ui OR 25073599:ui OR 25079072:ui OR 25080522:ui OR 25082551:ui OR 25084801:ui OR 25093848:ui OR 25100943:ui OR 25100943:ui OR 25106037:ui OR 25111603:ui OR 25128690:ui OR 25129075:ui OR 25129077:ui OR 25131541:ui OR 25131542:ui OR 25131545:ui OR 25131546:ui OR 25135956:ui OR 25135975:ui OR 25136509:ui OR 25139739:ui OR 25142710:ui OR 25142710:ui OR 25147915:ui OR 25150398:ui OR 25154414:ui OR 25154622:ui OR 25157507:ui OR 25158004:ui OR 25158004:ui OR 25165434:ui OR 25175428:ui OR 25187986:ui OR 25189356:ui OR 25189741:ui OR 25195580:ui OR 25204312:ui OR 25204312:ui OR 25206360:ui OR 25209898:ui OR 25210621:ui OR 25217125:ui OR 25217366:ui OR 25219617:ui OR 25220861:ui OR 25223903:ui OR 25230881:ui OR 25241074:ui OR 25243493:ui OR 25247593:ui OR 25247593:ui OR 25250332:ui OR 25250735:ui OR 25251895:ui OR 25270639:ui OR 25276493:ui OR 25280896:ui OR 25284466:ui OR 25287548:ui OR 25287655:ui OR 25298004:ui OR 25300531:ui OR 25303747:ui OR 25303766:ui OR 25304910:ui OR 25309578:ui OR 25309581:ui OR 25316150:ui OR 25316457:ui OR 25316630:ui OR 25317338:ui OR 25317338:ui OR 25319267:ui OR 25319577:ui OR 25328870:ui OR 25336695:ui OR 25345474:ui OR 25351997:ui OR 25355443:ui OR 25360083:ui OR 25364284:ui OR 25364288:ui OR 25364290:ui OR 25364831:ui OR 25365775:ui OR 25371448:ui OR 25375882:ui OR 25378359:ui OR 25383518:ui OR 25383518:ui OR 25385582:ui OR 25387785:ui OR 25391375:ui OR 25395183:ui OR 25404168:ui OR 25406351:ui OR 25410540:ui OR 25410542:ui OR 25410543:ui OR 25415302:ui OR 25417108:ui OR 25424692:ui OR 25424713:ui OR 25427939:ui OR 25429046:ui OR 25434007:ui OR 25436676:ui OR 25437055:ui OR 25440020:ui OR 25444161:ui OR 25445077:ui OR 25450314:ui OR 25451398:ui OR 25451482:ui OR 25452147:ui OR 25470042:ui OR 25476119:ui OR 25476907:ui OR 25476907:ui OR 25479006:ui OR 25480659:ui OR 25480889:ui OR 25484875:ui OR 25496377:ui OR 25497042:ui OR 25501227:ui OR 25505691:ui OR 25522715:ui OR 25523945:ui OR 25524307:ui OR 25537884:ui OR 25537983:ui OR 25538225:ui OR 25541727:ui OR 25545759:ui OR 25545759:ui OR 25550231:ui OR 25552301:ui OR 25554495:ui OR 25568448:ui OR 25571874:ui OR 25575480:ui OR 25579139:ui OR 25579386:ui OR 25583993:ui OR 25583993:ui OR 25585531:ui OR 25587773:ui OR 25588181:ui OR 25589731:ui OR 25590632:ui OR 25590632:ui OR 25592753:ui OR 25598842:ui OR 25599223:ui OR 25600633:ui OR 25612291:ui OR 25617346:ui OR 25627160:ui OR 25640836:ui OR 25642163:ui OR 25642241:ui OR 25643298:ui OR 25646466:ui OR 25648279:ui OR 25650246:ui OR 25652815:ui OR 25673968:ui OR 25677560:ui OR 25692999:ui OR 25695604:ui OR 25695604:ui OR 25701644:ui OR 25701668:ui OR 25702208:ui OR 25708826:ui OR 25710121:ui OR 25710121:ui OR 25713357:ui OR 25716985:ui OR 25734057:ui OR 25739736:ui OR 25744402:ui OR 25745553:ui OR 25749033:ui OR 25749789:ui OR 25750274:ui OR 25752754:ui OR 25755749:ui OR 25762136:ui OR 25762210:ui OR 25771167:ui OR 25779370:ui OR 25780369:ui OR 25793257:ui OR 25793259:ui OR 25796320:ui OR 25798107:ui OR 25801168:ui OR 25808061:ui OR 25815259:ui OR 25815896:ui OR 25817407:ui OR 25818247:ui OR 25825351:ui OR 25827039:ui OR 25836028:ui OR 25838322:ui OR 25840049:ui OR 25841663:ui OR 25849321:ui OR 25849984:ui OR 25849984:ui OR 25858873:ui OR 25861810:ui OR 25864559:ui OR 25867743:ui OR 25873928:ui OR 25875334:ui OR 25878217:ui OR 25880215:ui OR 25881116:ui OR 25884492:ui OR 25884492:ui OR 25887538:ui OR 25900210:ui OR 25906782:ui OR 25906783:ui OR 25909085:ui OR 25918994:ui OR 25918994:ui OR 25919230:ui OR 25921703:ui OR 25925670:ui OR 25927346:ui OR 25928123:ui OR 25929520:ui OR 25937794:ui OR 25940437:ui OR 25941532:ui OR 25943100:ui OR 25944472:ui OR 25946384:ui OR 25953647:ui OR 25954173:ui OR 25957495:ui OR 25960086:ui OR 25960947:ui OR 25961270:ui OR 25962115:ui OR 25972179:ui OR 25974314:ui OR 25977295:ui OR 25979369:ui OR 25982659:ui OR 25988933:ui OR 25993294:ui OR 25994230:ui OR 26003740:ui OR 26003805:ui OR 26004081:ui OR 26005852:ui OR 26019538:ui OR 26023847:ui OR 26027495:ui OR 26034042:ui OR 26038090:ui OR 26040959:ui OR 26042083:ui OR 26044726:ui OR 26061800:ui OR 26074864:ui OR 26077430:ui OR 26082730:ui OR |

| | |
|---|---|
| | 26089329:ui OR 26089374:ui OR 26090903:ui OR 26108221:ui OR 26109954:ui OR 26123324:ui OR 26137221:ui OR 26147665:ui OR 26160260:ui OR 26160290:ui OR 26163462:ui OR 26163724:ui OR 26173148:ui OR 26173806:ui OR 26175754:ui OR 26184742:ui OR 26188143:ui OR 26203295:ui OR 26206863:ui OR 26216840:ui OR 26228432:ui OR 26236318:ui OR 26239289:ui OR 26239616:ui OR 26241031:ui OR 26249223:ui OR 26251247:ui OR 26275347:ui OR 26280510:ui OR 26285129:ui OR 26292618:ui OR 26313133:ui OR 26315760:ui OR 26316079:ui OR 26322052:ui OR 26325208:ui OR 26339299:ui OR 26339299:ui OR 26340055:ui OR 26351077:ui OR 26355017:ui OR 26365416:ui OR 26366233:ui OR 26366233:ui OR 26381263:ui OR 26384369:ui OR 26388941:ui OR 26401555:ui OR 26402772:ui OR 26404404:ui OR 26404710:ui OR 26407342:ui OR 26414157:ui OR 26415774:ui OR 26429811:ui OR 26433268:ui OR 26441003:ui OR 26442668:ui OR 26442668:ui OR 26450699:ui OR 26452058:ui OR 26452058:ui OR 26456533:ui OR 26457534:ui OR 26462620:ui OR 26472529:ui OR 26473610:ui OR 26484112:ui OR 26484121:ui OR 26484201:ui OR 26485544:ui OR 26491010:ui OR 26493382:ui OR 26494515:ui OR 26499864:ui OR 26502731:ui OR 26503909:ui OR 26505665:ui OR 26508088:ui OR 26508964:ui OR 26510954:ui OR 26515765:ui OR 26519441:ui OR 26527070:ui OR 26546125:ui OR 26551297:ui OR 26553366:ui OR 26556287:ui OR 26556287:ui OR 26557993:ui OR 26567340:ui OR 26574146:ui OR 26575221:ui OR 26575221:ui OR 26578794:ui OR 26583053:ui OR 26589234:ui OR 26592203:ui OR 26594218:ui OR 26602018:ui OR 26608002:ui OR 26609481:ui OR 26619357:ui OR 26619358:ui OR 26619905:ui OR 26621122:ui OR 26631489:ui OR 26632874:ui OR 26635563:ui OR 26639084:ui OR 26643952:ui OR 26649946:ui OR 26655997:ui OR 26656881:ui OR 26666968:ui OR 26669319:ui OR 26670097:ui OR 26670691:ui OR 26673150:ui OR 26690673:ui OR 26715857:ui OR 26723856:ui OR 26731791:ui OR 26734709:ui OR 26736035:ui OR 26742120:ui OR 26744350:ui OR 26746105:ui OR 26748095:ui OR 26760777:ui OR 26763658:ui OR 26774789:ui OR 26784970:ui OR 26784970:ui OR 26790970:ui OR 26792232:ui OR 26796812:ui OR 26798408:ui OR 26801497:ui OR 26803900:ui OR 26807331:ui OR 26809779:ui OR 26811207:ui OR 26813121:ui OR 26821212:ui OR 26822446:ui OR 26826707:ui OR 26827652:ui OR 26828581:ui OR 26829381:ui OR 26830004:ui OR 26834773:ui OR 26842588:ui OR 26847422:ui OR 26848854:ui OR 26851233:ui OR 26851828:ui OR 26857655:ui OR 26859583:ui OR 26861258:ui OR 26866842:ui OR 26867769:ui OR 26869977:ui OR 26870046:ui OR 26876488:ui OR 26877199:ui OR 26884981:ui OR 26889735:ui OR 26889969:ui OR 26890800:ui OR 26902949:ui OR 26909392:ui OR 26921221:ui OR 26921879:ui OR 26934681:ui OR 26937014:ui OR 26944296:ui OR 26945040:ui OR 26957081:ui OR 26963595:ui OR 26965544:ui OR 26966136:ui OR 26966390:ui OR 26973511:ui OR 26974950:ui OR 26976342:ui OR 26986767:ui OR 26997371:ui OR 27001315:ui OR 27006279:ui OR 27013344:ui OR 27013903:ui OR 27014599:ui OR 27014601:ui OR 27015559:ui OR 27015559:ui OR 27016692:ui OR 27021816:ui OR 27029739:ui OR 27038596:ui OR 27038728:ui OR 27047397:ui OR 27051467:ui OR 27057640:ui OR 27058395:ui OR 27060332:ui OR 27065321:ui OR 27066855:ui OR 27074206:ui OR 27087318:ui OR 27088078:ui OR 27089367:ui OR 27096222:ui OR 27096222:ui OR 27096366:ui OR 27096366:ui OR 27105112:ui OR 27109935:ui OR 27111133:ui OR 27113501:ui OR 27114850:ui OR 27115769:ui OR 27118839:ui OR 27120258:ui OR 27127229:ui OR 27127229:ui OR 27128683:ui OR 27150399:ui OR 27153397:ui OR 27158905:ui OR 27160082:ui OR 27165353:ui OR 27175219:ui OR 27200029:ui OR 27207465:ui OR 27208563:ui OR 27209362:ui OR 27212030:ui OR 27212896:ui OR 27213019:ui OR 27215977:ui OR 27217153:ui OR 27229519:ui OR 27229531:ui OR 27243754:ui OR 27250257:ui OR 27250540:ui OR 27252401:ui OR 27267954:ui OR 27269943:ui OR 27271857:ui OR 27286573:ui OR 27294413:ui OR 27301936:ui OR 27302240:ui OR 27302749:ui OR 27303265:ui OR 27303926:ui OR 27304417:ui OR 27310475:ui OR 27311772:ui OR 27319317:ui OR 27323309:ui OR 27328823:ui OR 27332624:ui OR 27349968:ui OR 27350097:ui OR 27350810:ui OR 27350810:ui OR 27358065:ui OR 27358653:ui OR 27366929:ui OR 27367046:ui OR 27378372:ui OR 27380887:ui OR 27381812:ui OR 27382982:ui OR 27386500:ui OR 27388583:ui OR 27390218:ui OR 27393146:ui OR 27411884:ui OR 27416614:ui OR 27417856:ui OR 27423554:ui OR 27423601:ui OR 27424800:ui OR 27425608:ui OR |

| | |
|---|---|
| | 27426126:ui OR 27440233:ui OR 27440388:ui OR 27453791:ui OR 27454463:ui OR 27454463:ui OR 27461410:ui OR 27464224:ui OR 27466229:ui OR 27466272:ui OR 27468947:ui OR 27469238:ui OR 27478543:ui OR 27493699:ui OR 27498152:ui OR 27509014:ui OR 27510902:ui OR 27512403:ui OR 27516062:ui OR 27518140:ui OR 27525637:ui OR 27526315:ui OR 27527274:ui OR 27529193:ui OR 27531226:ui OR 27533309:ui OR 27542345:ui OR 27544872:ui OR 27545218:ui OR 27551116:ui OR 27554040:ui OR 27554040:ui OR 27556923:ui OR 27556923:ui OR 27571009:ui OR 27572077:ui OR 27573688:ui OR 27582743:ui OR 27585064:ui OR 27585440:ui OR 27585440:ui OR 27585862:ui OR 27586304:ui OR 27587204:ui OR 27588021:ui OR 27589228:ui OR 27595594:ui OR 27596955:ui OR 27601590:ui OR 27602171:ui OR 27602171:ui OR 27603714:ui OR 27610205:ui OR 27617302:ui OR 27620842:ui OR 27623749:ui OR 27628303:ui OR 27629381:ui OR 27630645:ui OR 27634380:ui OR 27643477:ui OR 27643478:ui OR 27650867:ui OR 27653855:ui OR 27657733:ui OR 27662297:ui OR 27663196:ui OR 27667199:ui OR 27672717:ui OR 27681803:ui OR 27687908:ui OR 27688075:ui OR 27709425:ui OR 27712141:ui OR 27720004:ui OR 27733502:ui OR 27759212:ui OR 27759945:ui OR 27762073:ui OR 27773727:ui OR 27774408:ui OR 27775175:ui OR 27780202:ui OR 27799474:ui OR 27800026:ui OR 27806357:ui OR 27816213:ui OR 27822313:ui OR 27822318:ui OR 27824361:ui OR 27826092:ui OR 27829866:ui OR 27837582:ui OR 27843151:ui OR 27843151:ui OR 27862945:ui OR 27863252:ui OR 27864917:ui OR 27865774:ui OR 27865783:ui OR 27866097:ui OR 27867363:ui OR 27870405:ui OR 27874848:ui OR 27875544:ui OR 27877179:ui OR 27880996:ui OR 27883060:ui OR 27884142:ui OR 27884142:ui OR 27886132:ui OR 27886174:ui OR 27886370:ui OR 27903967:ui OR 27911095:ui OR 27912315:ui OR 27913161:ui OR 27918238:ui OR 27922636:ui OR 27922854:ui OR 27933312:ui OR 27935556:ui OR 27936112:ui OR 27939088:ui OR 27968732:ui OR 27973581:ui OR 27974237:ui OR 27980682:ui OR 27982725:ui OR 27983595:ui OR 27992891:ui OR 27993893:ui OR 27997041:ui OR 27997543:ui OR 27997543:ui OR 27998510:ui OR 28011714:ui OR 28018143:ui OR 28018572:ui OR 28031556:ui OR 28032389:ui OR 28035180:ui OR 28036297:ui OR 28044061:ui OR 28045465:ui OR 28053049:ui OR 28056824:ui OR 28057210:ui OR 28058257:ui OR 28058582:ui OR 28063933:ui OR 28065402:ui OR 28067462:ui OR 28070760:ui OR 28078450:ui OR 28082079:ui OR 28083869:ui OR 28089213:ui OR 28090150:ui OR 28094813:ui OR 28103696:ui OR 28114032:ui OR 28117656:ui OR 28117840:ui OR 28119721:ui OR 28123152:ui OR 28129888:ui OR 28137310:ui OR 28140666:ui OR 28149334:ui OR 28165855:ui OR 28172975:ui OR 28173571:ui OR 28186131:ui OR 28194101:ui OR 28195572:ui OR 28203683:ui OR 28210622:ui OR 28220432:ui OR 28220606:ui OR 28223918:ui OR 28225026:ui OR 28229104:ui OR 28230862:ui OR 28231797:ui OR 28235564:ui OR 28241754:ui OR 28241794:ui OR 28242325:ui OR 28246044:ui OR 28248924:ui OR 28254385:ui OR 28255110:ui OR 28263309:ui OR 28270234:ui OR 28272318:ui OR 28276657:ui OR 28289216:ui OR 28289275:ui OR 28289478:ui OR 28291257:ui OR 28293299:ui OR 28293733:ui OR 28300138:ui OR 28302556:ui OR 28303968:ui OR 28306543:ui OR 28314689:ui OR 28319850:ui OR 28320757:ui OR 28325913:ui OR 28334972:ui OR 28335437:ui OR 28335457:ui OR 28343758:ui OR 28346445:ui OR 28346830:ui OR 28349893:ui OR 28357254:ui OR 28358823:ui OR 28359301:ui OR 28360962:ui OR 28364364:ui OR 28373915:ui OR 28374847:ui OR 28382934:ui OR 28387675:ui OR 28387726:ui OR 28388012:ui OR 28392843:ui OR 28401334:ui OR 28401839:ui OR 28412501:ui OR 28412600:ui OR 28418861:ui OR 28421459:ui OR 28421636:ui OR 28428831:ui OR 28430931:ui OR 28432360:ui OR 28439722:ui OR 28441426:ui OR 28444219:ui OR 28458684:ui OR 28459998:ui OR 28460316:ui OR 28473755:ui OR 28475857:ui OR 28475860:ui OR 28476540:ui OR 28477725:ui OR 28482637:ui OR 28485726:ui OR 28493018:ui OR 28496451:ui OR 28506242:ui OR 28512237:ui OR 28515796:ui OR 28515798:ui OR 28516910:ui OR 28520754:ui OR 28523543:ui OR 28523545:ui OR 28523546:ui OR 28523547:ui OR 28523549:ui OR 28523553:ui OR 28523554:ui OR 28523562:ui OR 28528868:ui OR 28533441:ui OR 28540675:ui OR 28540928:ui OR 28545823:ui OR 28554408:ui OR 28560252:ui OR 28560642:ui OR 28575645:ui OR 28578378:ui OR 28586827:ui OR 28587281:ui OR 28593903:ui OR 28595673:ui OR 28597562:ui OR 28601225:ui OR 28603776:ui OR 28613964:ui OR 28614626:ui OR |

| | |
|---|---|
| | 28614721:ui OR 28618313:ui OR 28620317:ui OR 28621414:ui OR 28621701:ui OR 28623522:ui OR 28624982:ui OR 28630453:ui OR 28630479:ui OR 28637423:ui OR 28645536:ui OR 28645745:ui OR 28645747:ui OR 28650998:ui OR 28653979:ui OR 28654093:ui OR 28659968:ui OR 28663050:ui OR 28666525:ui OR 28669576:ui OR 28676013:ui OR 28678593:ui OR 28680507:ui OR 28686328:ui OR 28691784:ui OR 28693600:ui OR 28696412:ui OR 28696507:ui OR 28698603:ui OR 28714605:ui OR 28731985:ui OR 28735395:ui OR 28740569:ui OR 28743056:ui OR 28747766:ui OR 28750583:ui OR 28753533:ui OR 28754344:ui OR 28761062:ui OR 28761347:ui OR 28764913:ui OR 28777307:ui OR 28785368:ui OR 28790168:ui OR 28798405:ui OR 28803920:ui OR 28805248:ui OR 28806956:ui OR 28807678:ui OR 28807811:ui OR 28809857:ui OR 28811542:ui OR 28811569:ui OR 28812058:ui OR 28812221:ui OR 28813320:ui OR 28816414:ui OR 28824374:ui OR 28824629:ui OR 28827549:ui OR 28830794:ui OR 28844656:ui OR 28848059:ui OR 28848234:ui OR 28850114:ui OR 28851441:ui OR 28859997:ui OR 28866733:ui OR 28867284:ui OR 28867940:ui OR 28869584:ui OR 28879201:ui OR 28879201:ui OR 28883893:ui OR 28887651:ui OR 28900432:ui OR 28904069:ui OR 28912427:ui OR 28914102:ui OR 28918100:ui OR 28925810:ui OR 28937692:ui OR 28938182:ui OR 28938489:ui OR 28942748:ui OR 28948711:ui OR 28955602:ui OR 28956815:ui OR 28957772:ui OR 28958479:ui OR 28960623:ui OR 28962690:ui OR 28964710:ui OR 28967789:ui OR 28972577:ui OR 28984592:ui OR 28986235:ui OR 28990704:ui OR 28993028:ui OR 29017764:ui OR 29017800:ui OR 29021802:ui OR 29026446:ui OR 29026448:ui OR 29028111:ui OR 29028941:ui OR 29030221:ui OR 29032046:ui OR 29034560:ui OR 29036598:ui OR 29042220:ui OR 29044166:ui OR 29047347:ui OR 29049855:ui OR 29051472:ui OR 29053637:ui OR 29056292:ui OR 29059379:ui OR 29061165:ui OR 29066808:ui OR 29066820:ui OR 29073110:ui OR 29082433:ui OR 29082545:ui OR 29084334:ui OR 29093763:ui OR 29098673:ui OR 29099282:ui OR 29106553:ui OR 29106556:ui OR 29118371:ui OR 29123172:ui OR 29123662:ui OR 29128257:ui OR 29131015:ui OR 29133290:ui OR 29142228:ui OR 29159506:ui OR 29163031:ui OR 29174947:ui OR 29178831:ui OR 29179742:ui OR 29180699:ui OR 29190000:ui OR 29191807:ui OR 29197086:ui OR 29215712:ui OR 29221202:ui OR 29225347:ui OR 29237550:ui OR 29243873:ui OR 29246185:ui OR 29247774:ui OR 29249830:ui OR 29257320:ui OR 29258399:ui OR 29258561:ui OR 29259014:ui OR 29263199:ui OR 29269116:ui OR 29269866:ui OR 29271864:ui OR 29272728:ui OR 29274998:ui OR 29277323:ui OR 29285792:ui OR 29288745:ui OR 29302075:ui OR 29302075:ui OR 29302221:ui OR 29305581:ui OR 29311653:ui OR 29311901:ui OR 29315108:ui OR 29317608:ui OR 29321033:ui OR 29321734:ui OR 29321829:ui OR 29324665:ui OR 29325019:ui OR 29325449:ui OR 29325450:ui OR 29325565:ui OR 29330379:ui OR 29331712:ui OR 29334322:ui OR 29342273:ui OR 29344313:ui OR 29350455:ui OR 29357833:ui OR 29360511:ui OR 29362489:ui OR 29365082:ui OR 29365082:ui OR 29374233:ui OR 29375678:ui OR 29382824:ui OR 29391398:ui OR 29394898:ui OR 29395064:ui OR 29395951:ui OR 29397389:ui OR 29399631:ui OR 29410024:ui OR 29410319:ui OR 29418072:ui OR 29419728:ui OR 29422978:ui OR 29425374:ui OR 29429989:ui OR 29430824:ui OR 29431744:ui OR 29442441:ui OR 29444334:ui OR 29444950:ui OR 29453446:ui OR 29456765:ui OR 29458840:ui OR 29464486:ui OR 29467291:ui OR 29470513:ui OR 29476661:ui OR 29478063:ui OR 29478670:ui OR 29482655:ui OR 29484035:ui OR 29492189:ui OR 29492189:ui OR 29492790:ui OR 29499969:ui OR 29500431:ui OR 29500446:ui OR 29506027:ui OR 29507413:ui OR 29512462:ui OR 29513726:ui OR 29514460:ui OR 29514626:ui OR 29520039:ui OR 29522357:ui OR 29526602:ui OR 29528196:ui OR 29532581:ui OR 29550519:ui OR 29552298:ui OR 29554300:ui OR 29555185:ui OR 29555928:ui OR 29560787:ui OR 29569031:ui OR 29579179:ui OR 29587883:ui OR 29587883:ui OR 29589131:ui OR 29594113:ui OR 29599094:ui OR 29601581:ui OR 29604450:ui OR 29605850:ui OR 29610404:ui OR 29622492:ui OR 29623448:ui OR 29625067:ui OR 29633022:ui OR 29635492:ui OR 29636708:ui OR 29649500:ui OR 29653195:ui OR 29653565:ui OR 29659581:ui OR 29667924:ui OR 29676232:ui OR 29685789:ui OR 29686828:ui OR 29689690:ui OR 29692214:ui OR 29702606:ui OR 29703944:ui OR 29704316:ui OR 29707254:ui OR 29710702:ui OR 29713391:ui OR 29736028:ui OR 29740486:ui OR 29745836:ui OR 29747567:ui OR 29748862:ui OR 29754148:ui OR |

| | |
|---|---|
| | 29760392:ui OR 29765957:ui OR 29766299:ui OR 29775682:ui OR 29777097:ui OR 29780318:ui OR 29781947:ui OR 29790956:ui OR 29790996:ui OR 29794476:ui OR 29795411:ui OR 29796118:ui OR 29796838:ui OR 29800330:ui OR 29803207:ui OR 29813122:ui OR 29844126:ui OR 29847446:ui OR 29850476:ui OR 29888306:ui OR 29888956:ui OR 29891550:ui OR 29891723:ui OR 29896231:ui OR 29896466:ui OR 29897034:ui OR 29901097:ui OR 29904359:ui OR 29920826:ui OR 29927689:ui OR 29930222:ui OR 29930742:ui OR 29931221:ui OR 29933950:ui OR 29933952:ui OR 29933955:ui OR 29935433:ui OR 29946108:ui OR 29948938:ui OR 29953671:ui OR 29955163:ui OR 29956760:ui OR 29963072:ui OR 29966198:ui OR 29976992:ui OR 29980228:ui OR 29981347:ui OR 29981347:ui OR 29982329:ui OR 29982912:ui OR 29983834:ui OR 29988321:ui OR 29997237:ui OR 29997355:ui OR 29997473:ui OR 29998287:ui OR 30001177:ui OR 30007940:ui OR 30009687:ui OR 30010719:ui OR 30010856:ui OR 30010932:ui OR 30016403:ui OR 30018734:ui OR 30019219:ui OR 30021660:ui OR 30022827:ui OR 30022974:ui OR 30030436:ui OR 30030655:ui OR 30035013:ui OR 30036399:ui OR 30036794:ui OR 30038276:ui OR 30038744:ui OR 30045751:ui OR 30050033:ui OR 30051352:ui OR 30052471:ui OR 30053126:ui OR 30053206:ui OR 30055328:ui OR 30061860:ui OR 30061930:ui OR 30065345:ui OR 30067287:ui OR 30068421:ui OR 30068429:ui OR 30071357:ui OR 30072053:ui OR 30072061:ui OR 30073537:ui OR 30075802:ui OR 30077874:ui OR 30081103:ui OR 30082643:ui OR 30083112:ui OR 30084836:ui OR 30084846:ui OR 30085031:ui OR 30089309:ui OR 30090644:ui OR 30091005:ui OR 30097719:ui OR 30099093:ui OR 30103066:ui OR 30105933:ui OR 30118972:ui OR 30134995:ui OR 30135428:ui OR 30136078:ui OR 30140277:ui OR 30144780:ui OR 30148158:ui OR 30153076:ui OR 30153567:ui OR 30157848:ui OR 30159897:ui OR 30174526:ui OR 30176915:ui OR 30178254:ui OR 30182732:ui OR 30189241:ui OR 30200279:ui OR 30205119:ui OR 30206277:ui OR 30209346:ui OR 30214456:ui OR 30214617:ui OR 30223541:ui OR 30223673:ui OR 30224877:ui OR 30226399:ui OR 30233327:ui OR 30237808:ui OR 30239726:ui OR 30239759:ui OR 30242228:ui OR 30246859:ui OR 30248838:ui OR 30250424:ui OR 30250452:ui OR 30251590:ui OR 30252044:ui OR 30252101:ui OR 30253793:ui OR 30256915:ui OR 30259378:ui OR 30264654:ui OR 30264654:ui OR 30269040:ui OR 30271706:ui OR 30287754:ui OR 30294340:ui OR 30302047:ui OR 30305647:ui OR 30305995:ui OR 30308117:ui OR 30317068:ui OR 30322087:ui OR 30326163:ui OR 30332658:ui OR 30339835:ui OR 30341348:ui OR 30343628:ui OR 30349106:ui OR 30351206:ui OR 30352697:ui OR 30353170:ui OR 30354327:ui OR 30356121:ui OR 30362655:ui OR 30364173:ui OR 30365064:ui OR 30373123:ui OR 30373123:ui OR 30373676:ui OR 30380818:ui OR 30386171:ui OR 30389222:ui OR 30392435:ui OR 30392813:ui OR 30394136:ui OR 30395198:ui OR 30396857:ui OR 30397132:ui OR 30397880:ui OR 30398472:ui OR 30399374:ui OR 30401887:ui OR 30402028:ui OR 30402861:ui OR 30403795:ui OR 30405117:ui OR 30405699:ui OR 30411855:ui OR 30412705:ui OR 30416380:ui OR 30439595:ui OR 30447004:ui OR 30451334:ui OR 30452941:ui OR 30454681:ui OR 30456986:ui OR 30457060:ui OR 30458022:ui OR 30458693:ui OR 30459647:ui OR 30463905:ui OR 30463905:ui OR 30471082:ui OR 30474186:ui OR 30480232:ui OR 30480619:ui OR 30483072:ui OR 30483149:ui OR 3048441 |
| #18 | L350236397:id OR L350247486:id OR L350277586:id OR L351122343:id OR L351308294:id OR L351425881:id OR L351442119:id OR L35148576:id OR L351489337:id OR L35175098:id OR L351858741:id OR L352054224:id OR L352079252:id OR L352324729:id OR L352456352:id OR L352497776:id OR L352568504:id OR L352775813:id OR L352790091:id OR L352847461:id OR L35314717:id OR L354058214:id OR L354160835:id OR L354249182:id OR L354341300:id OR L354382118:id OR L354438416:id OR L354520438:id OR L354667484:id OR L354728937:id OR L354769550:id OR L354853328:id OR L354944534:id OR L355113438:id OR L355153681:id OR L355155826:id OR L355189182:id OR L355386014:id OR L355458526:id OR L355596225:id OR L355715591:id OR L355767923:id OR L355824920:id OR L358066026:id OR L358071973:id OR L358095449:id OR L358102507:id OR L358201511:id OR L358243101:id OR L358260059:id OR L358298284:id OR L358325383:id OR L358436916:id |

| | |
|---|---|
| | OR L358512641:id OR L358516320:id OR L358602661:id OR L358647513:id OR L358730961:id OR L358782049:id OR L358786304:id OR L358844708:id OR L358844710:id OR L358880695:id OR L359088644:id OR L359126696:id OR L359127557:id OR L359139693:id OR L359236005:id OR L359243543:id OR L359345104:id OR L359358215:id OR L359362764:id OR L359380534:id OR L359404939:id OR L359481979:id OR L359517091:id OR L359660904:id OR L359671905:id OR L359671908:id OR L359674197:id OR L359693735:id OR L359769498:id OR L359790550:id OR L359807145:id OR L359849937:id OR L359854192:id OR L359979461:id OR L360017968:id OR L360069919:id OR L360277307:id OR L361020582:id OR L361023742:id OR L361090246:id OR L361136627:id OR L361157382:id OR L361157941:id OR L361158293:id OR L361203256:id OR L361341791:id OR L361373704:id OR L361385274:id OR L361472769:id OR L361476400:id OR L361502080:id OR L361507406:id OR L361589196:id OR L361620029:id OR L361653008:id OR L361708547:id OR L361771005:id OR L361794408:id OR L361890954:id OR L361923709:id OR L361973532:id OR L362042659:id OR L362109956:id OR L362168325:id OR L362246450:id OR L362358857:id OR L362372591:id OR L362416855:id OR L362539905:id OR L362592400:id OR L362708519:id OR L362721139:id OR L362765220:id OR L362769697:id OR L362882644:id OR L362923253:id OR L362947720:id OR L362967344:id OR L363006368:id OR L363054579:id OR L363072425:id OR L363087496:id OR L364010929:id OR L364102828:id OR L364103882:id OR L364155261:id OR L364282417:id OR L364329924:id OR L364336662:id OR L364505993:id OR L364584718:id OR L364623236:id OR L364684619:id OR L364705028:id OR L364705036:id OR L364717726:id OR L364721050:id OR L364730515:id OR L364834555:id OR L365012628:id OR L365062554:id OR L365079513:id OR L365166425:id OR L365217853:id OR L365233520:id OR L365235621:id OR L365235991:id OR L365270075:id OR L365284243:id OR L365298296:id OR L365345557:id OR L365368069:id OR L365391613:id OR L365471024:id OR L365482188:id OR L365522394:id OR L365529045:id OR L365600181:id OR L365601403:id OR L365626287:id OR L365682929:id OR L365742943:id OR L365765304:id OR L365765307:id OR L365795486:id OR L365854516:id OR L365891754:id OR L365955248:id OR L366039698:id OR L366041848:id OR L366042298:id OR L366084601:id OR L366119575:id OR L366130897:id OR L366156308:id OR L366164323:id OR L366214166:id OR L366234498:id OR L366234503:id OR L366284993:id OR L366300680:id OR L366391051:id OR L368007272:id OR L368013343:id OR L368018038:id OR L368065325:id OR L368081425:id OR L368091797:id OR L368113884:id OR L368127533:id OR L368129774:id OR L368147145:id OR L368226731:id OR L368259257:id OR L368308942:id OR L368326352:id OR L368331159:id OR L368398483:id OR L368405521:id OR L368417943:id OR L368445086:id OR L368490590:id OR L368613820:id OR L368625335:id OR L368695894:id OR L368710621:id OR L368743306:id OR L368758937:id OR L368763377:id OR L368785936:id OR L368798508:id OR L368798602:id OR L368798635:id OR L368801276:id OR L368850149:id OR L368927354:id OR L368952816:id OR L368971695:id OR L369000726:id OR L369031465:id OR L369034671:id OR L369053721:id OR L369059766:id OR L369086186:id OR L369110314:id OR L369184493:id OR L369192267:id OR L369228280:id OR L369244398:id OR L369285639:id OR L369286848:id OR L369320177:id OR L369337260:id OR L369393631:id OR L369409056:id OR L369463955:id OR L369487456:id OR L369535087:id OR L369565785:id OR L369615680:id OR L369634930:id OR L369651483:id OR L369790960:id OR L369819783:id OR L369873455:id OR L369881711:id OR L369884915:id OR L369884917:id OR L369896693:id OR L369903459:id OR L369903999:id OR L369931509:id OR L369933639:id OR L369938365:id OR L369938366:id OR L369951230:id OR L369956523:id OR L369963417:id OR L369965473:id OR L370048629:id OR L370060797:id OR L370103818:id OR L370103823:id OR L370103826:id OR L370103827:id OR L370103829:id OR L370103833:id OR L370114891:id OR L370131747:id OR L370150883:id OR L370183205:id OR L370304416:id OR L370311386:id OR L370318939:id |

| | |
|---|---|
| | OR L370329447:id OR L370341239:id OR L370364147:id OR L370368839:id OR L370379434:id OR L370386815:id OR L370431310:id OR L370485605:id OR L370507261:id OR L372011263:id OR L372059017:id OR L372146425:id OR L372157458:id OR L372167567:id OR L372195203:id OR L372232427:id OR L372317359:id OR L372333514:id OR L372335507:id OR L372361981:id OR L372384643:id OR L372626474:id OR L372658633:id OR L372679125:id OR L372787015:id OR L372796176:id OR L372803653:id OR L372927070:id OR L372988513:id OR L373007650:id OR L373007684:id OR L373015431:id OR L373022991:id OR L373048254:id OR L373050913:id OR L373058584:id OR L373060191:id OR L373128461:id OR L373128558:id OR L373187650:id OR L373193835:id OR L373224407:id OR L373250187:id OR L373412477:id OR L373417821:id OR L373446301:id OR L373491219:id OR L373543060:id OR L373553078:id OR L373678155:id OR L373752258:id OR L373762007:id OR L373762011:id OR L373762012:id OR L373766519:id OR L373780794:id OR L373788613:id OR L373791991:id OR L373796674:id OR L373867295:id OR L373970442:id OR L38480890:id OR L38686937:id OR L39362754:id OR L39642671:id OR L40:id OR L40:id OR L40139589:id OR L40984964:id OR L41150417:id OR L41219442:id OR L41297195:id OR L41359114:id OR L41773901:id OR L43106714:id OR L43329343:id OR L43356058:id OR L43372216:id OR L43372219:id OR L43780485:id OR L44425447:id OR L46439286:id OR L46515140:id OR L46652469:id OR L46830294:id OR L46855952:id OR L47232962:id OR L47244871:id OR L47244892:id OR L47321449:id OR L47423686:id OR L47547720:id OR L50124190:id OR L50209494:id OR L50221243:id OR L50237240:id OR L50380773:id OR L50444307:id OR L50448555:id OR L50543981:id OR L50659899:id OR L50716480:id OR L50740710:id OR L50743941:id OR L50747351:id OR L50830863:id OR L50894436:id OR L50901408:id OR L50905815:id OR L50940547:id OR L50992249:id OR L50993114:id OR L51042816:id OR L51087935:id OR L51092969:id OR L51119491:id OR L51121651:id OR L51124945:id OR L51146756:id OR L51149962:id OR L51182886:id OR L51195272:id OR L51242597:id OR L51271135:id OR L51347196:id OR L51352309:id OR L51363971:id OR L51367070:id OR L51376180:id OR L51406150:id OR L51407771:id OR L51432071:id OR L51459444:id OR L51464312:id OR L51490478:id OR L51518051:id OR L51521081:id OR L51541525:id OR L51553239:id OR L51566180:id OR L51579482:id OR L51600351:id OR L51601576:id OR L51651875:id OR L51678767:id OR L51697409:id OR L51701694:id OR L51721309:id OR L51734154:id OR L51739297:id OR L51757261:id OR L51764320:id OR L51766900:id OR L51768591:id OR L51781169:id OR L51795380:id OR L51796824:id OR L51832971:id OR L51836163:id OR L51855938:id OR L51863307:id OR L51874064:id OR L51887167:id OR L51893218:id OR L51909195:id OR L51934508:id OR L51979385:id OR L51991631:id OR L51995365:id OR L52020877:id OR L52057016:id OR L52059778:id OR L52059779:id OR L52068894:id OR L52073183:id OR L52079925:id OR L52085914:id OR L52087665:id OR L52110465:id OR L52135182:id OR L52140772:id OR L52167068:id OR L52167952:id OR L52172632:id OR L52173720:id OR L52174867:id OR L52191345:id OR L52206857:id OR L52214961:id OR L52248783:id OR L52248790:id OR L52259030:id OR L52276533:id OR L52282499:id OR L52302221:id OR L52328316:id OR L52359666:id OR L52385479:id OR L52400274:id OR L52411458:id OR L52411685:id OR L52425464:id OR L52430696:id OR L52468129:id OR L52477986:id OR L52483937:id OR L52494673:id OR L52510563:id OR L52521308:id OR L52522645:id OR L52549173:id OR L52558810:id OR L52583644:id OR L52601303:id OR L52616856:id OR L52673403:id OR L52715001:id OR L52715240:id OR L52719994:id OR L52730879:id OR L52747987:id OR L52754683:id OR L52783271:id OR L52791827:id OR L52806187:id OR L52806193:id OR L52810299:id OR L52811747:id OR L52822214:id OR L52829203:id OR L52831240:id OR L52843043:id OR L52856293:id OR L52860482:id OR L52872055:id OR L52878435:id OR L52883497:id OR L52886402:id OR L52926761:id OR L52938955:id OR L52943216:id OR L52946215:id OR L52953880:id OR L52989949:id OR L53005518:id OR L53030072:id OR L53040318:id OR L53040819:id OR |

| | |
|---|---|
| | L53070788:id OR L53085547:id OR L53105027:id OR L53110551:id OR L53129302:id OR L53144445:id OR L53145787:id OR L53166059:id OR L53187853:id OR L53196248:id OR L53210322:id OR L53212190:id OR L53231869:id OR L53245565:id OR L53252581:id OR L53272812:id OR L53298707:id OR L53303842:id OR L563069065:id OR L563080947:id OR L60:id OR L60:id OR L600013881:id OR L600069174:id OR L600087061:id OR L600122352:id OR L600123084:id OR L600130788:id OR L600171221:id OR L600192078:id OR L600215231:id OR L600219890:id OR L600223456:id OR L600232202:id OR L600263370:id OR L600277935:id OR L600279637:id OR L600285939:id OR L600311090:id OR L600341252:id OR L600342602:id OR L600487534:id OR L600546982:id OR L600555570:id OR L600570615:id OR L600622021:id OR L600637206:id OR L600648934:id OR L600684490:id OR L600741823:id OR L600749169:id OR L600762247:id OR L600764107:id OR L600777030:id OR L600785131:id OR L600794679:id OR L600982955:id OR L601033588:id OR L601088465:id OR L601108919:id OR L601172183:id OR L601465293:id OR L601499133:id OR L601565052:id OR L601592612:id OR L601698614:id OR L601923956:id OR L601954939:id OR L601969340:id OR L602111909:id OR L602141945:id OR L602152413:id OR L602195084:id OR L602223678:id OR L602223682:id OR L602244213:id OR L602450654:id OR L602458524:id OR L602460432:id OR L602603976:id OR L602645176:id OR L602646204:id OR L602799119:id OR L602811620:id OR L602883832:id OR L602892495:id OR L602970787:id OR L602976312:id OR L602983872:id OR L603235930:id OR L603284195:id OR L603284953:id OR L603464751:id OR L603464774:id OR L603518440:id OR L603524290:id OR L603579721:id OR L603670211:id OR L603860621:id OR L603871188:id OR L603884971:id OR L603887527:id OR L603936890:id OR L603945428:id OR L603991510:id OR L604018796:id OR L604203298:id OR L604231978:id OR L604235219:id OR L604299792:id OR L604312689:id OR L604334548:id OR L604417323:id OR L604424388:id OR L604438990:id OR L604461117:id OR L604592375:id OR L604612185:id OR L604678857:id OR L604710914:id OR L604715147:id OR L604733861:id OR L604767499:id OR L604818855:id OR L604848363:id OR L604848374:id OR L604864790:id OR L604871399:id OR L604883209:id OR L604907859:id OR L604916367:id OR L604954771:id OR L604982541:id OR L605024647:id OR L605076532:id OR L605094239:id OR L605100752:id OR L605147984:id OR L605190771:id OR L605210080:id OR L605254027:id OR L605299379:id OR L605337829:id OR L605417513:id OR L605552356:id OR L605553679:id OR L605565352:id OR L605579706:id OR L605584106:id OR L605674915:id OR L605684348:id OR L605694783:id OR L605701099:id OR L605703451:id OR L605784859:id OR L605803412:id OR L605848441:id OR L605900174:id OR L605985206:id OR L606001920:id OR L606026864:id OR L606031511:id OR L606031553:id OR L606056888:id OR L606077803:id OR L606082526:id OR L606152698:id OR L606170709:id OR L606228538:id OR L606413373:id OR L606422277:id OR L606439656:id OR L606602564:id OR L606684633:id OR L606782418:id OR L606830593:id OR L606941030:id OR L606957370:id OR L606981881:id OR L607065886:id OR L607106687:id OR L607111791:id OR L607149564:id OR L607185574:id OR L607209193:id OR L607226025:id OR L607254090:id OR L607275318:id OR L607394876:id OR L607405713:id OR L607408901:id OR L607412630:id OR L607430490:id OR L607579528:id OR L607627031:id OR L607722570:id OR L607722865:id OR L607766994:id OR L607837339:id OR L607846274:id OR L607878079:id OR L607910516:id OR L607916313:id OR L607971510:id OR L608064505:id OR L608197351:id OR L608233137:id OR L608321564:id OR L608489782:id OR L608646186:id OR L608668499:id OR L608673940:id OR L608695403:id OR L608756543:id OR L608826634:id OR L608831492:id OR L608871815:id OR L608874444:id OR L608964846:id OR L608974822:id OR L608994277:id OR L609033163:id OR L609159330:id OR L609194062:id OR L609206163:id OR L609223045:id OR L609294098:id OR L609389738:id OR L609602973:id OR L609721333:id OR L609867687:id OR L610014521:id OR L610035650:id OR L610250583:id OR L610260174:id OR L610261199:id OR L610329253:id |

| | |
|---|---|
| | OR L610337212:id OR L610343324:id OR L610343957:id OR L610441711:id OR L610457324:id OR L610468555:id OR L610473737:id OR L610557742:id OR L610586692:id OR L610626070:id OR L610658957:id OR L610712422:id OR L610781585:id OR L610834517:id OR L610861890:id OR L610892272:id OR L610895394:id OR L610917918:id OR L610952324:id OR L610969175:id OR L610981013:id OR L610981030:id OR L611017329:id OR L611064651:id OR L611171493:id OR L611224825:id OR L611245326:id OR L611281636:id OR L611391721:id OR L611498330:id OR L611499650:id OR L611530377:id OR L611551810:id OR L611554652:id OR L611584430:id OR L611650229:id OR L611653376:id OR L611668900:id OR L611678068:id OR L611695269:id OR L611841206:id OR L611842082:id OR L611880079:id OR L611908701:id OR L611986636:id OR L611989808:id OR L612009488:id OR L612061171:id OR L612075644:id OR L612097572:id OR L612194438:id OR L612244453:id OR L612249503:id OR L612259107:id OR L612276421:id OR L612320213:id OR L612340250:id OR L612669190:id OR L612682041:id OR L612717416:id OR L612833514:id OR L612833905:id OR L612890642:id OR L612919970:id OR L612948401:id OR L612962422:id OR L612986839:id OR L613106503:id OR L613167891:id OR L613173139:id OR L613222026:id OR L613237376:id OR L613330454:id OR L613346428:id OR L613355227:id OR L613363078:id OR L613483643:id OR L613497915:id OR L613533626:id OR L613545427:id OR L613694797:id OR L613711580:id OR L613734162:id OR L613819061:id OR L613922016:id OR L613941774:id OR L614005526:id OR L614007616:id OR L614008729:id OR L614022418:id OR L614099067:id OR L614099391:id OR L614133776:id OR L614146526:id OR L614181168:id OR L614223151:id OR L614242043:id OR L614263388:id OR L614285409:id OR L614298896:id OR L614303425:id OR L614330644:id OR L614336128:id OR L614350729:id OR L614361497:id OR L614390599:id OR L614427347:id OR L614460569:id OR L614476262:id OR L614476430:id OR L614493625:id OR L614515287:id OR L614565625:id OR L614574026:id OR L614604153:id OR L614635694:id OR L614643384:id OR L614748536:id OR L614767849:id OR L614787251:id OR L614823066:id OR L614850939:id OR L614865805:id OR L614923384:id OR L614928375:id OR L615072977:id OR L615082913:id OR L615100545:id OR L615125703:id OR L615161967:id OR L615175555:id OR L615175899:id OR L615183815:id OR L615190662:id OR L615256561:id OR L615299812:id OR L615330399:id OR L615340872:id OR L615489484:id OR L615616929:id OR L615664391:id OR L615702810:id OR L615765916:id OR L615775375:id OR L615775818:id OR L615873090:id OR L615926589:id OR L615959355:id OR L615976256:id OR L616053451:id OR L616065600:id OR L616104379:id OR L616104549:id OR L616109827:id OR L616110039:id OR L616163969:id OR L616222548:id OR L616272859:id OR L616274815:id OR L616403252:id OR L616404215:id OR L616441491:id OR L616477117:id OR L616484309:id OR L616486783:id OR L616490452:id OR L616707571:id OR L616716460:id OR L616786422:id OR L616850672:id OR L616866006:id OR L616914282:id OR L616956496:id OR L616995130:id OR L617070582:id OR L617085834:id OR L617180728:id OR L617183204:id OR L617279617:id OR L617314986:id OR L617314997:id OR L617351137:id OR L617400133:id OR L617420845:id OR L617423407:id OR L617459523:id OR L617460510:id OR L617474084:id OR L617547324:id OR L617703290:id OR L617781438:id OR L617838383:id OR L617980928:id OR L618014975:id OR L618026058:id OR L618047749:id OR L618057077:id OR L618058051:id OR L618071287:id OR L618155124:id OR L618203829:id OR L618229050:id OR L618276881:id OR L618291661:id OR L618433327:id OR L618437401:id OR L618450488:id OR L618451203:id OR L618473384:id OR L618501732:id OR L618551681:id OR L618555251:id OR L618603721:id OR L618618967:id OR L618620503:id OR L618661640:id OR L618752532:id OR L618790969:id OR L618803591:id OR L618941883:id OR L619021758:id OR L619029531:id OR L619042207:id OR L619046898:id OR L619090971:id OR L619161605:id OR L619164266:id OR L619205889:id OR L619206142:id OR L619225355:id OR L619233647:id OR L619238888:id OR L619255882:id OR L619301288:id |

| | |
|---|---|
| | OR L619305324:id OR L619348446:id OR L619425452:id OR L619427132:id OR L619431365:id OR L619483462:id OR L619487499:id OR L619625705:id OR L619644900:id OR L619661100:id OR L619665896:id OR L619678341:id OR L619707545:id OR L619864761:id OR L619898251:id OR L619904479:id OR L620020458:id OR L620025395:id OR L620026586:id OR L620115135:id OR L620160707:id OR L620178992:id OR L620257505:id OR L620258175:id OR L620329571:id OR L620330339:id OR L620338256:id OR L620374511:id OR L620378607:id OR L620380321:id OR L620519305:id OR L620535359:id OR L620542050:id OR L620557062:id OR L620604858:id OR L620624536:id OR L620676727:id OR L620684642:id OR L620757310:id OR L620850618:id OR L621002016:id OR L621060022:id OR L621125996:id OR L621164156:id OR L621195877:id OR L621200743:id OR L621211566:id OR L621302440:id OR L621340421:id OR L621392904:id OR L621428568:id OR L621441813:id OR L621468013:id OR L621484433:id OR L621484450:id OR L621543270:id OR L621575174:id OR L621598364:id OR L621599857:id OR L621622320:id OR L621685316:id OR L621707083:id OR L621771959:id OR L621862786:id OR L621897284:id OR L621949213:id OR L622045547:id OR L622049753:id OR L622058760:id OR L622067604:id OR L622104240:id OR L622161262:id OR L622161317:id OR L622203706:id OR L622236885:id OR L622240453:id OR L622289331:id OR L622340700:id OR L622389579:id OR L622400549:id OR L622410080:id OR L622433748:id OR L622636501:id OR L622646893:id OR L622647064:id OR L622647260:id OR L622658447:id OR L622688197:id OR L622688362:id OR L622738610:id OR L622755023:id OR L622766724:id OR L622777394:id OR L622814644:id OR L622827400:id OR L622916009:id OR L622943791:id OR L622948105:id OR L622984854:id OR L622984898:id OR L622997207:id OR L623029864:id OR L623038714:id OR L623039355:id OR L623083416:id OR L623175669:id OR L623190518:id OR L623233885:id OR L623234410:id OR L623361731:id OR L623427374:id OR L623433848:id OR L623544884:id OR L623546089:id OR L623546258:id OR L623626998:id OR L623633775:id OR L623653343:id OR L623722950:id OR L623729076:id OR L623813737:id OR L623816637:id OR L623826714:id OR L623847767:id OR L623853205:id OR L623853299:id OR L623933521:id OR L623938355:id OR L623981616:id OR L623996897:id OR L624033983:id OR L624064462:id OR L624102295:id OR L624159967:id OR L624162860:id OR L624202882:id OR L624222367:id OR L624272625:id OR L624283430:id OR L624286184:id OR L624294237:id OR L624297739:id OR L624346202:id OR L624409922:id OR L624430404:id OR L624434199:id OR L624440691:id OR L624463034:id OR L624498657:id OR L624522493:id OR L624556064:id OR L624611402:id OR L624629215:id OR L624632698:id OR L624654560:id OR L624748575:id OR L624752092:id OR L624760947:id OR L625004289:id OR L625018121:id OR L625033634:id OR L625061348:id OR L625068017:id OR L625090799:id OR L625095546:id OR L625193738:id OR L625204340:id OR L625271286:id OR L625290731:id OR L625291558:id OR L625323396:id OR L625513149:id OR L625527611:id OR L625530932:id OR L625535182:id OR L625557221:id OR L625558018:id OR L625585246:id OR L625585259:id OR L625629033:id OR L625672991:id OR L625726005:id OR L625732052:id OR L625830692:id OR L625853347:id OR L625860945:id OR L625903398:id OR L626052960:id OR L626082483:id OR L626124967:id OR L626126921:id OR L626177069:id OR L626189755:id OR L626206335:id OR L626218020:id OR L626219616:id OR L626231327:id OR L626238132:id OR L626271137:id OR L626296926:id OR L626300234:id OR L626366033:id OR L626442030:id OR L626452211:id OR L626510812:id OR L626566450:id OR L626668618:id OR L626674647:id OR L626723663:id OR L626814196:id OR L626872752:id OR L626924485:id OR L626988709:id OR L627011373:id OR L627177906:id OR L627218559:id OR L627234389:id OR L627235494:id OR L627272173:id OR L627372762:id OR L627389133:id OR L627405185:id OR L627418462:id OR L627425977:id OR L627452521:id OR L627473337:id OR L627525751:id OR L627559486:id OR L627590451:id OR L627617366:id OR L627686954:id OR L627793592:id OR L627805977:id OR L627808699:id |

| | |
|---|---|
| | OR L627845736:id OR L627865239:id OR L627929403:id OR L627955695:id OR L627960369:id OR L628043493:id OR L628046688:id OR L628086811:id OR L628126220:id OR L628128346:id OR L628151794:id OR L628184594:id OR L628214602:id OR L628224880:id OR L628233419:id OR L628252077:id OR L628278978:id OR L628371388:id OR L628456649:id OR L628490338:id OR L628649068:id OR L628659600:id OR L628685967:id OR L628687045:id OR L628699272:id OR L628714576:id OR L628857845:id OR L628903701:id OR L628915346:id OR L628924017:id OR L628924268:id OR L629089109:id OR L629111291:id OR L629304013:id OR L629351702:id OR L629458883:id OR L629462604:id OR L629462917:id OR L629525723:id OR L629580538:id OR L629587554:id OR L629638341:id OR L629657928:id OR L629693773:id OR L629698630:id OR L629732396:id OR L629775932:id OR L629780604:id OR L629791821:id OR L629796077:id OR L629870493:id OR L629875583:id OR L629876110:id OR L629917088:id OR L630022971:id OR L630031973:id OR L630038819:id OR L630045301:id OR L630056923:id OR L630246307:id OR L630286089:id OR L630286324:id OR L630286774:id OR L630287095:id OR L630287727:id OR L630288792:id OR L630288907:id OR L630601478:id OR L630814785:id OR L630823025:id OR L630934639:id OR L630934689:id OR L630982999:id OR L630992209:id OR L631005548:id OR L631005658:id OR L631025457:id OR L631043601:id OR L631052038:id OR L631090729:id OR L631201970:id OR L631202012:id OR L631223482:id OR L631228993:id OR L631235517:id OR L631238264:id OR L631241731:id OR L631258386:id OR L631265777:id OR L631283290:id OR L631286601:id OR L631286946:id OR L631289965:id OR L631294114:id OR L631334001:id OR L631352467:id OR L631358847:id OR L631368185:id OR L631368917:id OR L631379563:id OR L631406243:id OR L631411235:id OR L631412741:id OR L631419403:id OR L631419914:id OR L631423447:id OR L631423920:id OR L631433717:id OR L631443738:id OR L631445339:id OR L631480747:id OR L631482200:id OR L631522017:id |
| #19 | #15 NOT (#17 OR #18) |

**Table 1c: APA PsycInfo® search strategy**

| | |
|---|---|
| **Provider/Interface** | Ovid |
| **Database** | APA PsycInfo® |
| **Date searched** | April 29, 2020 |
| **Database update** | 1806 to April Week 3 2020 |
| **Search developer(s)** | Helena M. VonVille |
| **Limit to English?** | Yes |
| **Date Range** | No limit by date |
| **Publication Types** | Journal articles only |
| **Search filter source** | No search filter used |

| **Note:** All items were downloaded to EndNote and duplicates removed. | |
|---|---|
| 1 | ((brain derived neurotrophic factor/ or (brain-derived neurotrophic factor or BDNF or val66met).ti,ab,id.) and (microrna/ or (epigenetic or epigenetics or epigenomic or epigenomics or hypermethylation or hypermethylated or hypermethylation or methylation or methylated or methylation or micro rna* or microrna* or mirna* or non-coding rna or noncoding rna).ti,ab,id.)) or (Histone* and (modification or acetylation or acetylated or deacetylation)).ti,ab,id. |
| 2 | 1 not (animal.po. not (animal.po. and human.po.)) |
| 3 | exp central nervous system disorders/ or cognitive neuroscience/ or exp mental disorders/ |
| 4 | (abuse or abused or abuser or abusers or abusive or abusively or abusiveness or adhd or Adjustment Disorder* or adversities or adversity or Affective disorder* or aging or Agoraphobia or Alcoholism or alzheimer* or Amnesia or Anorexia or antisocial behavior or Anxiety or anxiousness or Aphasia or ASPD or Attachment disorder* or Attention Deficit or Avoidant Restrictive Food Intake or Behavior or Binge-Eating or Bipolar or Body Dysmorphi* or Borderline Personality or Brain cancer* or brain injury or Brain neoplasm* or Brain Stem cancer* or Brain Stem Neoplasm* or Brain Stem tumor* or Brain Stem tumour* or Brain tumor* or Brain tumour* or Bulimia or Capgras or Cerebellar cancer* or Cerebellar Neoplasm* or Cerebellar tumor* or Cerebellar tumour* or Cerebral Ventricle cancer* or Cerebral Ventricle Neoplasm* or Cerebral Ventricle tumor* or Cerebral Ventricle tumour* or Child Development Disorder* or Choroid Plexus cancer* or Choroid Plexus Neoplasm* or Choroid Plexus tumor* or Choroid Plexus tumour* or Cognition or cognitive or Communication Disorder* or Compulsive Personality or Conduct Disorder* or Consciousness Disorder* or Conversion disorder* or Creutzfeldt-Jakob or Cyclothymic or Delirium or |

| | |
|---|---|
| | Delusional Parasitosis or dementia or Dependent Personality or deployment or depression or depressive or deprivation or deprive or deprived or deprivement or Developmental Disabilities or Diabulimia or dissociation or Dissociative Disorder* or Dissociative Identity Disorder* or domestic violence or drug abuse or Dyslexia or dyslexic or Dyspareunia or Dyssomnias or early-life stress or Eating disorder* or Elimination Disorder* or ELS or emotion regulation or emotional abuse or emotional neglect or emotionally abused or emotionally neglected or Encopresis or Enuresis or epilepsy or Epilepsy or Epileptic or Erectile Dysfunction or Exhibitionism or Factitious disorder* or family violence or fetal alcohol syndrom or Fetishism or Firesetting or Food Addiction or Frontotemporal or Gambling or Gender Disorder* or Gender Dysphoria or Glioma* or Huntington* or Hypochondriasis or Infratentorial cancer* or Infratentorial Neoplasm* or Infratentorial tumor* or Infratentorial tumour* or Intellectual Disability or ischemia or Kluver-Bucy* or Learning Disabilities or Lewy Body* or maltreat or maltreated or maltreatment or Masochism or mdd or mental disorder* or mental illness or minDD or Mood disorder* or Morgellon* or Motor disorder* or Motor Skills disorder* or Mutism or negative experiences or neglected or Neonatal Abstinence Syndrome or Neurasthenia or Neurocirculatory Asthenia or Neurocognitive disorder* or Neurocytoma or Neurodevelopmental disorder* or neurological disorder* or neuropsychological or neuroses or Neurotic or Night Eating or Obsessive-Compulsive or ocd or Panic or paranoia or Paranoid or Paraphilic or Parasomnias or Parkinson* or Pedophilia or Personality Disorder* or phobia* or Phobic or Pica or Pinealoma or posttraumatic or post-traumatic or Premature Ejaculation or preterm birth or psychiatric or psychological or psychopathology or Psychoses or psychosis or psychosocial or psychotic or PTSD or Rumination Syndrome or Sadism or Schizoid or Schizophrenia or schizophrenic or sexual abuse or Sexual disorder* or Sexual Dysfunction* or Sleep Disorder* or Somatoform or Stereotypic Movement disorder* or stress or stressful or stressor or stroke or Substance Abuse or suicidal or suicide or Tic or tics or trauma* or Trichotillomania or Vaginismus or Voyeurism).ti,ab,id. |
| 5 | 3 or 4 |
| 6 | 2 and 5 and english.la. |
| 7 | limit 6 to all journals |

**Table 1d: Cochrane Protocols search strategy**

| | |
|---|---|
| **Provider/Interface** | Wiley |
| **Database** | Cochrane Protocols |
| **Date searched** | April 30, 2020 |
| **Database update** | April 30, 2020 |
| **Search developer(s)** | Helena M. VonVille |
| **Limit to English?** | No |
| **Date Range** | No limit by date |
| **Publication Types** | No limit by publication type |
| **Search filter source** | No search filter used |

| | |
|---|---|
| 1 | (brain-derived neurotrophic factor OR brain-derived neurotrophic factors OR BDNF OR val66met) |
